# The accuracy of novel antigen rapid diagnostics for SARS-CoV-2: a living systematic review and meta-analysis

**DOI:** 10.1101/2021.02.26.21252546

**Authors:** Lukas E. Brümmer, Stephan Katzenschlager, Mary Gaeddert, Christian Erdmann, Stephani Schmitz, Marc Bota, Maurizio Grilli, Jan Larmann, Markus A. Weigand, Nira R. Pollock, Aurélien Macé, Sergio Carmona, Stefano Ongarello, Jilian A. Sacks, Claudia M. Denkinger

## Abstract

**Background:** SARS-CoV-2 antigen rapid diagnostic tests (Ag-RDTs) are increasingly being integrated in testing strategies around the world. Studies of the Ag-RDTs have shown variable performance. In this systematic review and meta-analysis, we assessed the clinical accuracy (sensitivity and specificity) of commercially available Ag-RDTs.

**Methods and Results:** We registered the review on PROSPERO (Registration number: CRD42020225140). We systematically searched multiple databases (PubMed, Web of Science Core Collection, medRvix and bioRvix, FIND) for publications evaluating the accuracy of Ag-RDTs for SARS-CoV-2 up until April 30^th^, 2021. Descriptive analyses of all studies were performed and when more than four studies were available, a random-effects meta-analysis was used to estimate pooled sensitivity and specificity in comparison to reverse transcriptase polymerase chain reaction testing. We assessed heterogeneity by subgroup analyses, and rated study quality and risk of bias using the QUADAS 2 assessment tool. From a total of 14,254 articles, we included 133 analytical and clinical studies resulting in 214 clinical accuracy data sets with 112,323 samples. Across all meta-analyzed samples, the pooled Ag-RDT sensitivity was 71.2% (95% confidence interval [CI] 68.2 to 74.0) and increased to 76.3% (CI 73.1 to 79.2) if analysis was restricted to studies that followed the Ag-RDT manufacturers’ instructions. The LumiraDx showed the highest sensitivity with 88.2% (CI 59.0 to 97.5). Of instrument-free Ag-RDTs, Standard Q nasal performed best with 80.2% sensitivity (CI 70.3 to 87.4). Across all Ag-RDTs sensitivity was markedly better on samples with lower Ct-values, i.e., <20 (96.5%, CI 92.6 to 98.4) and <25 (95.8%, CI 92.3 to 97.8), in comparison to those with Ct ≥25 (50.7%, CI 35.6 to 65.8) and ≥30 (20.9%, CI 12.5 to 32.8). Testing in the first week from symptom onset resulted in substantially higher sensitivity (83.8%, CI 76.3 to 89.2) compared to testing after one week (61.5%, CI 52.2 to 70.0). The best Ag-RDT sensitivity was found with anterior nasal sampling (75.5%, CI 70.4 to 79.9) in comparison to other sample types (e.g., nasopharyngeal 71.6%, CI 68.1 to 74.9) although CIs were overlapping. Concerns of bias were raised across all data sets, and financial support from the manufacturer was reported in 24.1% of data sets. Our analysis was limited by the included studies’ heterogeneity in design and reporting, making it difficult to draw conclusions from.

**Conclusion:** In this study we found that Ag-RDTs detect the vast majority of cases within the first week of symptom onset and those with high viral load. Thus, they can have high utility for diagnostic purposes in the early phase of disease, making them a valuable tool to fight the spread of SARS-CoV-2. Standardization in conduct and reporting of clinical accuracy studies would improve comparability and use of data.

**AUTHOR SUMMARY:** Why was this study done?

– Antigen rapid diagnostic tests (Ag-RDTs) are considered an important diagnostic tool to fight the spread of SARS-CoV-2
– An increasing number of Ag-RDTs is offered on the market, and a constantly growing body of literature evaluating their performance is available
– To inform decision makers about the best test to choose, an up to date summary of their performance is needed

What did the researchers do and find?

– On a weekly basis, we search multiple data bases for evaluations of Ag-RDTs detecting SARS-CoV-2 and post the results on www.diagnosticsglobalhealth.org
– Based on the search results up until April 30^th^, 2021, we conducted a systematic review and meta-analysis, including a total of 133 clinical and analytical accuracy studies
– Across all meta-analyzed studies, when Ag-RDTs were performed according to manufacturers’ recommendations, they showed a sensitivity of 76.3% (CI 73.1 to 79.2), with the LumiraDx (sensitivity 88.2%, CI 59.0 to 97.5) and of the instrument-free Ag-RDT Standard Q (74.9% sensitivity, CI 69.3 to 79.7) performing best.
– Across all Ag-RDTs, sensitivity increased to 95.8% (CI 92.3 to 97.8) when restricting the analysis to samples with high viral loads (i.e., a Ct-value <25) and to 83.8% (CI 76.3 to 89.2) when tests were performed on patients within the first week after symptom onset

What do these findings mean?

– Ag-RDTs detect the vast majority of cases within the first week of symptom onset and those with high viral load. Thus, they can have high utility for diagnostic purposes in the early phase of disease
– Out of all assessed tests, the Lumira Dx showed the highest accuracy. The Standard Q was the best performing test when only considering those that don’t require an instrument
– A standardization of reporting methods for clinical accuracy studies would enhance future test-comparisons

## INTRODUCTION

As the COVID-19 pandemic continues around the globe, antigen rapid diagnostic tests (Ag-RDTs) for SARS-CoV-2 are seen as an important diagnostic tool to fight the virus’ spread [1, 2]. The number of Ag-RDTs on the market is increasing constantly [3]. Initial data from independent evaluations suggests that the performance of SARS-CoV-2 Ag-RDTs may be lower than what is reported by the manufacturers. In addition, Ag-RDT accuracy seems to vary substantially between tests [4–6].

With the increased availability of Ag-RDTs, an increasing number of independent validations have been published. Such evaluations differ widely in their quality, methods and results, making it difficult to assess the true performance of the respective tests [7]. To inform decision makers on the best choice of individual tests, an aggregated, widely available and frequently updated assessment of the quality, performance and independence of the data is urgently necessary. While other systematic reviews have been published, they only include data up until Nov 2020 [8–11], exclude preprints [12], or were industry sponsored [13]. In addition, only one assessed the quality of studies in detail, with data up until Nov, 2020 [7, 11].

With our systematic review and meta-analysis, we aim to close this gap in the literature and link to a website (www.diagnosticsglobalhealth.org) that is regularly updated.

## METHODS

We developed a study protocol following standard guidelines for systematic reviews [14, 15], which is available in the Supplement (S15). We have also completed the PRISMA checklist, which can be found in the Supplement (S1_PRISMA_Checklists) as well. Furthermore, we registered the review on PROSPERO (Registration number: CRD42020225140).

### SEARCH STRATEGY

We performed a search of the databases PubMed, Web of Science, medRxiv and bioRxiv using search terms that were developed with an experienced medical librarian (MauG) using combinations of subject headings (when applicable) and text-words for the concepts of the search question. The main search terms were “Severe Acute Respiratory Syndrome Corona-virus 2”, “COVID-19”, “Betacoronavirus”, “Coronavirus” and “Point of Care Testing”. The full list of search terms is available in the Supplement (S2). We also searched the FIND website (https://www.finddx.org/sarscov2-eval-antigen/) for relevant studies manually. We performed the search up until April 30^th^, 2021. No language restrictions were applied.

### INCLUSION CRITERIA

We included studies evaluating the accuracy of commercially available Ag-RDTs to establish a diagnosis of a SARS-CoV-2 infection against reverse transcriptase polymerase chain reaction (RT-PCR) or cell culture as reference standard. We included all study populations irrespective of age, presence of symptoms, or the study location. We considered cohort studies, nested cohort studies, case-control or cross-sectional studies and randomized studies. We included both peer reviewed publications and preprints.

We excluded studies in which patients were tested for the purpose of monitoring or ending quarantine. Also, publications with a population size smaller than 10 were excluded. Although the size threshold of 10 is arbitrary, such small studies are more likely to give unreliable estimates of sensitivity or specificity.

### INDEX TESTS

Ag-RDTs for SARS-CoV-2 aim to detect infection by recognizing viral proteins. Most Ag-RDTs use specific labeled antibodies attached to a nitrocellulose matrix strip, to capture the virus antigen. Successful binding of the antibodies to the antigen is either detected visually (through the appearance of a line on the matrix strip (lateral flow assay)) or requires a specific reader for fluorescence detection. Microfluidic enzyme-linked immunosorbent assays have also been developed. Ag-RDTs typically provide results within 10 to 30 minutes [6].

### REFERENCE STANDARD

Viral culture detects viable virus that is relevant for transmission but is available in research settings only. Since RT-PCR tests are more widely available and SARS-CoV-2 RNA (as reflected by RT-PCR cycle threshold (Ct) value) highly correlates with SARS-CoV-2 antigen quantities, we considered it an acceptable reference standard for the purposes of this systematic review [16]. It is of note that there is currently no international standard for the classification of viral load available.

### STUDY SELECTION AND DATA EXTRACTION

Two reviewers (LEB and CE, LEB and SS or LEB and MB) reviewed the titles and abstracts of all publications identified by the search algorithm independently, followed by a full-text review for those eligible, to select the articles for inclusion in the systematic review. Any disputes were solved by discussion or by a third reviewer (CMD).

A full list of the parameters extracted is included in the Supplement (S14) and the data extraction file is available upon request. Studies that assessed multiple Ag-RDTs or presented results based on differing parameters (e.g., various sample types) were considered as individual data sets.

At first, four authors (SK, CE, SS, MB) extracted five randomly selected papers in parallel to align data extraction methods. Afterwards, data extraction and the assessment of methodological quality and independence from test manufacturers (see below) were performed by one author per paper (SK, CE, SS, MB) and controlled by a second (LEB, SK, SS, MB). Any differences were resolved by discussion or by consulting a third author (CMD).

### STUDY TYPES

We differentiated between clinical accuracy studies (performed on clinical samples) or analytical accuracy studies (performed on spiked samples with a known quantity of virus). Analytical accuracy studies can differ widely in methodology, impeding an aggregation of their results. Thus, while we extracted the data for both kinds of studies, we only considered data from clinical accuracy studies as eligible for the meta-analysis. Separately, we summarized the results of analytical studies and compared them with the results of the meta-analysis for individual tests.

### ASSESSMENT OF METHODOLOGICAL QUALITY

The quality of the clinical accuracy studies was assessed by applying the QUADAS-2 tool [17]. The tool evaluates four domains: patient selection, index test, reference standard, and flow and timing. For each domain, the risk of bias is analyzed using different signaling questions. Beyond the risk of bias, the tool also evaluates the applicability of the study of each included study to the research question for every domain. The QUADAS 2 tool was adjusted to the needs of this review and can be found in the Supplement (S3).

### ASSESSMENT OF INDEPENDENCY FROM MANUFACTURERS

We examined whether a study received financial support from a test manufacturer (including the free provision of Ag-RDTs), whether any study author was affiliated with a test manufacturer, or a respective conflict of interest was declared. Studies were judged not to be independent from the test manufacturers if at least one of these aspects was present, otherwise they were considered to be independent.

### STATISTICAL ANALYSIS AND DATA SYNTHESIS

We extracted raw data from the studies and recalculated performance estimates where possible based on the extracted data. The raw data can be found in the Supplement (S4). We prepared forest plots for the sensitivity and specificity of each test and visually evaluated the heterogeneity between studies. If four or more data sets were available with at least 20 positive RT-PCR samples per data set for a predefined analysis, a meta-analysis was performed. We report point estimates of sensitivity and specificity for SARS-CoV-2 detection compared to the reference standard along with 95% confidence intervals (CI) using a bivariate model (implemented with the ‘reitsma’ command from the R package “mada” version 0.5.10). When there were less than four studies for an index test, only a descriptive analysis was performed and accuracy ranges were reported. In sub-group analyses where papers presented data only on sensitivity, a univariate random-effects inverse variance metaanalysis was done (using the ‘metagen’ command from the R package “meta” version 4.11-0). We predefined the following subgroups for meta-analysis: by Ct-value range, by sampling and testing procedure in accordance with manufacturer’s instructions as detailed in the instructions for use (henceforth called IFU-conforming) vs. not IFU-conforming, age (<18; ≥18), sample type, by presence or absence of symptoms, symptom duration (<7 days vs. ≥7 days), by viral load and by type of RT-PCR used.

In an effort to use as much of the heterogeneous data as possible, the cut-offs for the Ct-value groups were relaxed by 2-3 points within each range. The <20 group included values reported up to ≤20, the <25 group included values reported as ≤24 or <25 or 20-25, the <30 group included values from ≤29 to ≤33 and 25-30. The ≥25 group included values reported as ≥25 or 25-30, the ≥30 group included values from ≥30 to ≥35. For categorization by sample type, we assessed (1) nasopharyngeal (NP) alone or combined with other (e.g., oropharyngeal (OP)), (2) OP alone, (3) anterior nasal or mid-turbinate (AN/MT), (4) a combination of bronchial alveolar lavage and throat wash (BAL/TW) or (5) saliva. For categorization by age, the pediatric group included values reported as <16 or <18, whereas values reported as ≥16 or ≥18 were included in the adult group. Analyses were preformed using R 4.0.3 (R Foundation for Statistical Computing, Vienna, Austria).

We aimed to do meta-regression to examine the impact of covariates including symptom duration and Ct-value range. We also performed the Deeks’ test for funnel-plot asymmetry as recommended to investigate publication bias for diagnostic test accuracy meta-analyses ([18], using the ‘midas’ command in Stata version 15); a p-value< 0.10 for the slope coefficient indicates significant asymmetry.

### SENSITIVITY ANALYSIS

Two types of sensitivity analyses were planned: an estimation of sensitivity and specificity excluding case-control studies, and estimation of sensitivity and specificity excluding non-peer-reviewed studies. We compared the results of each sensitivity analysis against overall results to assess the potential bias introduced by considering case-control studies and non-peer reviewed studies.

## RESULTS

### SUMMARY OF STUDIES

The systematic search resulted in 14,254 articles. After removing duplicates, 8,921 articles were screened, and 266 papers were considered eligible for full-text review. Of these, 148 were excluded because they did not present primary data [13,19–131] or the Ag-RDT was not commercially available [16,132–164], leaving 133 studies to be included in the systematic review (Fig 1) [4,165–296].

**Figure 1.**
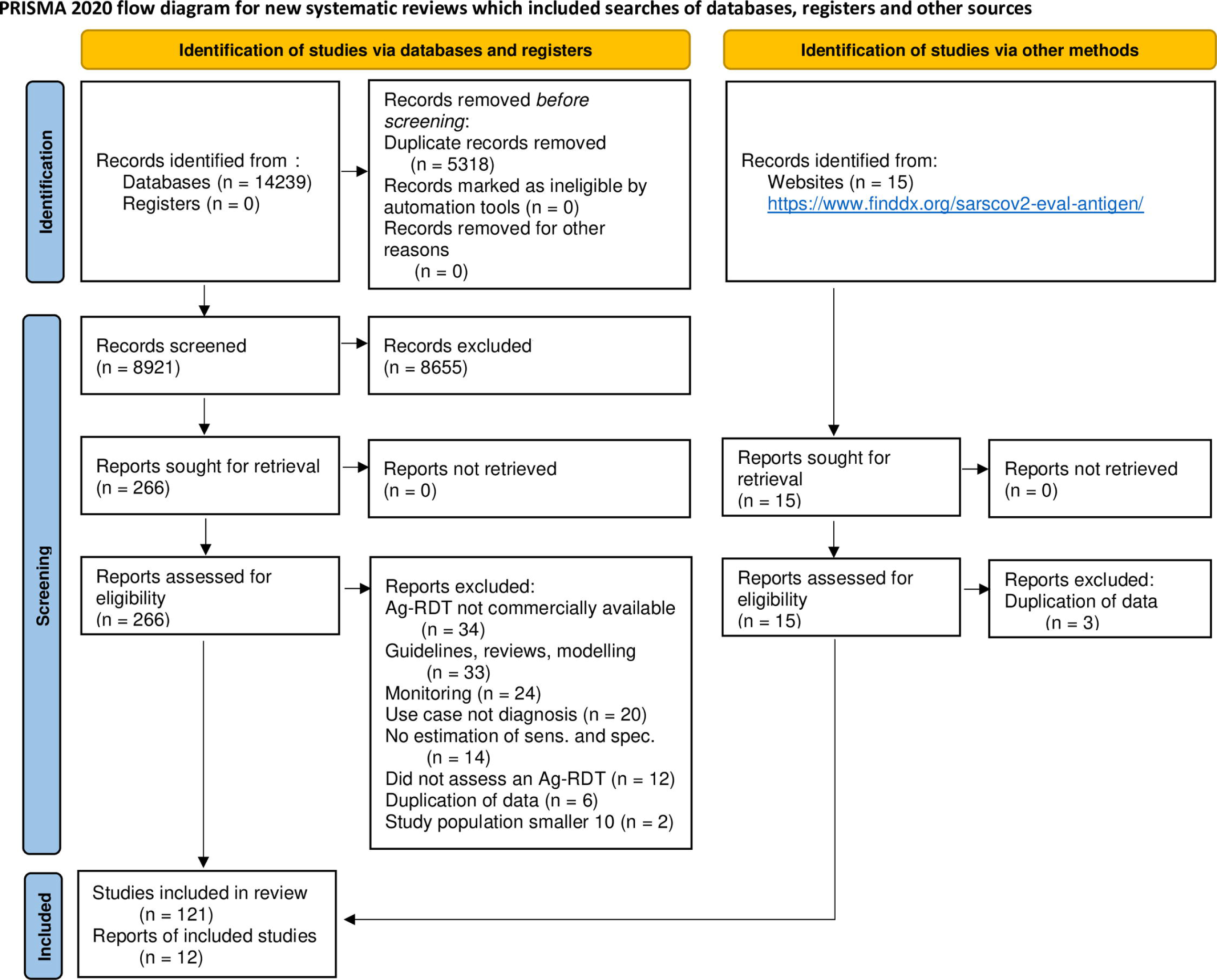

At the end of the data extraction process, 37 studies were still in preprint form [4,171,173,174,177,180,190,192,201,204,205,207,211,214–216,218,220,222,223,225,227,231,233,234,238,240,244,247,253,257,265,267,284,287,290,293]. All studies were written in English, except for two in Spanish [175, 280]. Out of the 133 studies, nine reported analytical accuracy [173,191,198,208,227,256,274,275,282] and the remaining 124 reported clinical accuracy.

The clinical accuracy studies were divided into 214 data sets, while the nine analytical accuracy studies accounted for 62 data sets. A total of 61 different Ag-RDTs were evaluated (48 lateral flow with visual readout, twelve requiring an automated reader), with 56 being assessed in a clinical accuracy study. 39 studies reported data for more than one test and 19 studies of these conducted a head-to-head assessment, i.e., testing at least two Ag-RDTs on the same sample or participant. The reference method was RT-PCR in all except one study, which used viral culture [281].

The most common reasons for testing were the occurrence of symptoms (55/19.9% of data sets), screening independent of symptoms (19/6.9%) and close contact to a SARS-CoV-2 confirmed case (10/3.6%). In 79 (28.6%) of the data sets, persons were tested due to more than one of the reasons mentioned before and for 163 (59.1%) the reason for testing was unclear.

In total, 113,242 Ag-RDTs were performed, 112,323 (99.2%) in clinical accuracy studies and 919 (0.8%) in analytical accuracy studies. In the clinical accuracy studies, the mean number of samples per study was 525 (Range 16 to 6,954). Only 4,752 (4.2%) tests were performed on pediatric samples and 21,351 (18.9%) on samples from adults. For the remaining 87,139 (76.9%) samples it was not specified whether they originate from adults or children. Symptomatic patients comprised 36,981 (32.7%) samples, 32,799 (29.0%) samples originated from asymptomatic patients, and for 42,462 (38.4%) samples the patient’s symptom status could not be identified. The most common sample type evaluated was NP and mixed NP/OP (67,036 samples, 59.2%), followed by AN/MT (27,045 samples, 23.9%). There was substantially less testing done for the other sample types, with 6,254 (5.5%) tests done from OP samples, 1,351 (1.2%) from saliva, 219 (0.2%) from BAL/TW and for 11,337 (10.0%) tests we could not tell the type of sample.

Of the data sets assessing clinical accuracy, 89 (41.6%) performed testing according to the manufacturers’ recommendations (i.e., IFU-conforming), while 100 (46.7%) were not IFU-conforming and for 25 (11.7%) it was unclear. The most common deviations from the IFU were (1) use of samples that were prediluted in transport media not recommended by the manufacturer (80 data sets; seven unclear), (2) use of banked samples (60 data sets; 14 unclear) and (3) a sample type that was not recommended for Ag-RDTs (17 data sets; 8 unclear).

A summary of the tests evaluated in clinical accuracy studies, including study author, sample size, sample type, sample condition and IFU conformity, can be found in Table 1. The Panbio test by Abbott Rapid Diagnostics (Germany; henceforth called Panbio) was reported the most frequently with 39 (18.2%) data sets and 28,089 (25.0%) tests, while Standard Q test by SD Biosensor (South Korea; distributed in Europe by Roche, Germany; henceforth called Standard Q) was assessed in 37 (17.3%) data sets with 16,820 (15.0%) tests performed. Detailed results for each clinical accuracy study are available in the Supplement (S 4).

**Table 1:**
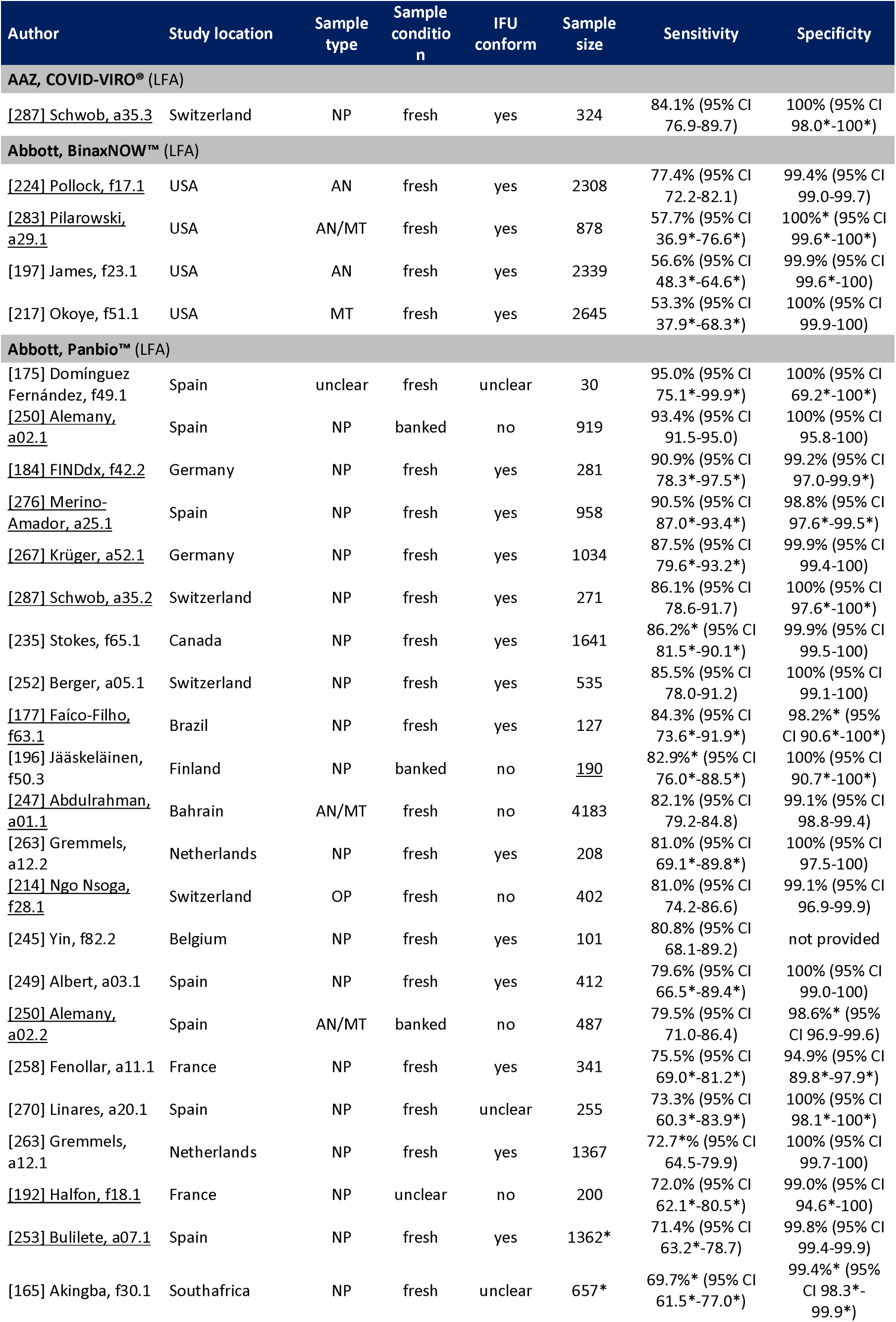
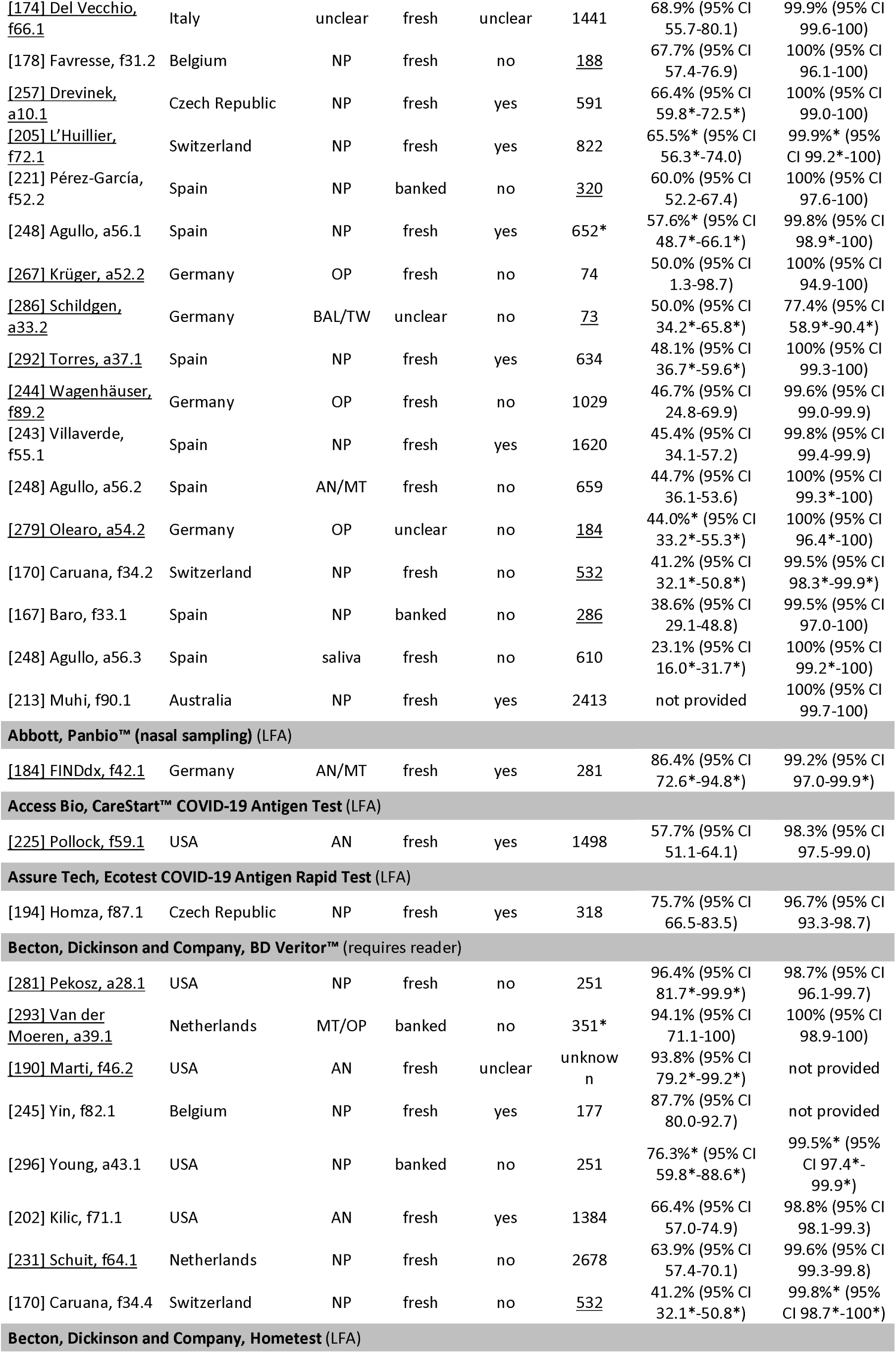
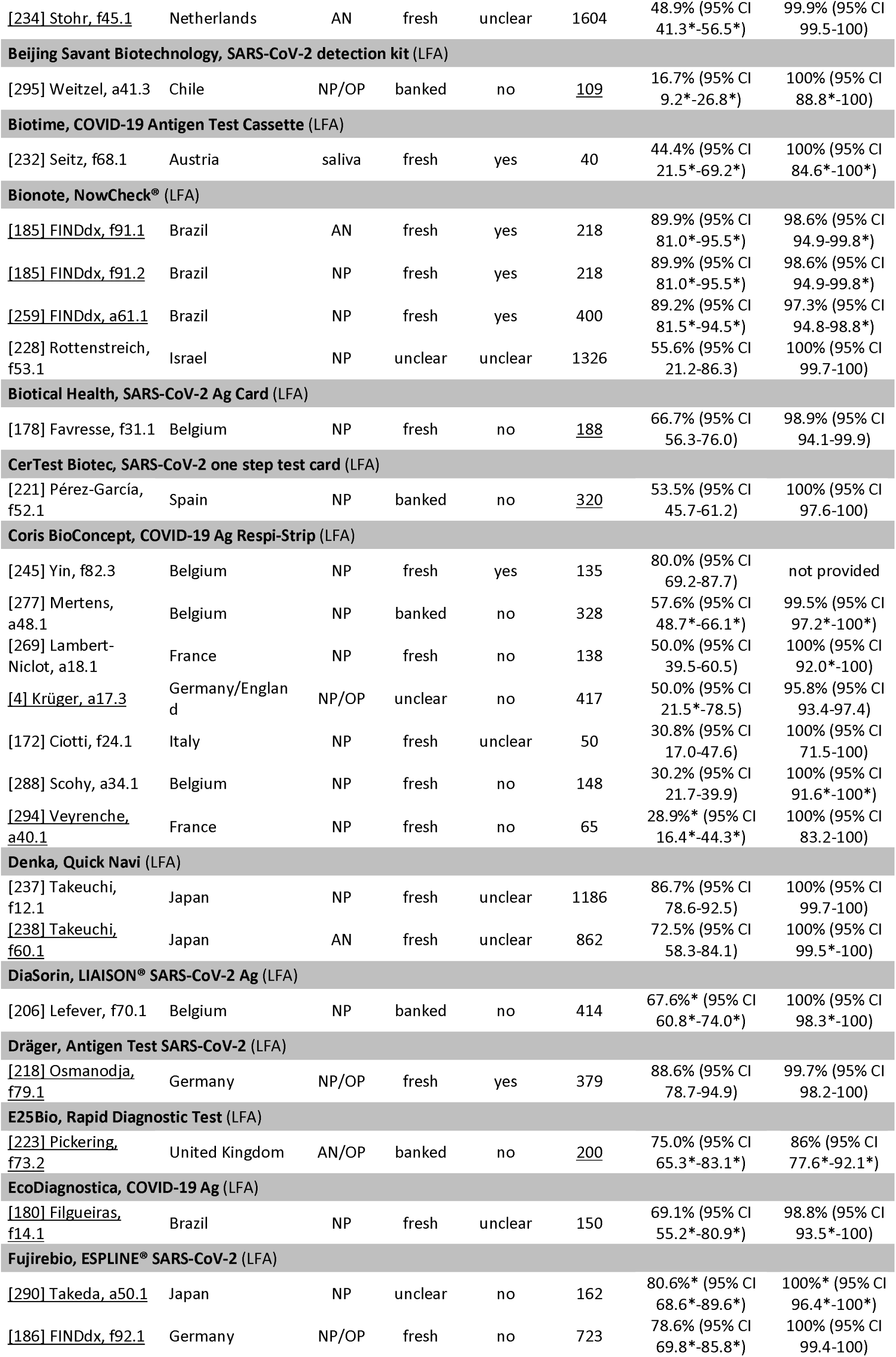
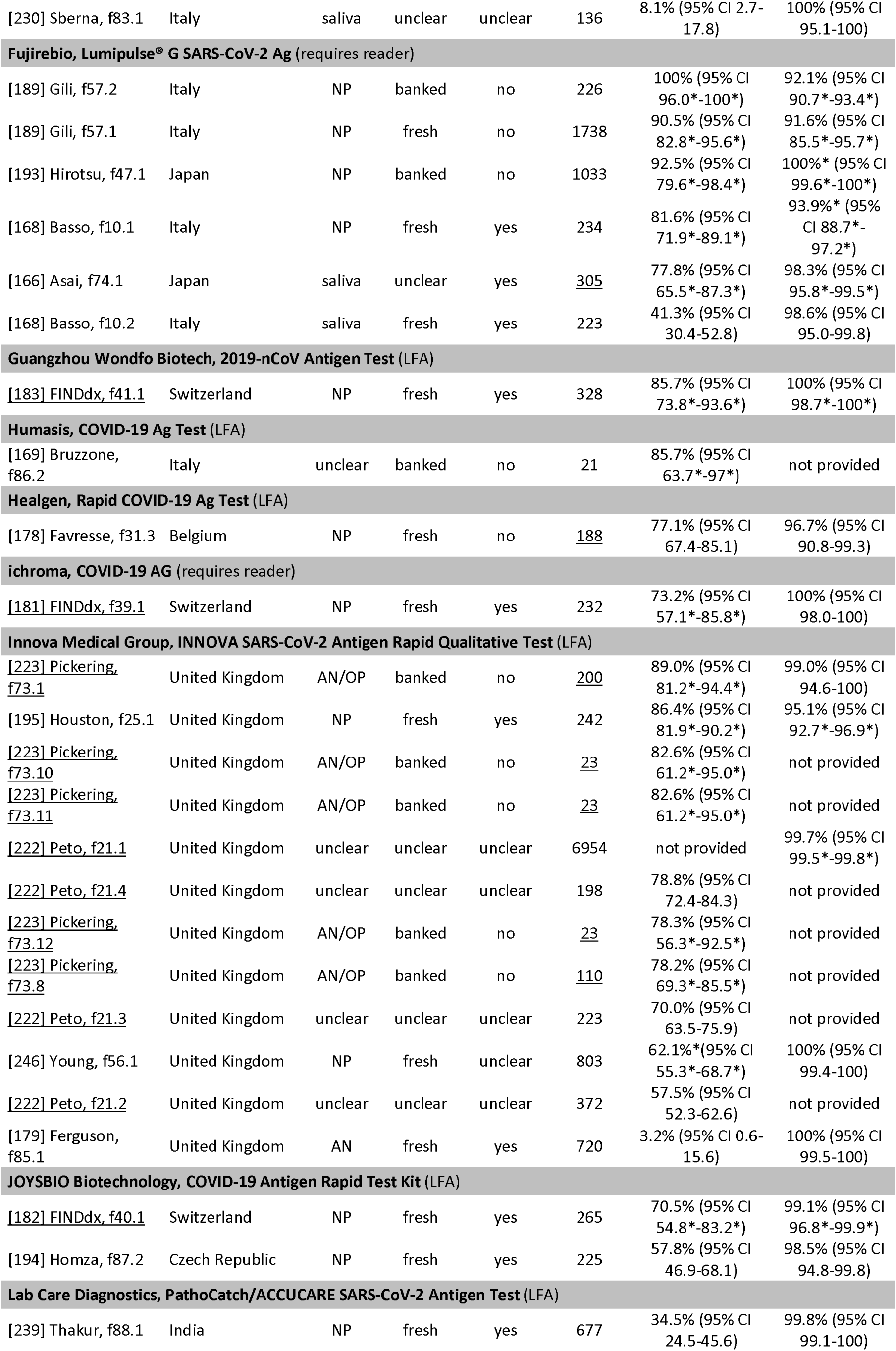
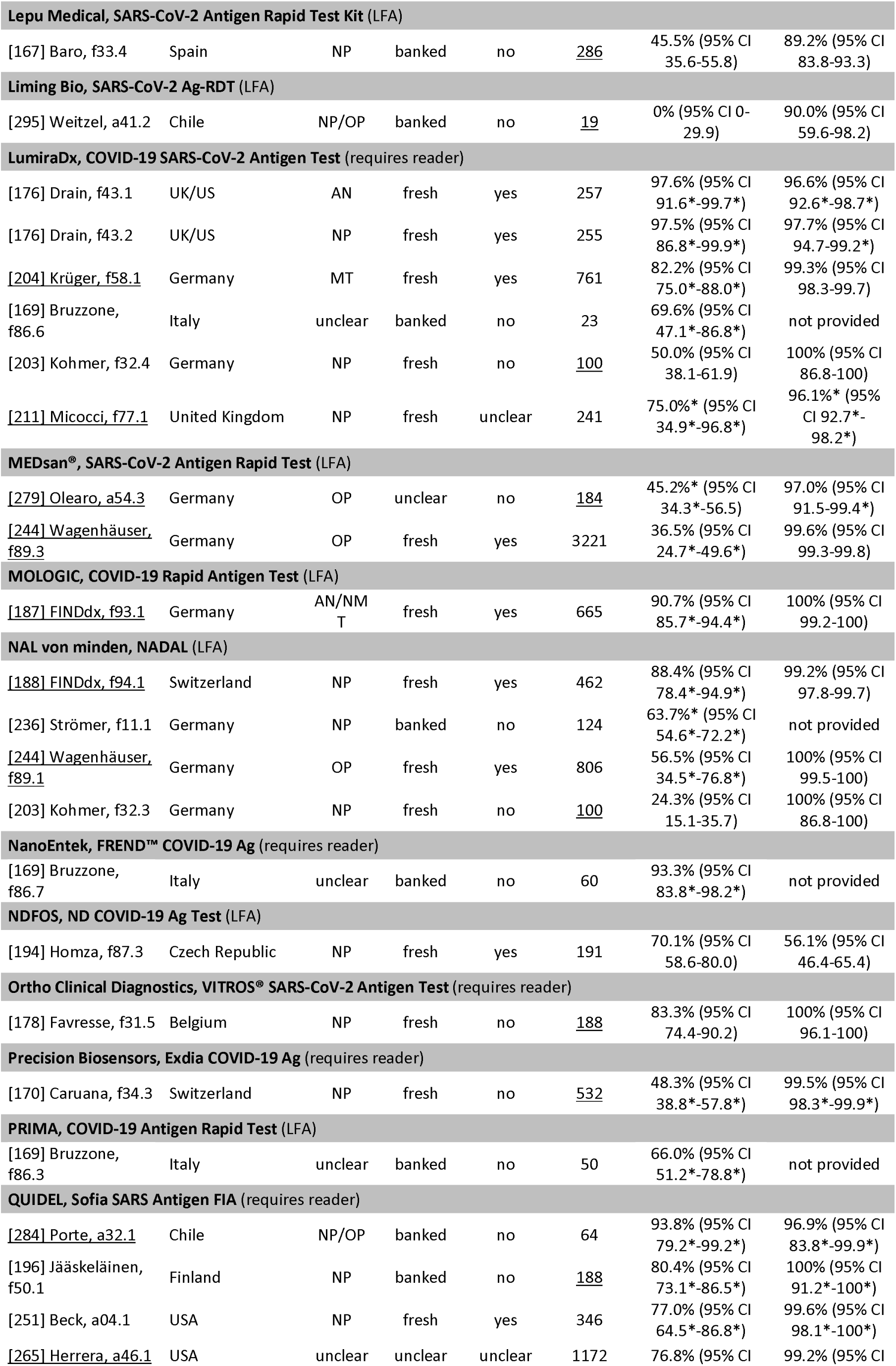
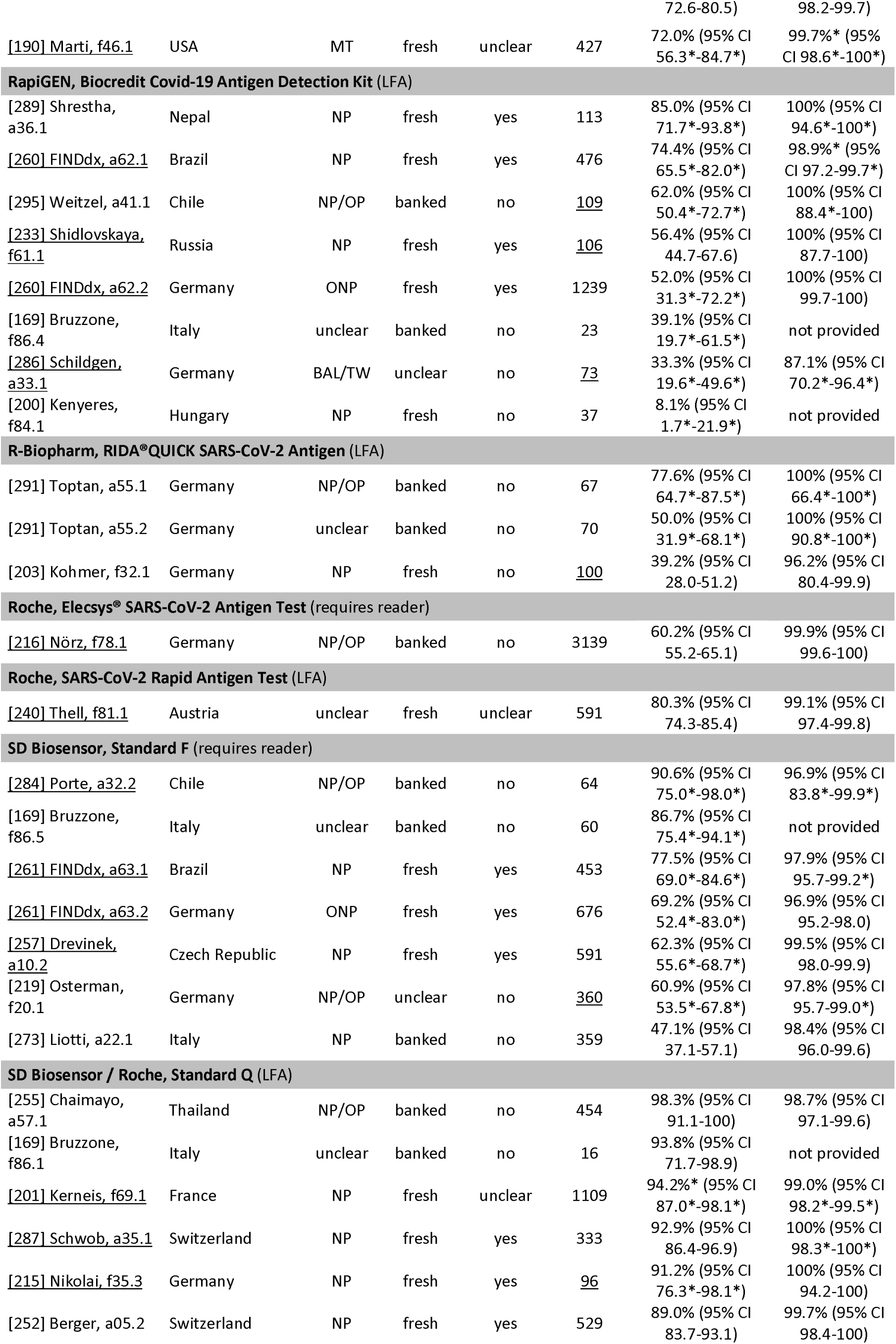
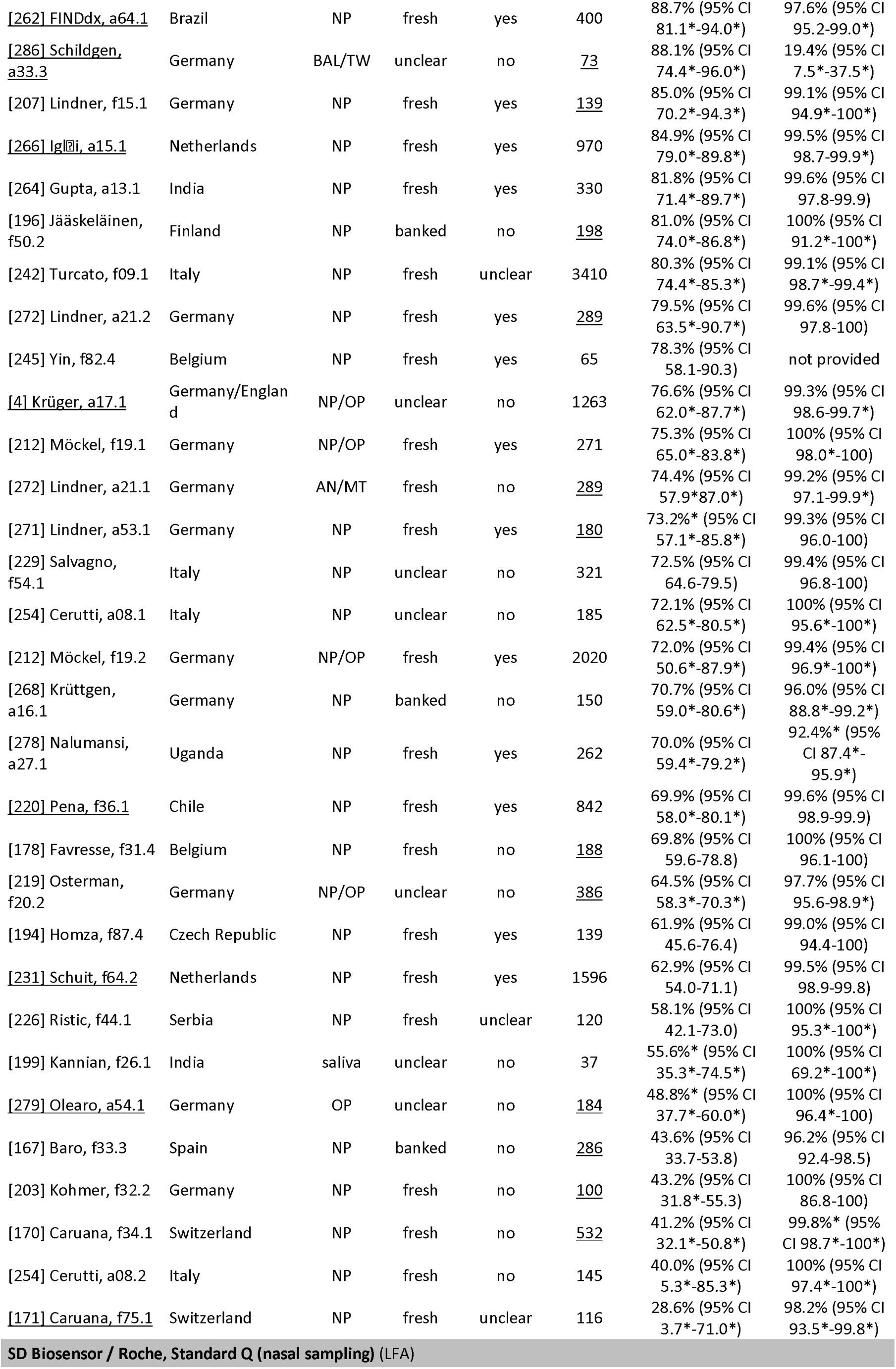
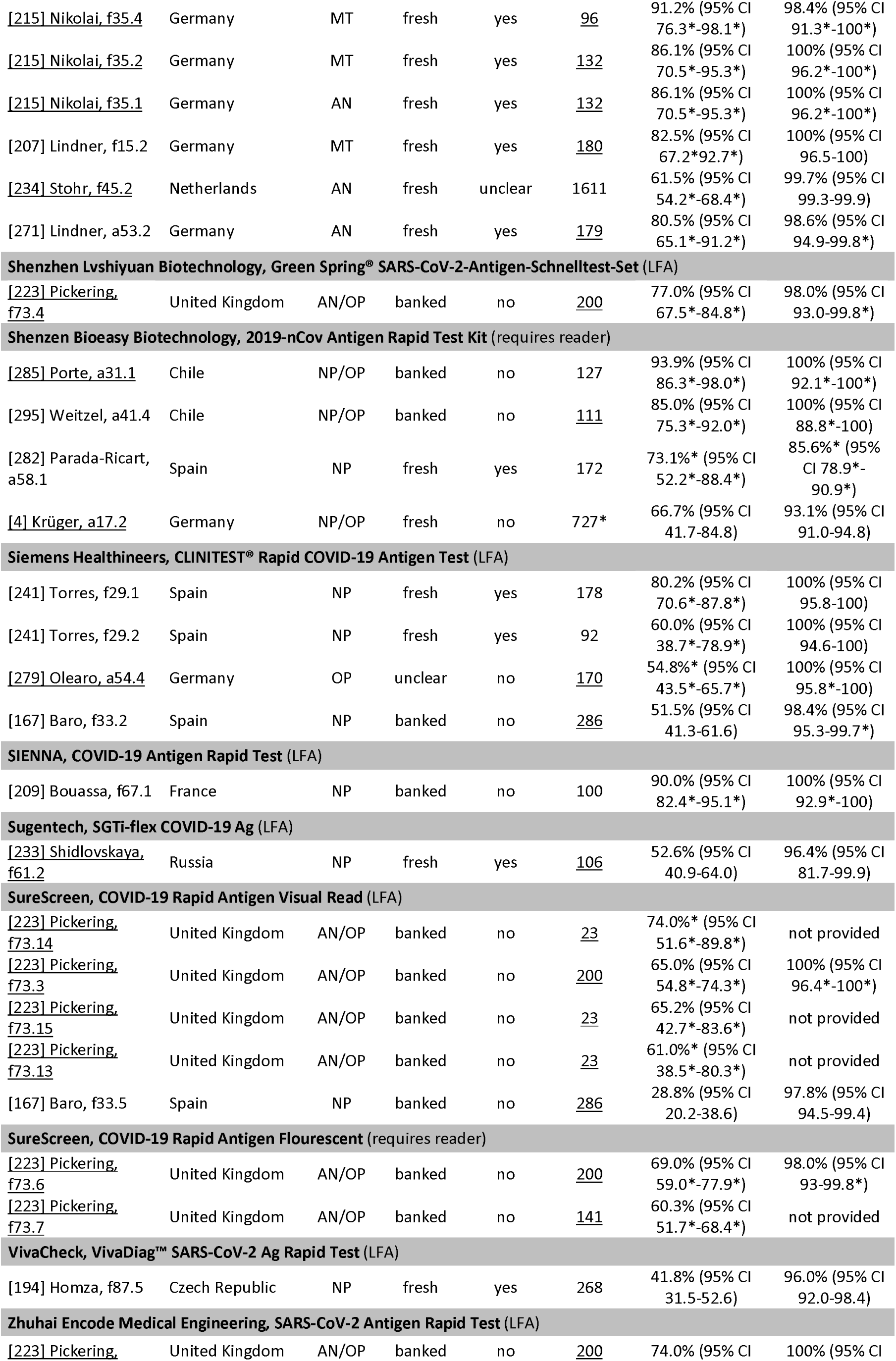
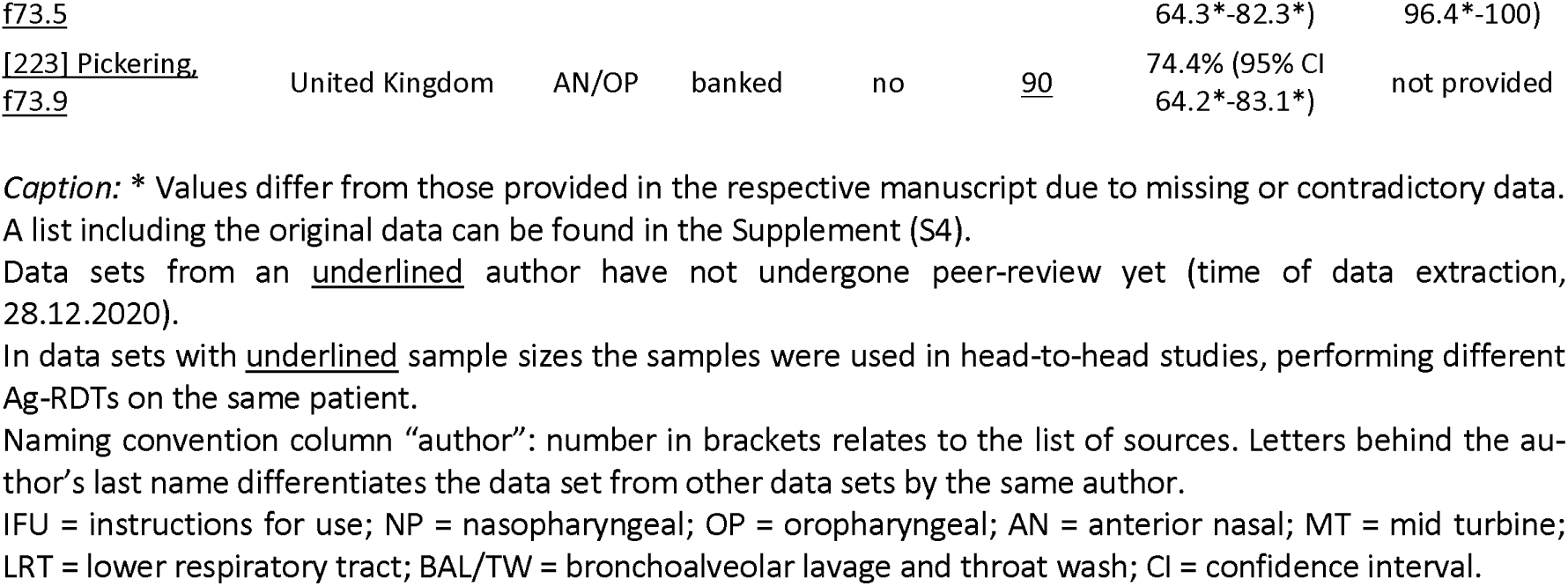
Clinical accuracy data for Ag-RDTs against SARS-CoV-2

### METHOLOGICAL QUALITY OF STUDIES

The findings on study quality using the QUADAS 2 tool are presented in Fig 2. In 190 (88.8%) data sets a relevant patient population was assessed. However, for only 44 (20.6%) of the data sets the patient selection was considered representative of the setting and population chosen (i.e., they avoided inappropriate exclusions, a case-control design and enrollment occurred consecutive or randomly).

**Figure 2.**
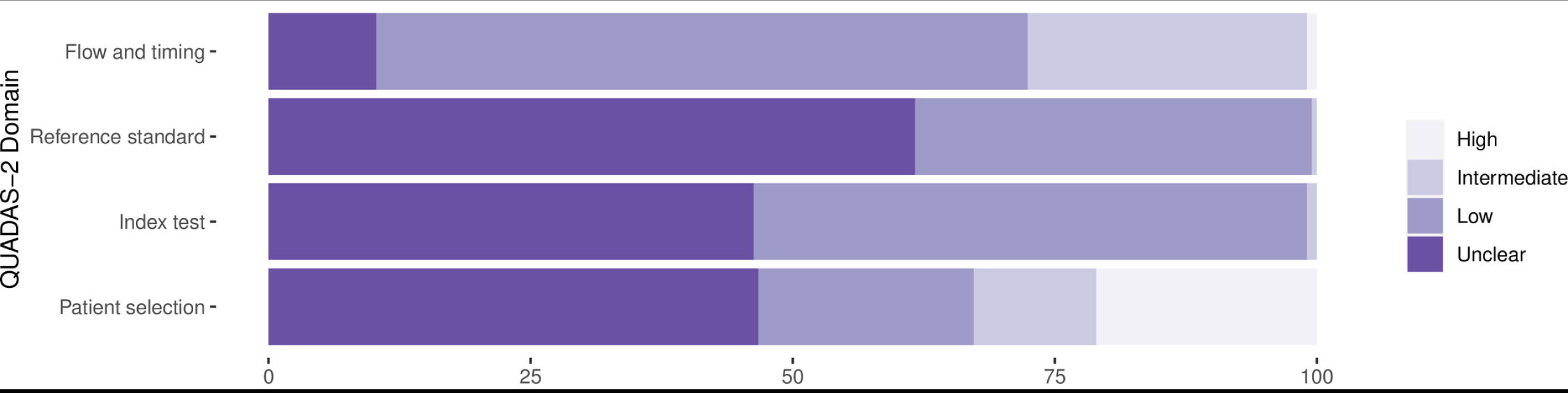
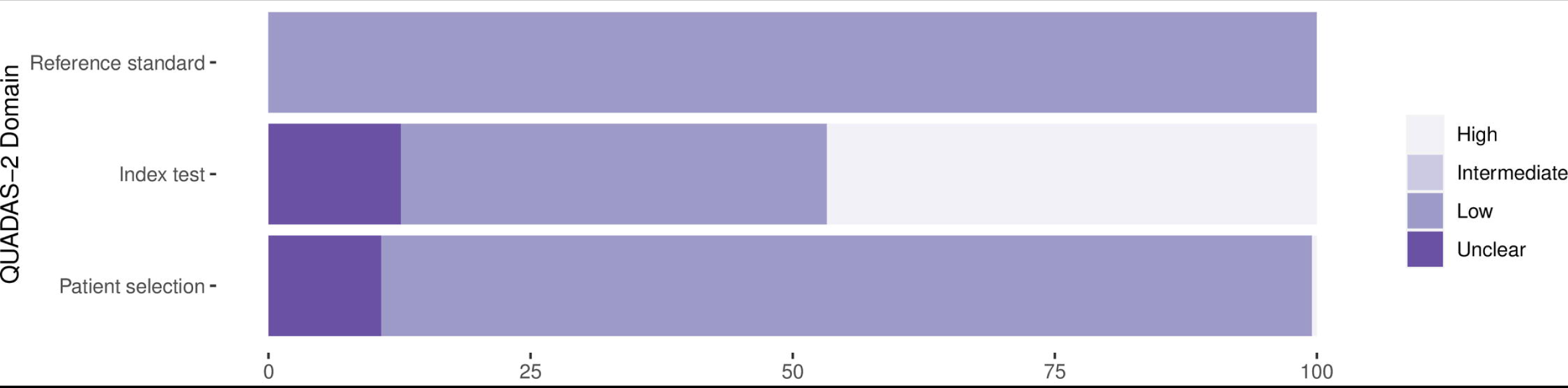

The conduct and interpretation of the index tests was considered to have low risk for introduction of bias in 113 (52.8%) of data sets (through e.g., appropriate blinding of persons interpreting the visual read-out). However, for 99 (46.3%) of data sets sufficient information to clearly judge the risk of bias was not provided. In only 89 (41.6%) of data sets the Ag-RDTs were performed according to IFU, while 100 (46.7%) were not IFU-conforming, potentially impacting the diagnostic accuracy (for 25 (11.7%) of data sets the IFU status was unclear).

In 81 (37.9%) of data sets the reference standard was performed before the Ag-RDT, or the operator conducting the reference standard was blinded to the Ag-RDT results, resulting in a low risk of bias. In almost all other data sets (132/61.7%) this risk could not be assessed due to missing data. The applicability of the reference test was judged to be of low concern for all data sets, as cell culture or RT-PCR are expected to adequately define the target condition.

In 209 (97.7%) data sets, the sample for the index test and reference test were obtained at the same time, while this was unclear in five (2.3%). All samples included in a data set were applied to the same type of RT-PCR in 145 (67.8%) data sets, while different types of RT-PCR were used within the same data set in 50 (23.4%) data sets. For 19 (8.9%), it was unclear. Furthermore, for 11 (5.1%) of data sets, there was a concern that not all selected patients were included in the analysis.

Finally, 32 (24.1%) of the studies received financial support from the Ag-RDT manufacturer and in another nine (6.8%) employment of the authors by the manufacturer of the Ag-RDT studied was indicated. Overall, a conflict of interest was found in 33 (24.8%) of the studies.

### DETECTION OF SARS-COV-2 INFECTION

Out of 214 clinical data sets (from 124 studies), 20 were excluded from the meta-analysis, as they included less than 20 RT-PCR positive samples. Further 21 data sets were missing either sensitivity or specificity and were only considered for univariate analyses. Across the remaining 173 data sets, including any test and type of sample, the pooled sensitivity and specificity were 71.2% (95%CI 68.2 to 74.0) and 98.9% (95%CI 98.6 to 99.1), respectively. If testing was performed in conformity with IFU, sensitivity increased to 76.3% (95%CI 73.1 to 79.2) compared to not IFU-conforming testing with a sensitivity of 65.9% (95%CI 60.6 to 70.8). Pooled specificity was similar in both groups (99.1% (95% CI 98.8-99.4)and 98.3% (95% CI 97.7 to 98.8), respectively).

### ANALYSIS OF SPECIFIC TESTS

Based on 119 data sets with 71,424 tests performed, we were able to perform bivariate meta-analysis of the sensitivity and specificity for twelve different Ag-RDTs (Fig 3A). Across these, pooled estimates of sensitivity and specificity on all samples were 72.1% (95%CI 68.8 to 75.3) and 99.0% (95% CI 98.7 to 99.2), which were very similar to the overall pooled estimate across all meta-analyzed data sets (71.2% and 98.9%, above).

**Figure 3a.**
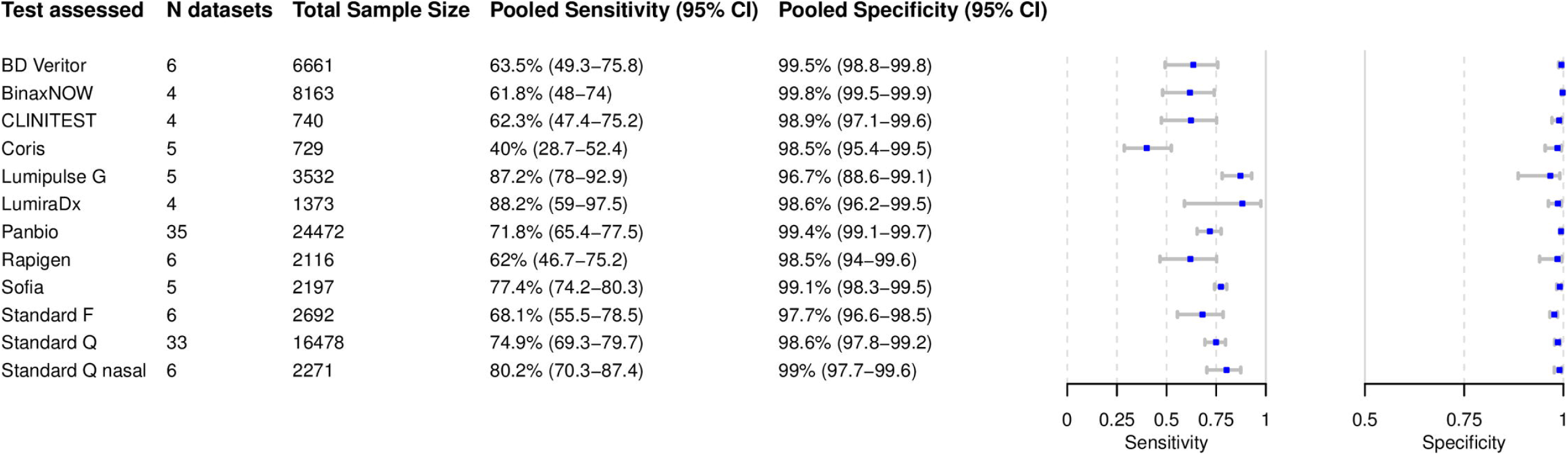

The highest pooled sensitivity was found for the SARS-CoV-2 Antigen Test by LumiraDx (United Kingdom; henceforth called LumiraDx) and the Lumipulse G SARS-CoV-2 Ag by Fujirebio (Japan; henceforth called Lumipulse G) with 88.2% (95% CI 59.0 to 97.5) and 87.2% (95% CI 78.0 to 92.9), respectively. The Sofia SARS Antigen FIA by QUIDEL (California, US; henceforth called Sofia) had a pooled sensitivity with 77.4% (95% CI 74.2 to 80.3). Of the non-instrument tests, the Standard Q and the Standard Q nasal test by SD Biosensor (South Korea; distributed in Europe by Roche, Germany; henceforth called Standard Q nasal) performed best with a pooled sensitivity of 74.9% (95% CI 69.3 to 79.7) and 80.2% (95% CI 70.3 to 87.4), respectively. The pooled sensitivity for Panbio was 71.8% (95%CI 65.4 to 77.5). From all Ag-RDTs, the COVID-19 Ag Respi-Strip by Coris BioConcept (Belgium; henceforth called Coris) had the lowest pooled sensitivity of 40.0% (95% CI 28.7 to 52.4).

The pooled specificity was above 98% for all of the tests, except for the Standard F by SD Biosensor (South Korea) and Lumipulse G with specificities of 97.7% (95% CI 96.6 to 98.5) and 96.7% (95% CI 88.6 to 99.1), respectively. Hierarchical summary receiver-operating characteristic for Standard Q and LumiraDx are available in the Supplement (S6).

Three Ag-RDTs did not have sufficient data to allow for a bivariate meta-analysis, wherefore a univariate analysis was conducted (Fig 3B). For the INNOVA SARS-CoV-2 Antigen Rapid Qualitative Test by Innova Medical Group (California, US) this resulted in a pooled sensitivity and specificity of 76.1% (95% CI 68.1 to 84.1) and 99.4% (95% CI 98.7 to 100), respectively. For the NADAL by NAL von Minden (Germany) and the COVID-19 Rapid Antigen Visual Read by SureScreen (United Kingdom), sufficient data was only available to analyze sensitivity, resulting in polled sensitivity estimates of 58.4% (95% CI 29.2 to 87.6) and 58.0% (95% CI 38.3 to 77.6) and, respectively.

**Figure 3b.**
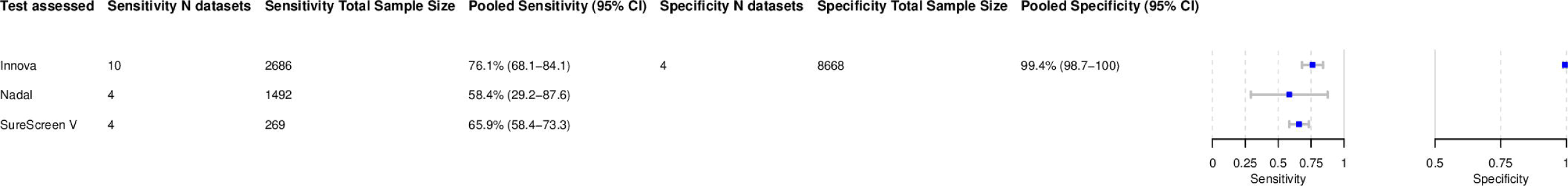

The remaining 35 Ag-RDTs did not present sufficient data for neither a uninor a bivariate meta-analysis. However, 9/35 have results presented in more than one data set and are summarized in Table 2. Herein, the widest ranges of sensitivity were found for the ESPLINE SARS-CoV-2 by Fujirebio (Japan) with sensitivity reported between 8.1% and 80.7%, and the RIDA®QUICK SARS-CoV-2 Antigen by R-Biopharm (Germany) with sensitivity between 39.2% and 77.6%, both with three data sets each. In contrast, two other test with two data sets each showed the least variability in sensitivity: the Zhuhai Encode Medical Engineering, SARS-CoV-2 Antigen Rapid Test (China) reported sensitivity between 74.0% and 74.4% and the COVID-19 Rapid Antigen Flourescent by SureScreen (UK) reported sensitivity between 60.3% and 69.0%. However, for both tests both data sets originated from the same studies. Overall, the lowest sensitivity range was reported for the SARS-CoV-2 Antigen Rapid Test by MEDsan (Germany) with 36.5% to 45.2% across two data sets. The specificity ranges were above 96% for most of the tests. A notable outlier was the 2019-nCov Antigen Rapid Test Kit by Shenzhen Bioeasy Biotechnology (China; henceforth called Bioeasy), reporting the worst with a specificity as low as 85.6% in one study. Forest plots for the data sets for each Ag-RDT are provided in the Supplement (S5). The remaining 26 Ag-RDTs that were evaluated in one data set only are included in Table 1 and Supplement (S5).

**Table 2:** Summary clinical accuracy data for major Ag-RDTs not included in the meta-analysis

| Manufacturer, Ag-RDT | Number of data sets | Sensitivity range | Specificity range | Comments |
| --- | --- | --- | --- | --- |
| Bionote, NowCheck® (LFA) | 3 | 55.6% to 89.9% | 97.3% to 100% | - Two of the studies were IFU-conform, whereas IFU-conformity for the study reporting 55.6% sensitivity was unclear |
| Denka, Quick Navi (LFA) | 2 | 72.5% to 86.7% | 100%* | - Both studies were conducted on fresh samples, but for the one reporting 72.5% IFU-conformity was unclear |
| Fujirebio, ESPLINE® SARS-CoV-2 (LFA) | 3 | 8.1% to 80.7% | 100%* | - The data set reporting 8.1% sensitivity used saliva samples (not IFU-conform) and the majority of samples showed a Ct-value > 25 |
| JOYSBIO Biotechnology, COVID-19 Antigen Rapid Test Kit (LFA) | 2 | 57.8% to 70.5% | 98.5% to 99.1% | - The datasets used NP and AN samples, respectively. Both were performed by IFU on symptomatic people or high-risk contacts |
| MEDsan®, SARS-CoV-2 Antigen Rapid Test (LFA) | 2 | 36.5% to 45.2% | 97% to 99.6% | - Both studies were conducted on OP samples, which is IFU-conforming for this test |
| R-Biopharm, RIDA®QUICK SARS-CoV-2 Antigen (lateral flow assay) | 3 | 39.2% to 77.6% | 96.2% to 100% | - Two data sets originate from the same study and no study was conducted as per IFU<br>- The data set reporting 39.2% included only asymptomatic persons with Ct-values between 22.1 and 36.4 |
| Shenzen Bioeasy Biotechnology, 2019-nCov Antigen Rapid Test Kit (requires reader) | 4 | 66.7% to 93.9% | 85.6% to 100% | - The data set reporting 85.6% specificity was conducted IFU-conforming<br>- The data sets reporting highest sensitivity were drawn from just symptomatic patients, for the others symptomatic patients made up more than two thirds of the study population |
| SureScreen, COVID-19 Rapid Antigen Fluorescent (requires reader) | 2 | 60.3% to 69.0% | 98%* | - Both datasets originate from the same study and were conducted not IFU-conforming on stored samples |
| Zhuhai Encode Medical Engineering, SARS-CoV-2 Antigen Rapid Test (LFA) | 2 | 74.0% to 74.4% | 100%* | - Both datasets originate from the same study, a retrospective head-to-head comparison<br>- Stored AN/MT samples were assessed |

*Caption:* \* only one data set for specificity was provided
IFU = instructions for use; Ag-RDT = antigen rapid diagnostic test; NP = nasopharyngeal; OP = oropharyngeal;
AN = anterior nasal; MT = mid-turbinate.

In total, 16 studies accounting for 53 data sets conducted head-to-head clinical accuracy evaluations of different tests using the same sample(s) from the same participant. These data sets are underlined in Table 1; 15 such studies included more than 100 samples, and one study included too few samples to draw clear conclusions [286]. Four studies performed their head-to-head evaluation as per manufacturers’ instructions and on symptomatic patients. Across three of them, the Standard Q (sensitivity 73.2% to 91.2%) and the Standard Q nasal (sensitivity 82.5% to 91.2%) showed a similar range of sensitivity [207,215,271]. The fourth reported a sensitivity of 56.4% (95% CI 44.7 to 67.6) for the Biocredit Covid-19 Antigen rapid test kit by RapiGEN (South Korea; henceforth called Rapigen) and 52.6% (95% CI 40.9 to 64) for the SGTi-flex COVID-19 Ag by Sugentech (South Korea) [233].

All other head-to-head comparisons were not IFU-conforming. In one of these, the Rapid COVID-19 Ag Test by Healgen (sensitivity 77.1%) performed better than the Standard Q and Panbio (sensitivity 69.8% and 67.7%, respectively) [178]. In contrast to the overall findings of the meta-analysis above, two other head-to-head studies found that both the Standard Q (sensitivity 43.6% and 49.4) and Panbio (sensitivity 38.6% and 44.6%) had lower performance than the CLINITEST® Rapid COVID-19 Antigen Test by Siemens Healthineers (Germany; henceforth called Clinitest), which reported sensitivities 51.5% and 54.9% [167, 279]. However, another study found both the Standard Q and Panbio (sensitivity 81.0% and 82.9%, respectively) to have a higher accuracy than the Sofia (sensitivity 80.4%) [196].

### SUBGROUP ANALYSIS

The results are presented in Fig 4. Detailed results for the subgroup analysis are available in the Supplement (S7 to 11).

#### Subgroup analysis by Ct-values

High sensitivity was achieved for Ct-values <20 with 96.5% (95% CI 92.6 to 98.4). The pooled sensitivity for Ct-values <25 was markedly better at 95.8% (95% CI 92.3 to 97.8) compared to the group with Ct ≥ 25 at 50.7% (95% CI 35.6 to 65.8). A similar pattern was observed when the Ct-values were analyzed using cut-offs <30 or ≥30, resulting in a sensitivity of 79.9% (95% CI 70.3 to 86.9) and 20.9% (95% CI 12.5 to 32.8), respectively (Fig 4A).

**Figure 4a.**
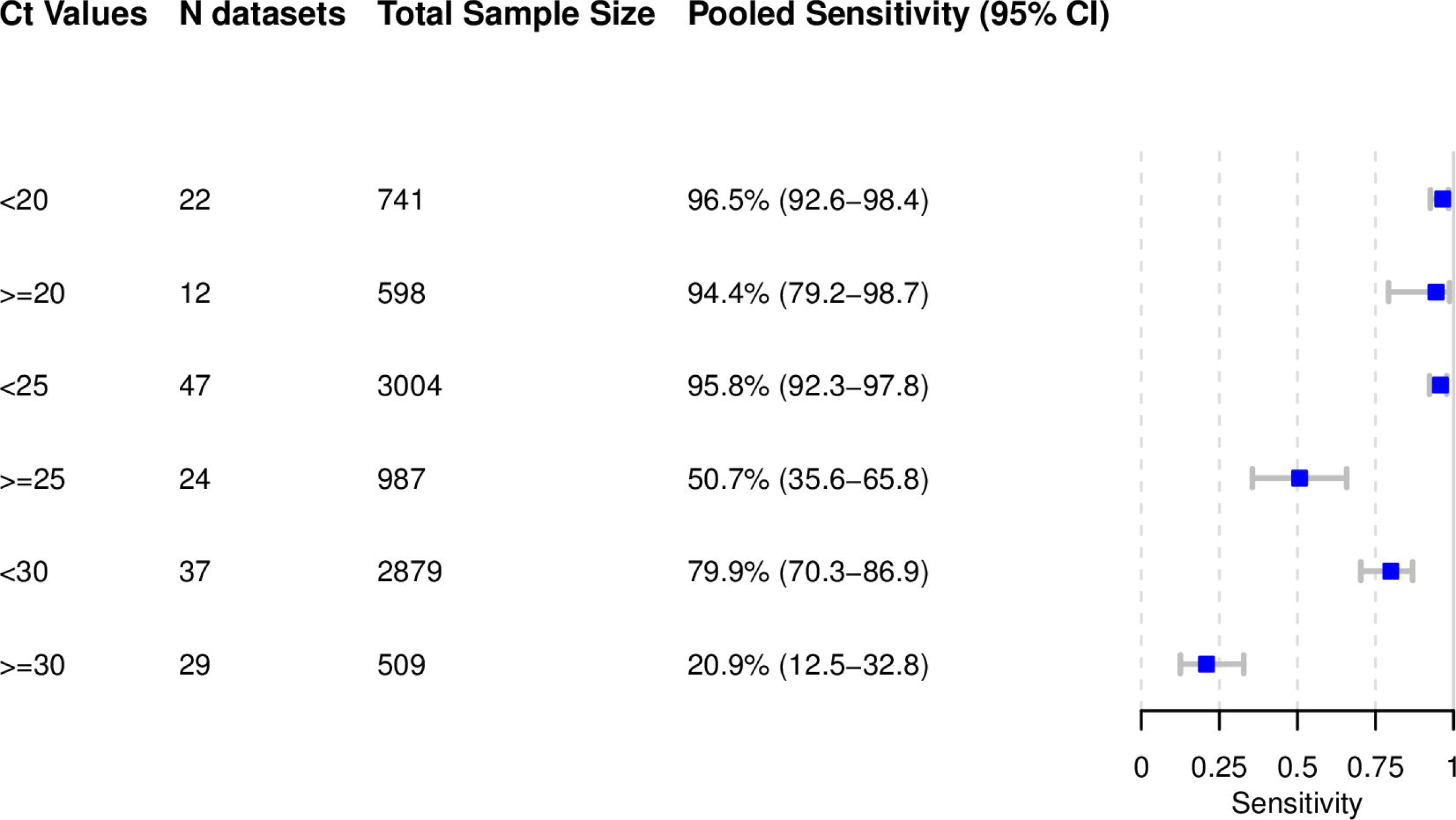

In addition, it was possible to meta-analyze test specific pooled sensitivity for the Panbio: 97.7% sensitivity (95% CI 95.3 to 98.9) for Ct-value <20, 95.8% (95% CI 92.3 to 97.8) for Ct-value <25 and 83.4% (95% CI 69.1 to 91.9) for Ct-value <30. For Ct-value ≥25 sensitivity was 61.2% (95% CI 38.8 to 79.7) and 30.5% (95% CI 16.0 to 50.4) for Ct-value ≥30. For the other Ag-RDTs only limited data was available, which is presented in the supplements (S10).

#### Subgroup analysis by IFU conformity

The summary results are presented in Fig 4B. When assessing only studies with an IFU-conforming testing, pooled sensitivity from 81 datasets with 49,643 samples was 76.3% (95% CI 73.1 to 79.2). When not IFU-conforming sampling (75 data sets, 31,416 samples) was performed sensitivity decreased to 65.9% (95% CI 60.6 to 70.8).

**Figure 4b.**
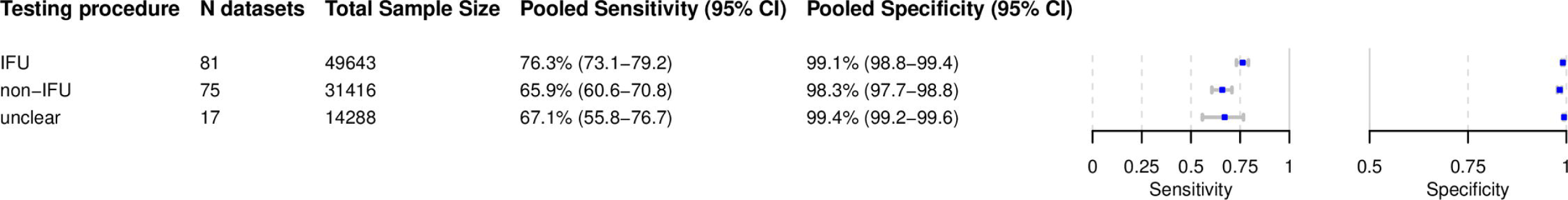

For five tests it was possible to calculate pooled sensitivity estimates only including data sets with an IFU-conforming testing: Panbio (sensitivity of 76.5% (95% CI 69.5 to 82.3); 17 data sets, 12,856 samples), Standard Q (sensitivity of 79.3% (95% CI 73.5 to 84.1); 15 data sets, 6,584 samples), BinaxNOW (sensitivity of 61.8% (95% CI 48.0 to 74.0); 4 data sets, 8,163 samples), Rapigen (sensitivity of 67.1% (95% CI 50.4 to 80.4); 4 data sets, 1,934 samples) and Standard Q nasal (sensitivity of 83.8% (95% CI 77.8 to 88.4); 5 data sets, 683 samples). Specificity was above 98.6% for all tests.

In contrast, when the Panbio (14 data sets, 9,233 samples) and Standard Q (14 data sets, 4,714 samples) tests were not performed according to IFU, pooled sensitivity decreased to 64.3% (95% CI 50.9 to 75.8) and 67.4 (95% CI 57.2 to 76.2), respectively.

#### Subgroup analysis by sample type

Most data sets evaluated NP or combined NP/OP swabs (122 data sets and 59,810 samples) as the sample type for the Ag-RDT. NP or combined NP/OP swabs achieved a pooled sensitivity of 71.6% (95% CI 68.1 to 74.9). Data sets that used AN/MT swabs for Ag-RDTs (32 data sets and 25,814 samples) showed a summary estimate for sensitivity of 75.5% (95% CI 70.4 to 79.9). This was confirmed by two studies that reported direct head-to-head comparison of NP and MT samples from the same participants using the same Ag-RDT (Standard Q), where the two sample types showed equivalent performance [271, 272]. Analysis of performance with an OP swab (seven data sets, 5,165 samples), showed pooled sensitivity of only 53.1% (95%CI 40.9 to 65.0). Saliva swabs (four data sets, 1,088 samples) showed the lowest pooled sensitivity with only 37.9% (95% CI 11.8 to 73.5) (Fig 4C).

**Figure 4c.**
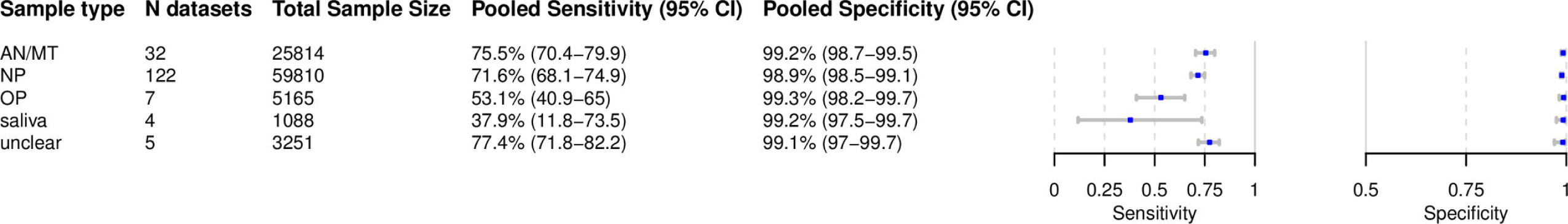

We were not able to perform a subgroup meta-analysis for BAL/TW due to insufficient data as there was only one study with 73 samples evaluating the Rapigen, Panbio and Standard Q [286]. However, BAL/TW would in any case be considered an off-label use.

#### Subgroup analysis in symptomatic and asymptomatic patients

Within the data sets possible to meta-analyze, 17,964 (54.1%) samples were from symptomatic and 15,228 (45.9%) from asymptomatic patients. The pooled sensitivity for symptomatic patients was markedly different compared to asymptomatic patients with 76.7% (95% CI 70.6 to 81.9) vs. 52.5% (95% CI 43.7 to 61.1). Specificity was 99% for both groups (Figu 4D). Median Ct-values differed in symptomatic and asymptomatic patients. For those studies where it was possible to extract a median Ct-value, it ranged from 20.5 to 27.0 in symptomatic [170,207,226,258,271,272] and from 27.2 to 30.5 in asymptomatic [170,201,258] patients.

#### Subgroup analysis comparing symptom duration

Data were analyzed for 5,538 patients with symptoms less than 7 days, but very limited data were available for patients with symptoms ≥ days (397 patients). The pooled sensitivity for patients with onset of symptoms <7 days was 83.8% (95% CI 76.3 to 89.2) which is markedly higher than the 61.5% (95% CI 52.2 to 70.0) sensitivity for individuals tested ≥ 7 days from onset of symptoms (Fig 4D).

**Figure 4d.**
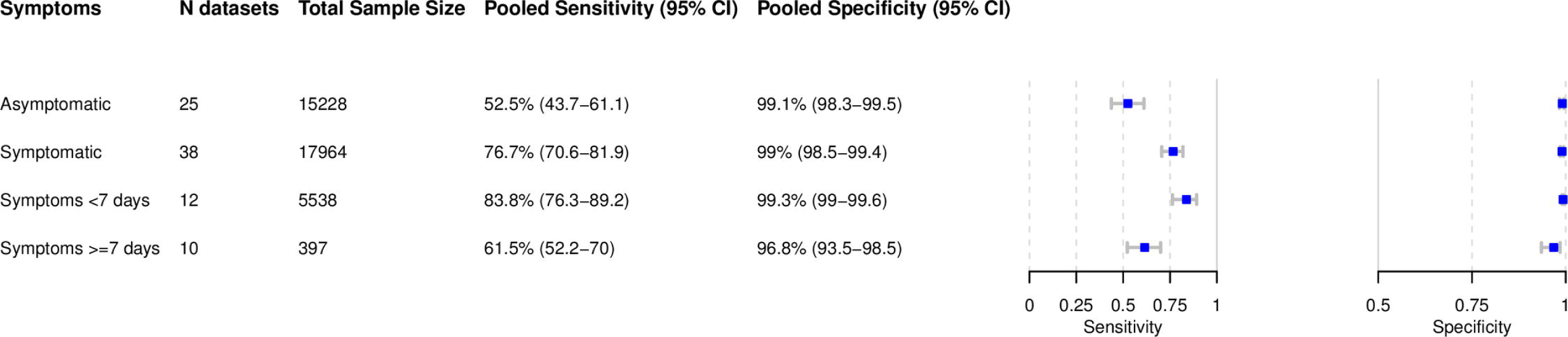

#### Subgroup analysis by age

For adult patients, it was possible to pool estimates across 3,837 samples, whereas the pediatric group included 7,326 samples. Sensitivity and specificity were 64.3% (95% CI 54.7 to 72.9) and 99.4% (95% CI 98.9 to 99.7) in mostly symptomatic patients <18 years. In patients ≥ sensitivity increased to 74.8% (95% CI 66.5 to 81.6), while the specificity was similar (98.7%, 95% CI 97.2 to 99.4) (Fig 4E).

**Figure 4e.**
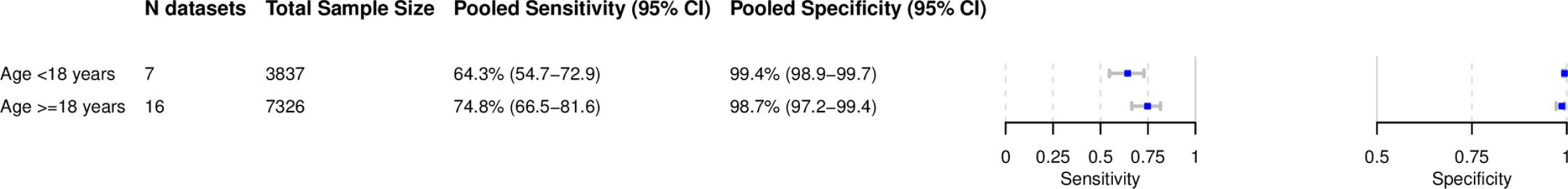

#### Subgroup analysis by type of RT-PCR and viral load

We were not able to perform a meta-analysis for the subgroups by type of RT-PCR or viral load (viral copies/mL) due to insufficient data.

In 152 (71.0%) of the data sets only one type of RT-PCR was used, whereas 37 (17.3%) tested samples in the same data set using different RT-PCRs. For 25 (11.7%) of the data sets the type of RT-PCR was not reported. The Cobas® SARS-CoV-2 Test from Roche (Germany) was used most frequently in 63 (29.4%) of the data sets, followed by the Allplex® 2019 n-CoV Assay from Seegene in 41 (19.2%) and the SARS CoV-2 assay from TaqPath in 20 (9.3%) of the data sets

Median sensitivity in samples with viral load of >5 log 10 copies/ml was 72.4% (range 46.9 to 100), 97.8% (range 71.4 to 100) for >6 log 10 copies/ml and 100% (range 93.8 to 100) for >7 log 10 copies/ml, showing that the sensitivity increases with increasing viral load.

#### Meta regression

We were not able to perform a meta-regression due to the considerable heterogeneity in reporting sub-groups, which resulted in too few studies with sufficient data for comparison.

#### Publication Bias

The result of the Deeks’ test (p=0.001) shows significant asymmetry in the funnel plot for all datasets with complete results. This indicates there may be publication bias from studies with small sample sizes. However, when looking at publications specifically for LumiraDx (p=0.567), Standard Q (p=0.23), and Panbio (p=0.35), the results do not show significant asymmetry in the funnel plots, indicating these tests may have less publication bias. All funnel plots are listed in the Supplement (S12).

### COMPARISON WITH ANALYTICAL STUDIES

The nine analytical studies were divided into 63 data sets, evaluating 23 different Ag-RDTs. Only seven studies reported a samples size, wherein 833 (90.6%) samples originated from NP swabs and for 86 (9.4%) the sample type was unclear. One of the two studies not reporting sample size used saliva samples [198], while the other used the sample type specified in the respective Ag-RDT’s IFU [173].

Overall, the reported analytical sensitivity (limit of detection) in the studies resembled the results of the meta-analysis presented above. Rapigen (limit of detection (LOD) in log10 copies per swab: 10.2) and Coris (LOD 7.46) were found to perform worse than Panbio (LOD 6.6 to 6.1) and Standard Q (LOD 6.8 to 6.0), whereas the Clinitest (LOD 6.0) and the BinaxNOW by Abbott (LOD 4.6 to 4.9) performed even better [191,256,282]. Similar results were found in another study, where the Standard Q showed the lowest LOD (detecting virus up to what is an equivalent Ct-value of 26.3 to 28.7), when compared to that of Rapigen and Coris (detecting virus up to what is an equivalent Ct-value of only 18.4 for both) [208,274,275]. However, another study found the Panbio, Standard Q, Coris and BinaxNOW to have a similar LOD of 5.0*10^3^ plaque forming units (pfu) /milliliter (ml), but the ESPLINE SARS-CoV-2 by Fujirebio (Japan), the COVID-19 Rapid Antigen Test by MOLOGIC (United Kingdom) and the Sure Status COVID-19 Antigen Card Test by Premier Medical Corporation (India) to perform markedly better (LOD 2.5 to 5.0*10^2^ pfu/ml) [173]. An overview of all LODs reported in the studies can be found in the Supplement (S13)

### SENSITIVITY ANALYSIS

When the data sets from case control studies (25/173) were excluded, the estimated sensitivity did not differ greatly, with a value of 70.9% (95% CI 67.7 to 73.9) compared to 71.2% (95% CI 68.2 to 74.0) in the overall analysis with no change in pooled specificity. When excluding the data sets from pre-prints (64/173), sensitivity decreased slightly to 67.2% (95% CI 62.9 to 71.3) compared to the overall analysis.

## DISCUSSION

In this comprehensive systematic review and meta-analysis, we have summarized the data of 133 studies evaluating the accuracy of 61 different Ag-RDTs. Across all meta-analyzed samples, our results show a pooled sensitivity and specificity of 71.2% (95% CI 68.2 to 74.0) and 98.9% (95% CI 98.6 to 99.1). Over half of the studies did not perform the Ag-RDT in accordance with the test manufacturers’ recommendation or the performance was unknown, which negatively impacted the sensitivity. When considering only IFU-conforming studies, the sensitivity increased to 76.3% (95% CI 73.1 to 79.2). While we found the sensitivity to vary across specific tests, the specificity was consistently high.

The two Ag-RDTs that have been approved through the WHO emergency use listing procedure, Abbott Panbio and SD Biosensor Standard Q (distributed by Roche in Europe), have not only drawn the largest research interest, but also perform at or above average when comparing their pooled accuracy to that of all Ag-RDTs (sensitivity of 71.8% for Panbio and of 74.9% for Standard Q). The Standard Q nasal demonstrated an even higher pooled sensitivity (80.2% compared to the NP test), although this is likely due to variability in populations tested, as head-to-head performance showed a comparable sensitivity. Three other Ag-RDTs showed an even higher accuracy with sensitivities ranging from 77.4% to 88.2% (namely Sofia, Lumipulse G and LumiraDX), but were only assessed on relatively small samples sizes (ranging from 1,373 to 3,532) and all required an instrument/reader.

Not surprisingly, lower Ct-values, the RT-PCR semi-quantitative correlate for a high virus concentration, resulted in a significantly higher Ag-RDT sensitivity than higher Ct-values (pooled sensitivity 96.5% and 95.8% for ct-values <20 and <25 vs. 50.7% and 20.9% for ct-values ≥25 and ≥30). This confirms prior data that suggested that antigen concentrations and Ct-values were highly correlated in NP samples [16]. Ag-RDTs also showed a higher sensitivity in patients within 7 days after symptom onset compared to patients later in the course of the disease (pooled sensitivity 83.8% vs. 61.5%), which is to be expected given that samples from patients within the first week after symptom onset have been shown to contain the highest virus concentrations [297]. In line with this, studies reporting an unexpectedly low overall sensitivity either shared a small population size with an on average high Ct-value [230,273,288] or performed the Ag-RDT not as per IFU, e.g., using saliva or prediluted samples [167,170,203,248,279]. In contrast, studies with an unusually high Ag-RDT sensitivity were based on study populations with a low median Ct-value, between 18 and 22 [189,255,284].

Our analysis also found that the accuracy of Ag-RDTs is substantially higher in symptomatic patients than in asymptomatic (pooled sensitivity 76.7% vs. 52.5%). This is not surprising as studies that enrolled symptomatic patients showed a lower range of median Ct-values (i.e., higher viral load), than studies enrolling asymptomatic patients. Given that other studies found symptomatic and asymptomatic patients to have comparable viral loads [298, 299], the differences found in our analysis are likely explained by the varied time in the course of the disease at which testing is performed in asymptomatic patients presenting for one-time screening testing. As symptoms start in the early phase of the disease when viral load is still high, studies testing only symptomatic patients have a higher chance of including patients with high viral loads. In contrast, study populations drawn from asymptomatic patients only have a higher chance of including patients at any point of disease (i.e., including late in disease, when PCR is still positive, but viable virus is rapidly decreasing) [300].

With regards to the sampling and testing procedure, we found Ag-RDTs to perform similarly across upper-respiratory swab samples (e.g., NP and AN/MT), particularly when considering the most reliable comparisons from head-to-head studies.

Similar to previous assessment [7], the methodological quality of the included studies revealed a very heterogenous picture. In the future, aligning the design of clinical accuracy studies to common agreed upon minimal specifications (e.g., by WHO or European Center of Disease Control) and reporting the results in a standardized way [301] would improve data quality and comparability.

The main strengths of our study lie in its comprehensive approach and continuous updates. By linking this review to our website www.diagnosticsglobalhealth.org, we strive to equip decision makers with the latest research findings on Ag-RDTs for SARS-CoV-2 and, to the best of our knowledge, are the first in doing so. At least once per week the website is updated by continuing the literature search and process described above. We plan to update the meta-analysis on a monthly basis and publish it on the website. Furthermore, our study used rigorous methods as both the study selection and data extraction were performed by one author and independently validated by a second, we conducted blinded pilot extractions before of the actual data extraction, and we prepared a detailed interpretation guide for the QUADAS-2 tool.

The study may be limited by the inclusion of both preprints and peer-reviewed literature, which could affect the quality of the data extracted. However, we aimed to balance this potential effect by applying a thorough assessment of all clinical studies included, utilizing the QUADAS-2 tool and performing a sensitivity analysis excluding preprint manuscripts. In addition, the studies included in our analysis varied widely in the reported range of viral loads, limiting the comparability of their results. To control for this, we analyzed the Ag-RDTs’ performance at different levels of viral load. Finally, even though we are aware that further data exits from other sources, for example from governmental research institutes exists [302], such data could not be included as sufficient detail describing the methods and results are not publicly available.

## CONCLUSION

In summary, it can be concluded that there are Ag-RDTs available that have high sensitivity, particularly when performed in the first week of illness when viral load is high, and excellent specificity. However, our analysis also highlights the variability in results between tests (which is not reflected in the manufacturer reported data), indicating the need for independent validations. Furthermore, the analysis highlights the importance of performing tests in accordance with the manufacturers’ recommended procedures, and in alignment with standard diagnostic evaluation and reporting guidelines. The accuracy achievable by the best-performing Ag-RDTs, combined with the rapid turn-around time compared to RT-PCR, suggests that these tests could have a significant impact on the pandemic if applied in thoughtful testing and screening strategies.

## Data Availability

All data is available upon request.

https://zenodo.org/record/4924035#.YMSDcS2230p

## ABBREVIATIONS

Ag-RDT: = antigen rapid diagnostic test
AN/MT: = anterior nasal or mid-turbinate
BAL/TW: = bronchoalveolar lavage or throat wash
CI: = confidence interval
Ct-value: = cycle threshold value
FIND: = Foundation for Innovative New Diagnostics
FP: = false positive
FN: = false negative
IFU: = instructions for use
LRT: = lower respiratory tract
ML: = milliliter
N: = sample size
NP: = nasopharyngeal
OP: = oropharyngeal
PFU: = plaque forming units
RT-PCR: = reverse transcriptase polymerase chain reaction
TP: = true positive
TN: = true negative
VTM/UT: = viral or universal transport medium

## Notes

### Competing Interest Statement

The authors have declared no competing interest.

### Funding Statement

The study was supported by the Ministry of Science, Research and Arts of the State of Baden-Wuerttemberg, Germany and internal funds from the Heidelberg University Hospital as well as grants from UK Department of International Development (DFID, recently replaced by FCMO), grants from World Health Organization (WHO), grants from Unitaid to Foundation of New Diagnostics (FIND). The corresponding author had access to all data at all time.

### Author Declarations

All relevant ethical guidelines have been followed. There were no IRB or ethics committee approvals required.

### Summary of Updates

This revision includes an update of the meta analysis up until 30th of April and a major revision from the respective journal

