## Supplement File for "The accuracy of novel antigen rapid diagnostics for SARS-CoV-2: a living systematic review and meta-analysis"

Supplementary File

#### S1 PRISMA Checklists

PRISMA Checklist 2020 and PRISMA Abstract Checklist 2020 are submitted as a separate file.

#### S2 Search strategy

### Review questions

To assess the accuracy and ease-of-use of marketable antigen point of care diagnostics for SARS-CoV-2 compared to RT-PCR based on manufacturer independent evaluations.

Restriction: start Dezember 2019

### Definitions

P

SARS-CoV-2

I

Antigen nachweisenden Schnelltests Ag RDT

### Strategie

| 1 | P |
| --- | --- |
| 2 | I |
| 3 | 1 AND 2 |

### Searched Databases

- PubMed
- Web of Science Core Collection
- BioRxiv
- MedRxiv

### PubMed

P

(**"Severe Acute Respiratory Syndrome Coronavirus 2"[Supplementary Concept] OR**

**"COVID-19" [Supplementary Concept] OR**

**"Betacoronavirus"[Mesh] OR**

**"Coronavirus"[Mesh] OR**

covid*[tw] OR

"coronavirus*"[tw] OR

"corona virus*"[tw] OR

ncov*[tw] OR

"n cov*"[tw] OR

sarscov*[tw] OR

"sars cov*"[tw] OR

"2019nCoV*"[tw] OR

"2019 nCoV*"[tw] OR

"sars2*"[tw] OR

"sars 2*"[tw])

I

**"Point-of-Care Testing"[Mesh] OR**

Antigen[tw] OR

“Lateral flow”[tw] OR

RDT[tw] OR

(("Point of Care*"[tw] OR

"Bedside*"[tw] OR

Rapid*[tw])

AND

Test*[tw])

**P**

**1 (99976)**

("Severe Acute Respiratory Syndrome Coronavirus 2"[Supplementary Concept] OR "COVID-19"[Supplementary Concept] OR "Betacoronavirus"[MeSH Terms] OR "Coronavirus"[MeSH Terms] OR "covid*"[Text Word] OR "coronavirus*"[Text Word] OR "corona virus*"[Text Word] OR "ncov*"[Text Word] OR "n cov*"[Text Word] OR "sarscov*"[Text Word] OR "sars cov*"[Text Word] OR "2019ncov*"[Text Word] OR "2019 ncov*"[Text Word] OR "sars2*"[Text Word] OR "sars 2*"[Text Word])

**I**

**2 (816722)**

("Point-of-Care Testing"[MeSH Terms] OR "antigen"[Text Word] OR "lateral flow"[Text Word] OR "RDT"[Text Word] OR (("point of care*"[Text Word] OR "bedside*"[Text Word] OR "rapid*"[Text Word]) AND "test*"[Text Word]))

**3 (1637239)**

2019/12/01:2020/12/11[Date - Publication]

**1 AND 2 AND 3 (2990)**

### Web of Science Core Collection

P

"covid*" OR

"coronavirus*" OR

"corona virus*" OR

"ncov*" OR

"n cov*" OR

"sarscov*" OR

"sars cov*" OR

"2019nCoV*" OR

"2019 nCoV*" OR

"sars2*" OR

"sars 2*"

I

"antigen" OR

"Lateral flow" OR

"RDT" OR

(("Point of Care*" OR

"Bedside*" OR

"Rapid*")

AND

Test*))

P

**1 (56968)**

TS=("covid*" OR "coronavirus*" OR "corona virus*" OR "ncov*" OR "n cov*" OR "sarscov*" OR "sars cov*" OR "2019nCoV*" OR "2019 nCoV*" OR "sars2*" OR "sars 2*")

I

**2 (34058)**

TS=("antigen" OR "Lateral flow" OR "RDT" OR (("Point of Care*" OR "Bedside*" OR "Rapid*")AND Test*))

**1 AND 2 (1568)**

Filter (year to date 2020/12/11)

### Bio_MedRxiv

https://europepmc.org/

P

Covid* OR

Coronavirus* OR

"corona virus*" OR

Ncov* OR

"n cov*" OR

Sarscov* OR

"Sars cov*" OR

2019nCoV* OR

"2019 nCoV*" OR

sars2* OR

"sars 2*"

I

"Antigen*" OR

"Lateral flow*" OR

"RDT" OR

"Rapid test*" OR

"Bedside*" OR

"Point of Care*"

**P AND I (2225)**

(covid* OR Coronavirus* OR "corona virus*" OR ncov* OR "n cov*" OR sarscov* OR "sars cov*" OR 2019nCov* OR "2019 nCov*" OR sars2* OR "sars 2*")

AND

(Antigen OR "Lateral flow" OR RDT OR "Rapid test*" OR Bedside* OR "Point of Care*")

AND

(PUBLISHER:MedRxiv OR PUBLISHER:BioRxiv)

AND

FIRST_PDATE:[2019-12-01 TO 2020-12-11]

#### S3 DETAILS OF QUADAS ASSESSMENT

Figure 1 - Details of QUADAS assessment

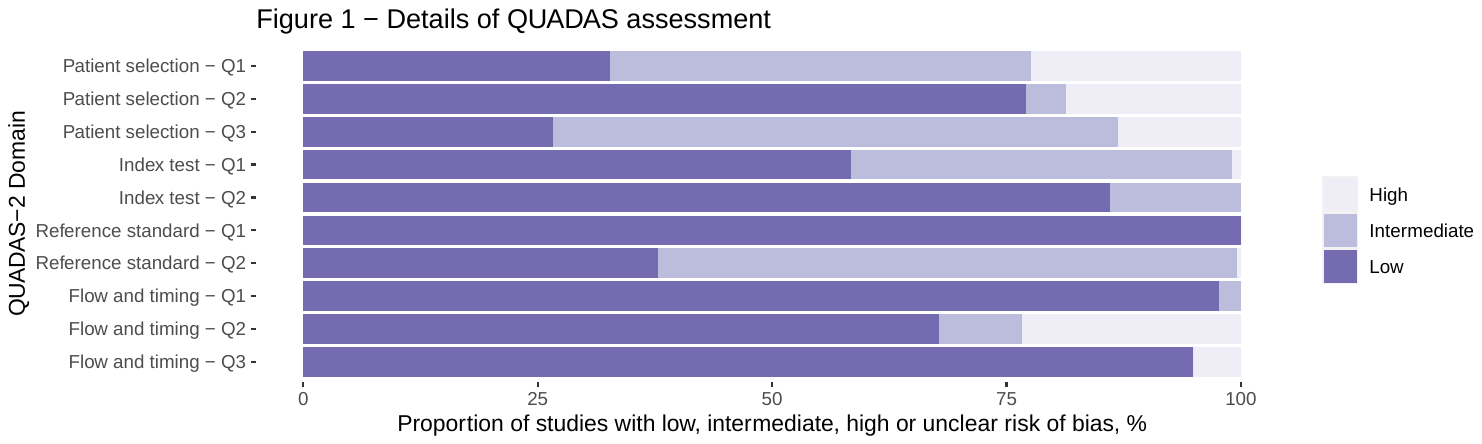

**Domain 1 Patient Selection:**

Risk of Bias: Could the selection of patients have introduced bias?

•	Signaling question 1: Was a consecutive or random sample of patients or specimens enrolled?

We scored ‘yes’ if the study enrolled a consecutive or random sample of eligible patients; ‘no’ if the study selected patients by convenience, and ‘unclear’ if the study did not report the manner of patient selection or unable to tell.

•	Signaling question 2: Was a case-control design avoided?

We scored ‘no’ if the study selected samples with a known rt-PCR results. We scored ‘yes’ if the status of the samples was unknown. We scored ‘unclear’ if we could not tell.

•	Signaling question 3: Did the study avoid inappropriate exclusions?

We scored ’yes’ to studies which included all participants regardless of symptoms or duration of symptoms. We scored ’no’ if studies excluded participants on the basis of symptoms or duration of symptoms. We scored ’unclear’ if we could not tell.

We considered any studies that included patients based on a previous positive rt-PCR results as monitoring studies and thus judged the applicability of the study population to be of ‘high concern’ (see below).

- Risk of Bias was scored ‘low concern’ if studies score ‘yes’ on all the question, ‘unclear concern’ if questions are answered with ‘yes’ and ‘unclear’, ‘intermediate concern’ if one question is answered with ‘no’, ‘high concern’ if two or more questions are answered with ‘no’.

Applicability: Are there concerns that the included patients and setting do not match the review question?

We were interested in how Ag-RDT performs in patients whose specimens were evaluated as they would be in routine practice. We scored ’low concern’ if the study was conducted in a routine practice setting. We scored ‘high concern’ if Ag-RDT were evaluated for end of quarantine evaluation or monitoring. We scored ‘unclear’ if we could not tell.

**Domain 2: Index Test**

Risk of Bias: Could the conduct or interpretation of the index test have introduced bias?

•	Signaling question 1: Were the index test results interpreted with knowledge of the results of the reference standard?

We answered ’yes’ if the study interpreted the result of Ag-RDT blinded to the result of the reference standard or for studies in which Ag-RDT was performed on fresh specimens, since reference standard results would be unavailable at the time of test interpretation. We answered ’no’ if the study did not interpret the result of Ag-RDT blinded to the result of the reference standard. We answered ’unclear’ if stored specimens were tested or we could not tell if the index test results were interpreted without knowledge of the reference standard results.

•	Signaling question 2: If a threshold was used, was it prespecified?

We answered ’yes’ if the threshold was prespecified or if the tests was performed by IFU. We scored ’no’ if the threshold was not prespecified, and ’unclear’ if we could not determine if the threshold was prespecified or not.

- Risk of Bias was scored ‘low concern’ if studies score ‘yes’ on all the question, ‘unclear concern’ if questions are answered with ‘yes’ and ‘unclear’, ‘intermediate concern’ if one question is answered with ‘no’, ‘high concern’ if two or more questions are answered with ‘no’.

Applicability: Are there concerns that the index test, its conduct, or its interpretation differ from the review question? If index test methods vary from those specified in the review question, concerns about applicability may exist. We judged ’high concern’ if the test procedure was inconsistent with the manufacturer recommendations, ’low concern’ if the test procedure was consistent with the manufacturer recommendations, and ’unclear concern’ if we could not tell.

**Domain 3: Reference Standard**

Risk of Bias: Could the reference standard, its conduct, or its interpretation have introduced bias?

•	Signaling question 1: Is the reference standard likely to correctly classify the target condition?

Viral culture is considered the gold standard for SARS-CoV-2 detection. Since viral culture is available in research settings only, NAAT is the considered routine standard for SARS-CoV-2 testing. However, the accuracy of this reference standard is not 100%, especially late in the disease, where it varies widely across the different non-respiratory samples and may detect non-viable virus Still, given that viral loads measured in NAATs correlate well with Antigen, we scored ‘yes’ for all studies using a NAAT as reference standard.

•	Signaling question 2: Were the reference standard results interpreted without knowledge of the results of the index test?

We scored ‘yes’ if the test was performed ahead of the Ag-RDT or blinding was specifically reported. We scored ‘unclear’ if we could not tell.

- Risk of Bias is scored ‘low concern’ if studies score ‘yes’ on all the question, ‘unclear concern’ if questions are answered with ‘yes’ and ‘unclear’, ‘intermediate concern’ if one question is answered with ‘no’, ‘high concern’ if two or more questions are answered with ‘no’.

Applicability: Are there concerns that the target condition as defined by the reference standard does not match the question?

We judged applicability to be of ‘low concern’ for all studies.

**Domain 4: Flow and Timing**

Risk of Bias: Could the patient flow have introduced bias?

•	Signaling question 1: Was there an appropriate interval between the index test and reference standard?

We expected specimens for Ag-RDT and the reference standards to be obtained at the same time and answered ’yes’ for all studies that meet these criteria. We answered ‘unclear’ if we could not tell.

•	Signaling question 2: Did all patients receive the same reference standard?

We answered this question ‘yes’ for all studies that used the same rt-PCR for all samples and ‘no’ if the samples were analyzed by different types of rt-PCR. We scored ‘unclear’ if we could not tell the used rt-PCR.

•	Signaling question 3: Were all patients included in the analysis?

We determined the answer to this question by comparing the stated population size with the number of samples included in the two-by-two tables. We answered ’yes’ if the whole population was included in the analysis or any excluded samples were reasoned for. We answered ‘no’ if samples were excluded without a given reason. We answered ‘unclear’ if we could not tell.

- Risk of Bias is scored ‘low concern’ if studies score ‘yes’ on all the question, ‘unclear concern’ if questions are answered with ‘yes’ and ‘unclear’, ‘intermediate concern’ if one question is answered with ‘no’, ‘high concern’ if two or more questions are answered with ‘no’.

#### S4 LIST OF ORIGINAL DATA

Figure 2 – List of original data

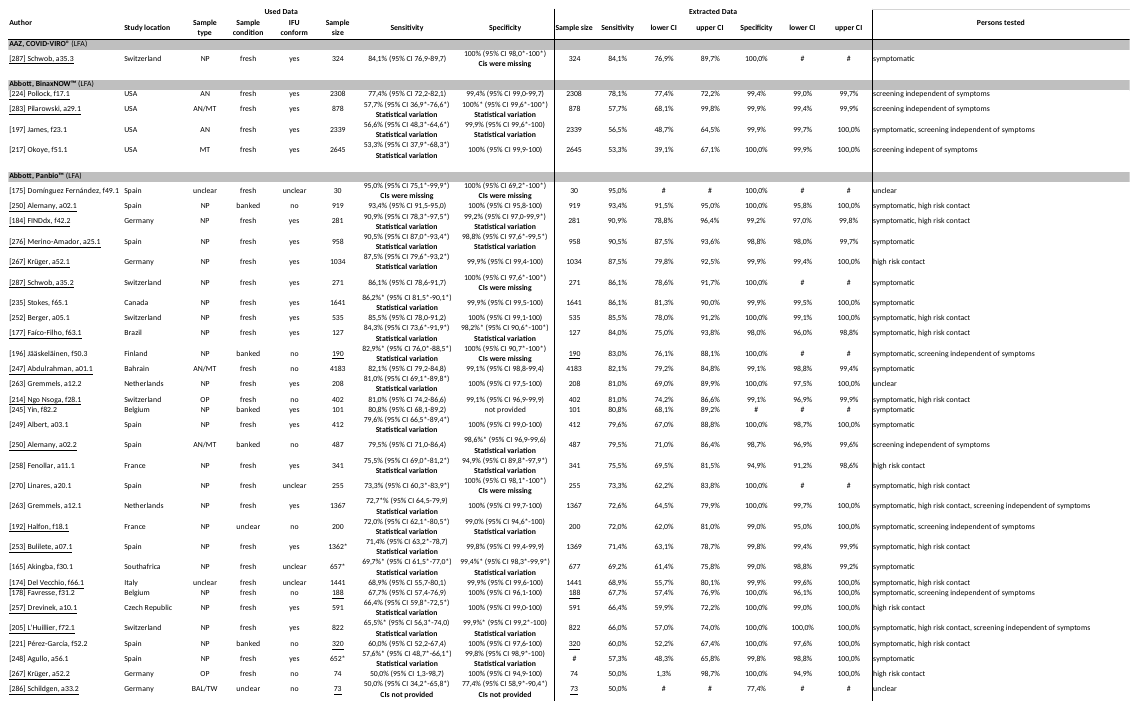

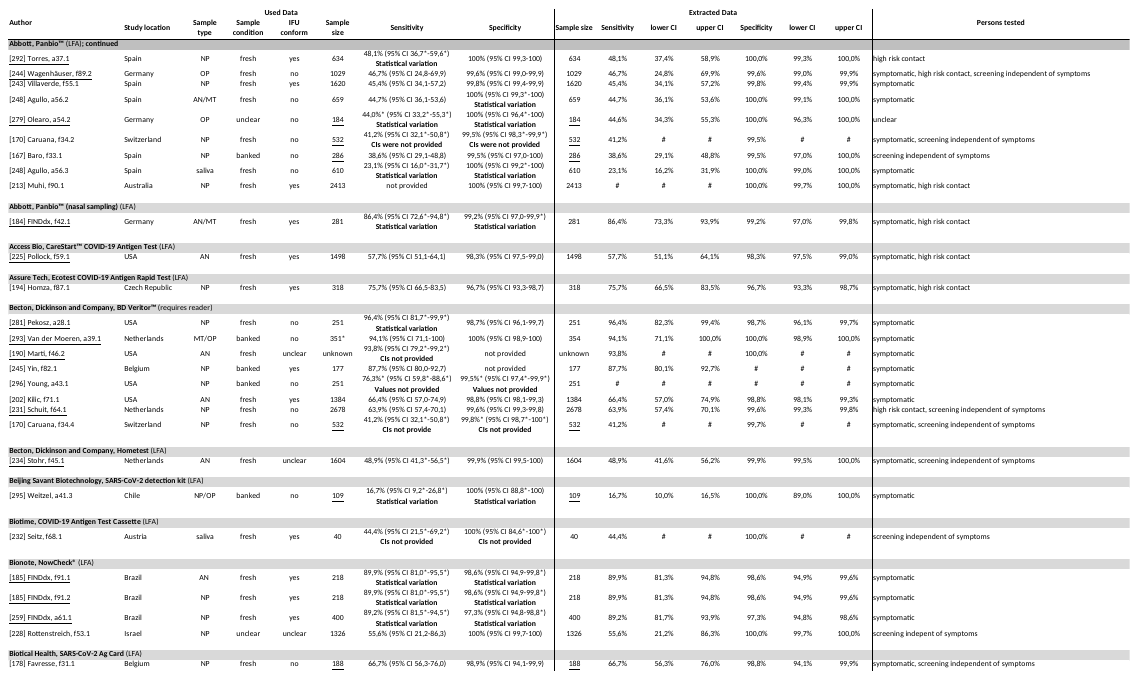

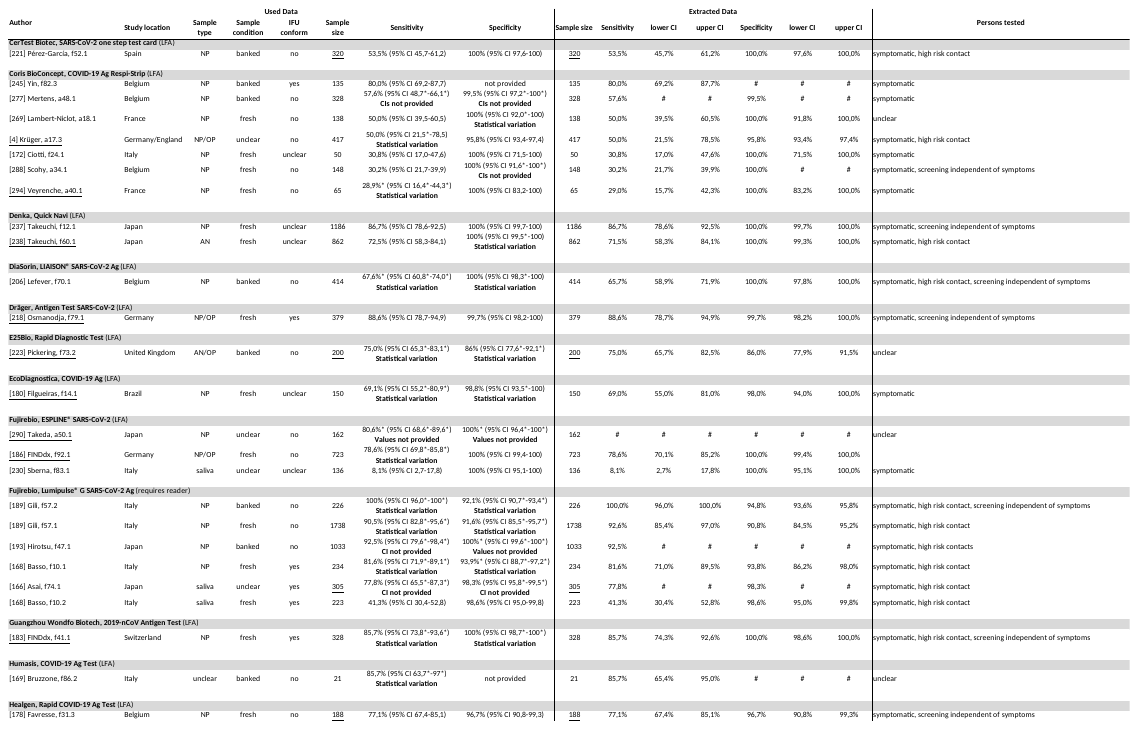

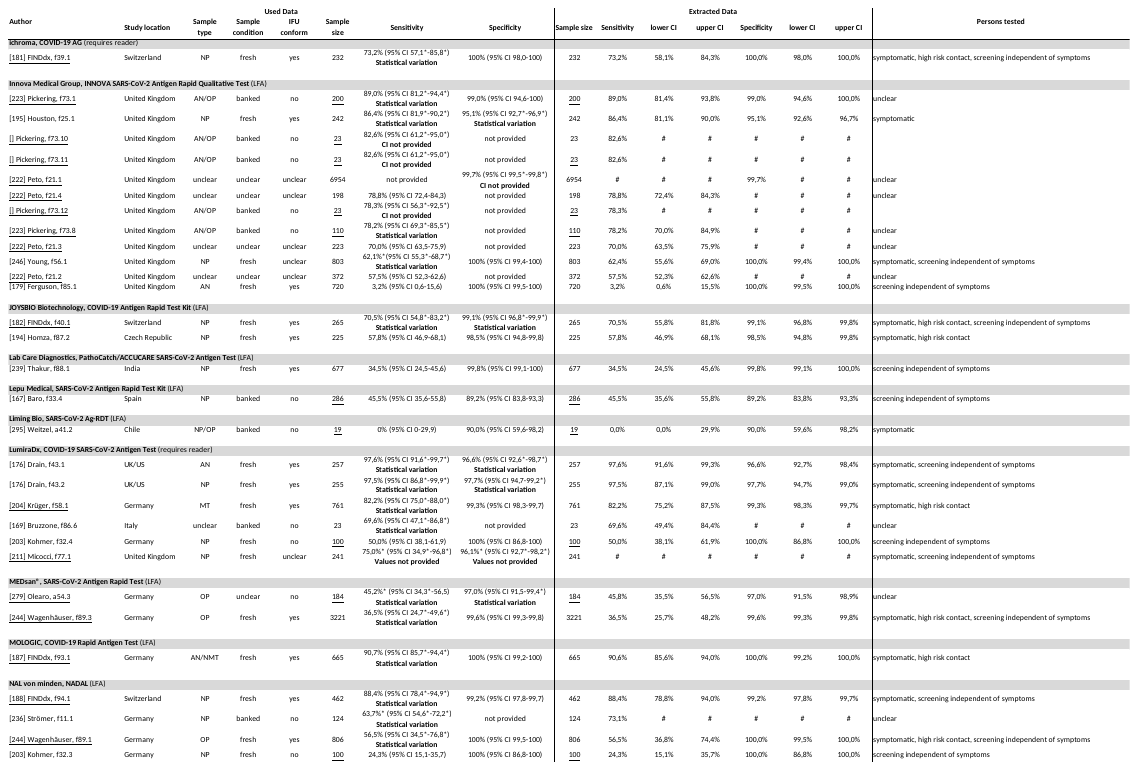

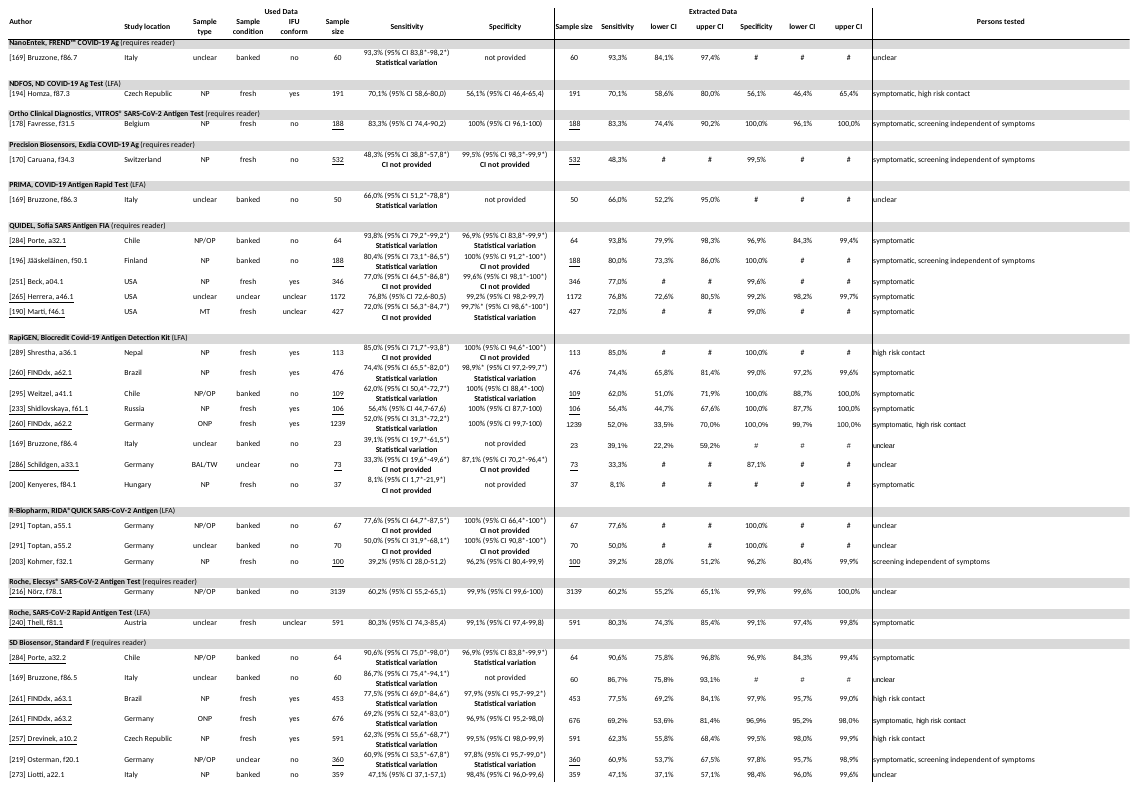

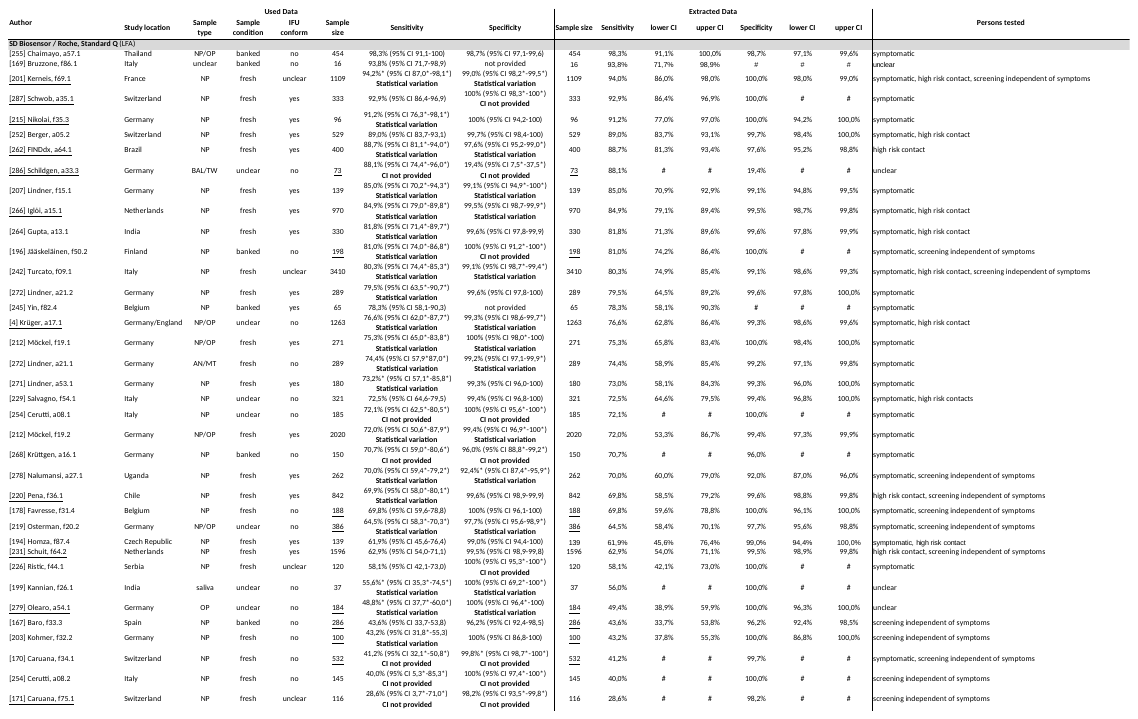

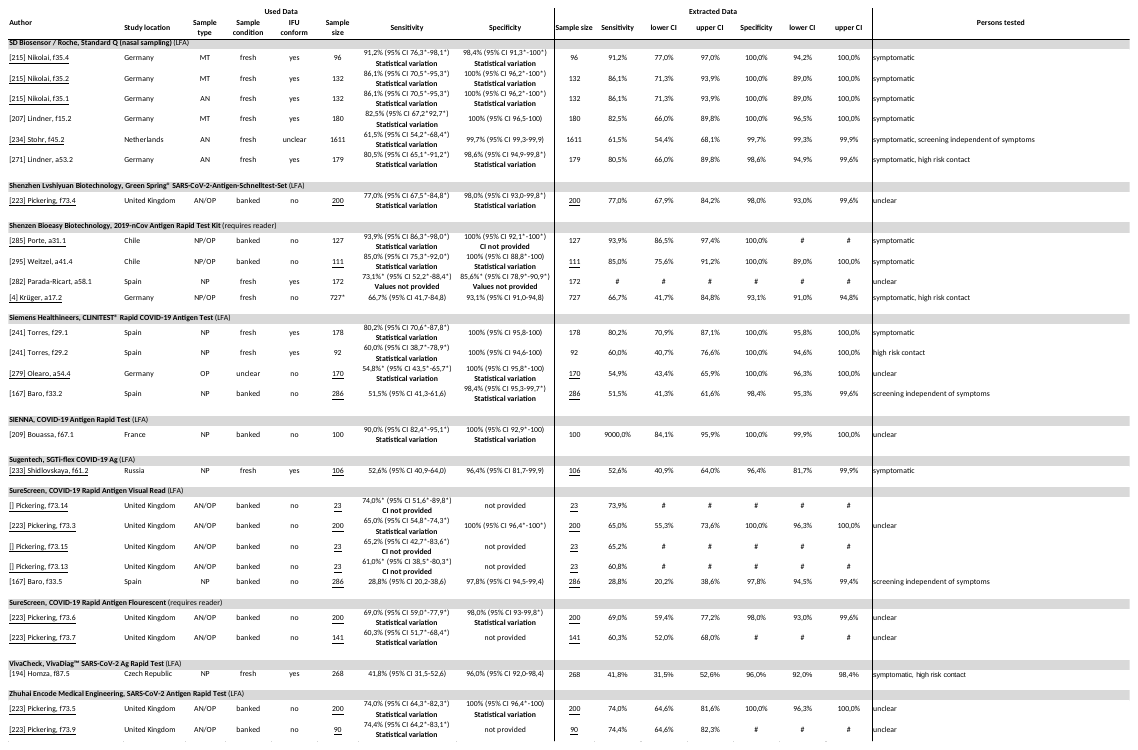

Caption: Naming convention column “author”: number in brackets relates to the list of sources. Letters behind the author’s last name differentiates the data set from other data sets by the same author.

IFU = instructions for use; NP = nasopharyngeal; OP = oropharyngeal; AN = anterior nasal; MT = mid turbine; LRT = lower respiratory tract; BAL/TW = bronchoalveolar lavage and throat wash; CI = confidence interval; Ar = Aruba; Ut = Utrecht; sc = self-collected; pc = professional-collected; ER = emergency room; tr = travelers.

#### S5 Forest plots of all Ag-RDTs

Figure 3 - Forest plots of all Ag-RDTs

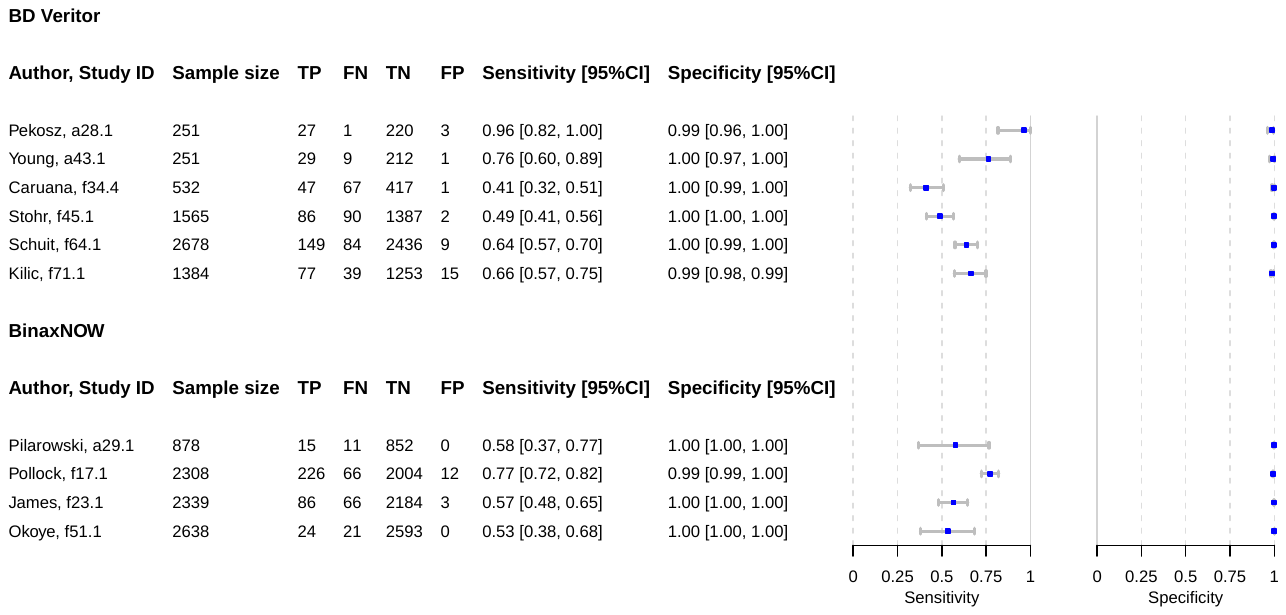

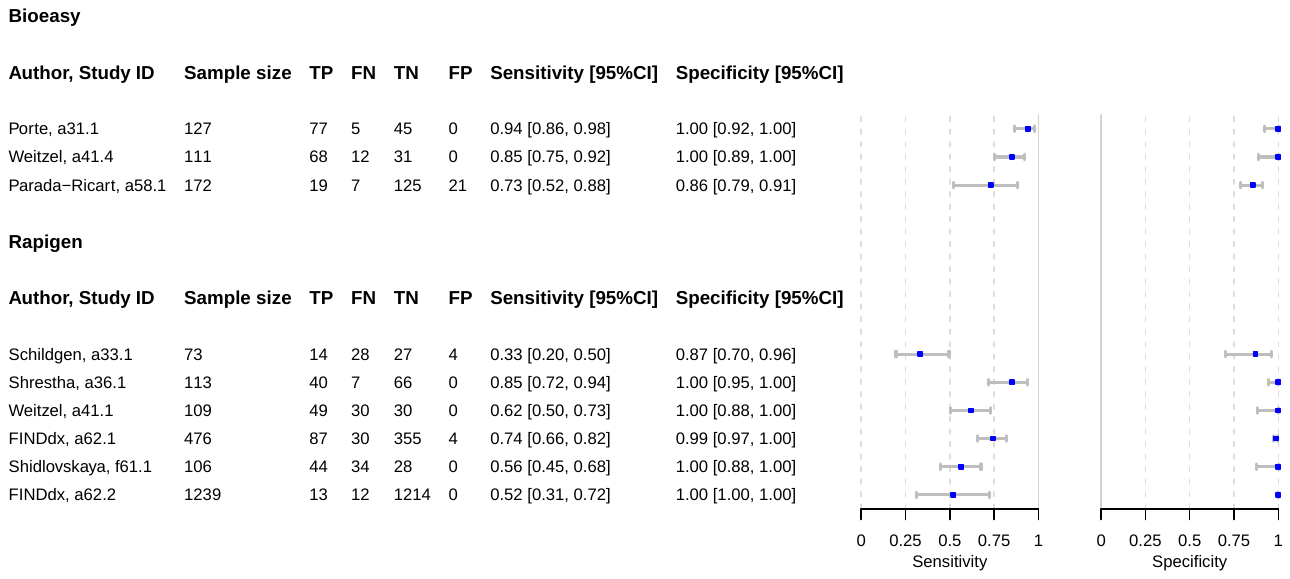

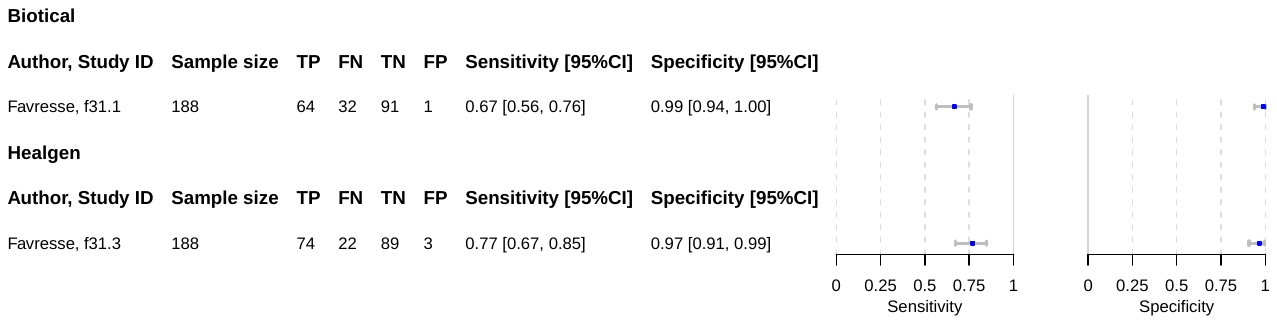

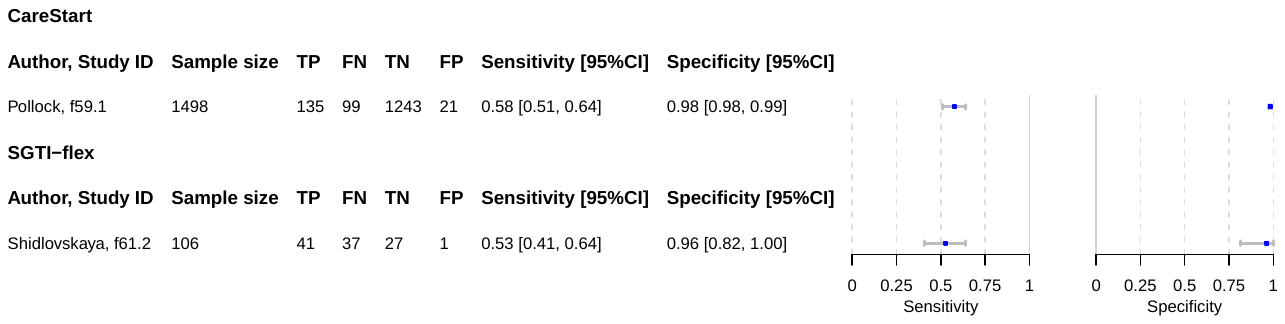

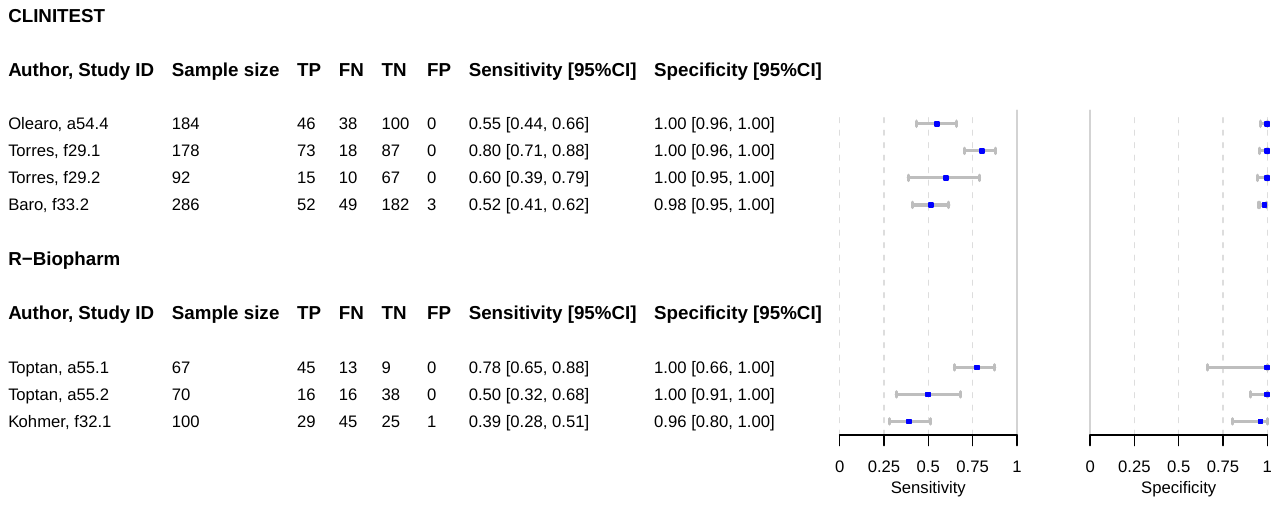

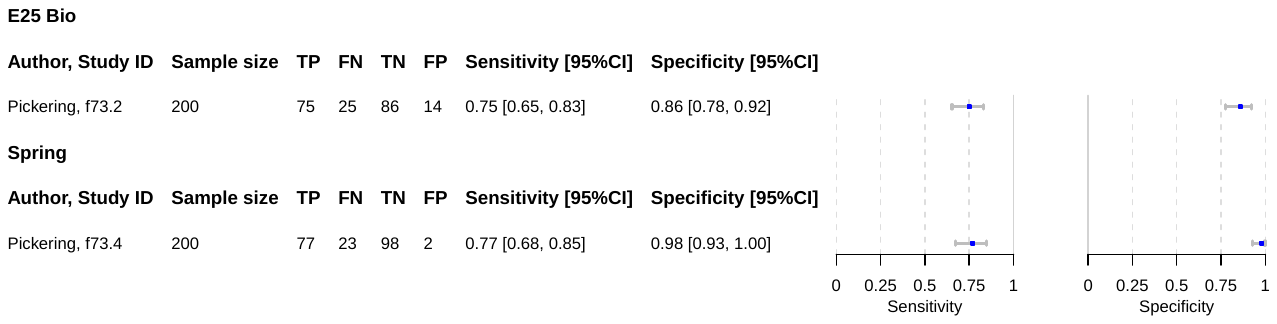

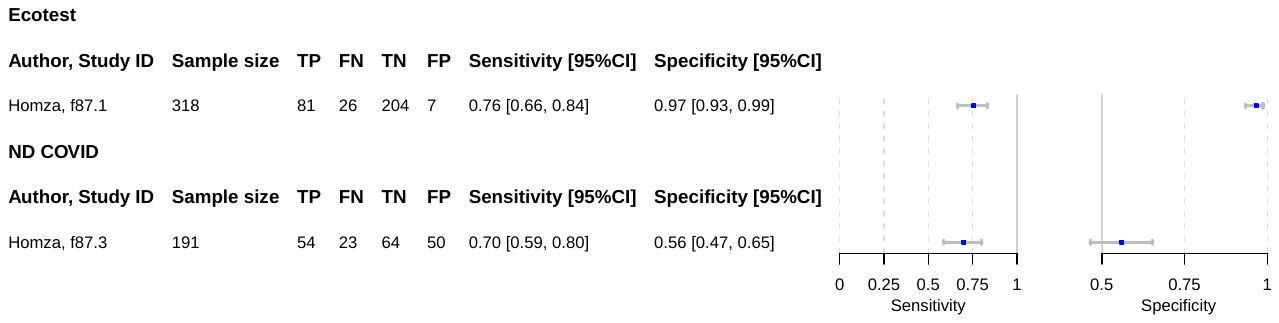

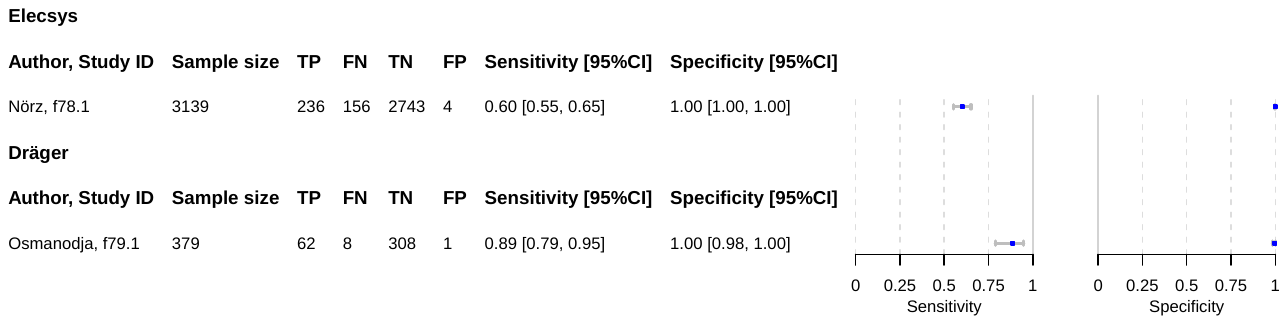

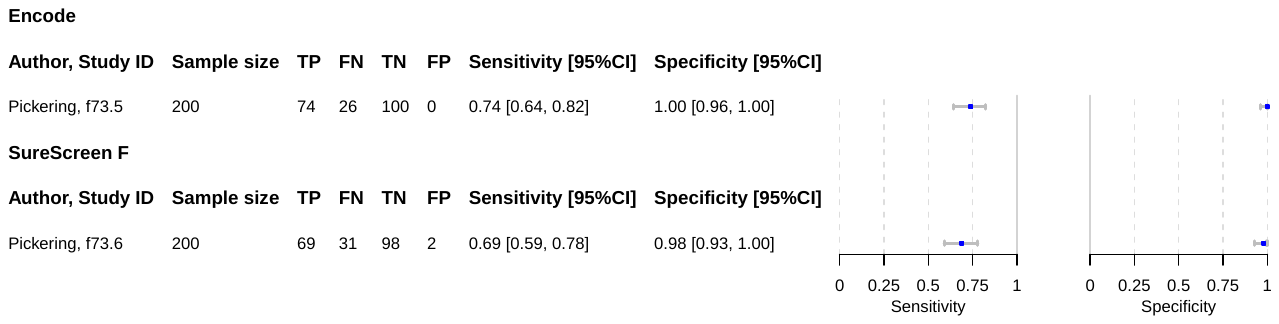

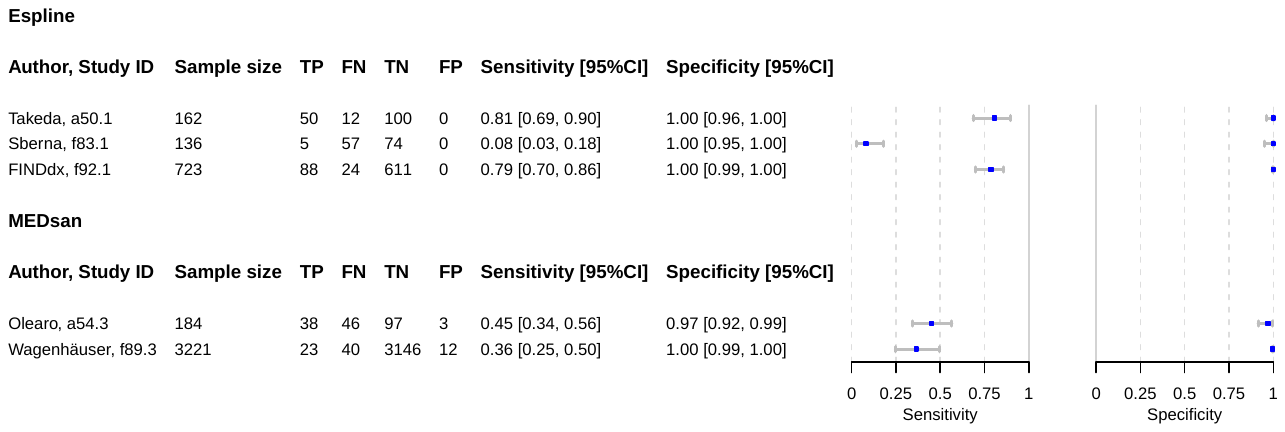

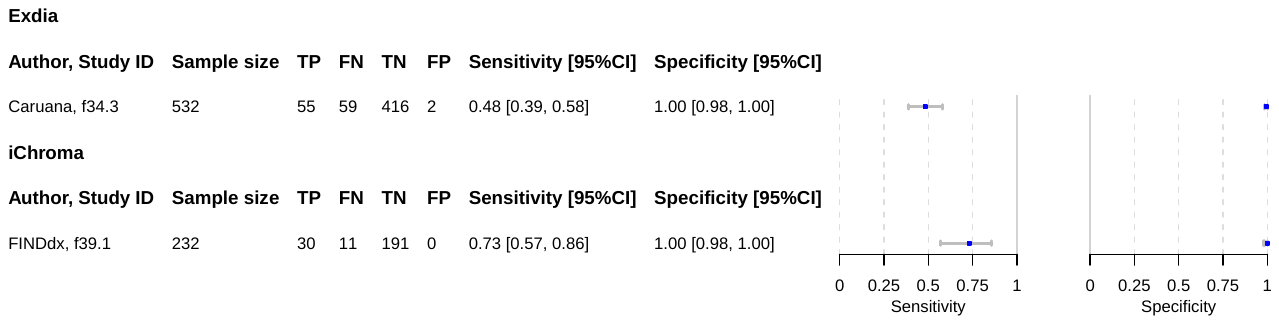

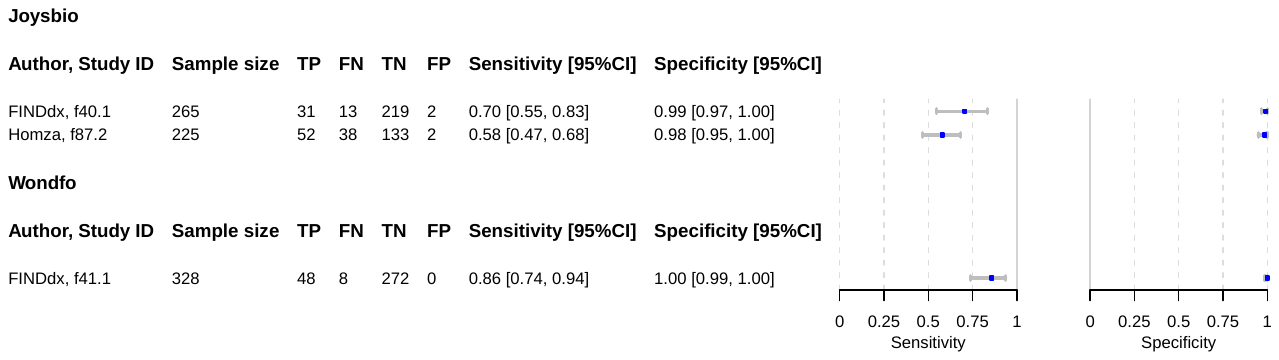

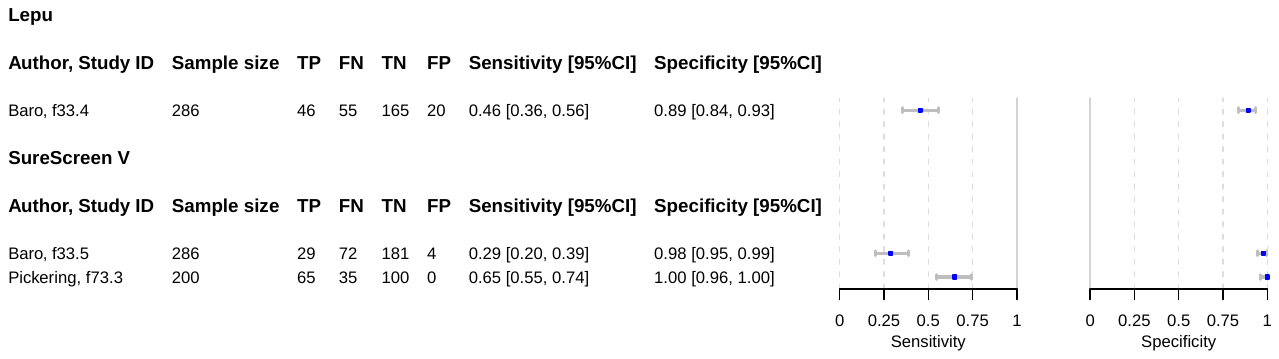

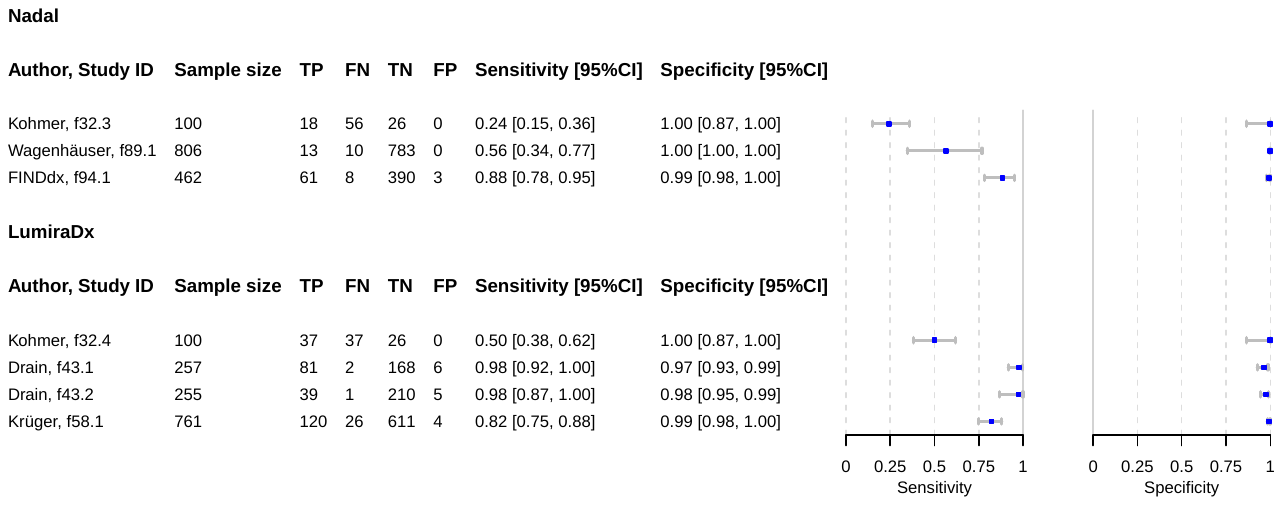

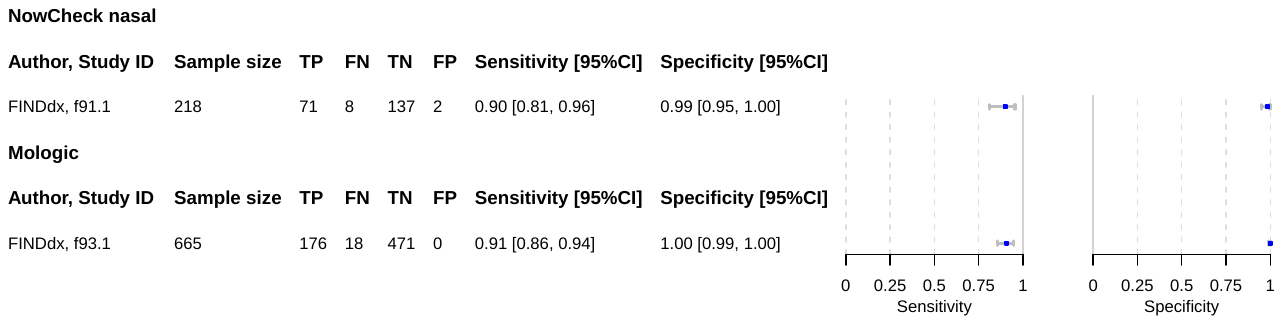

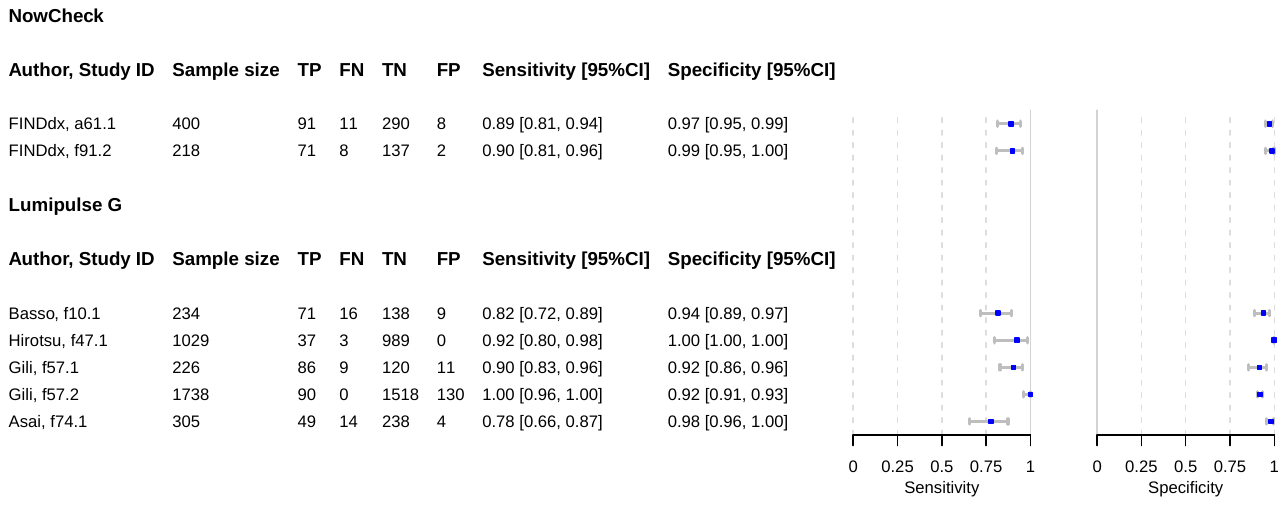

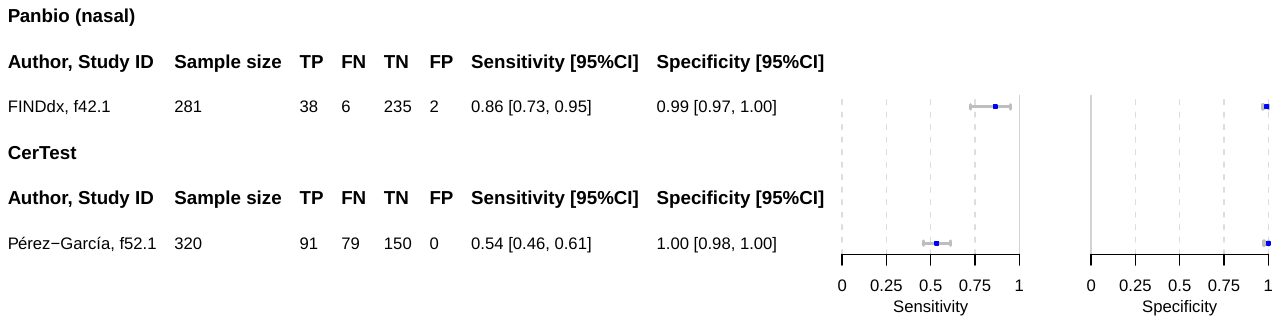

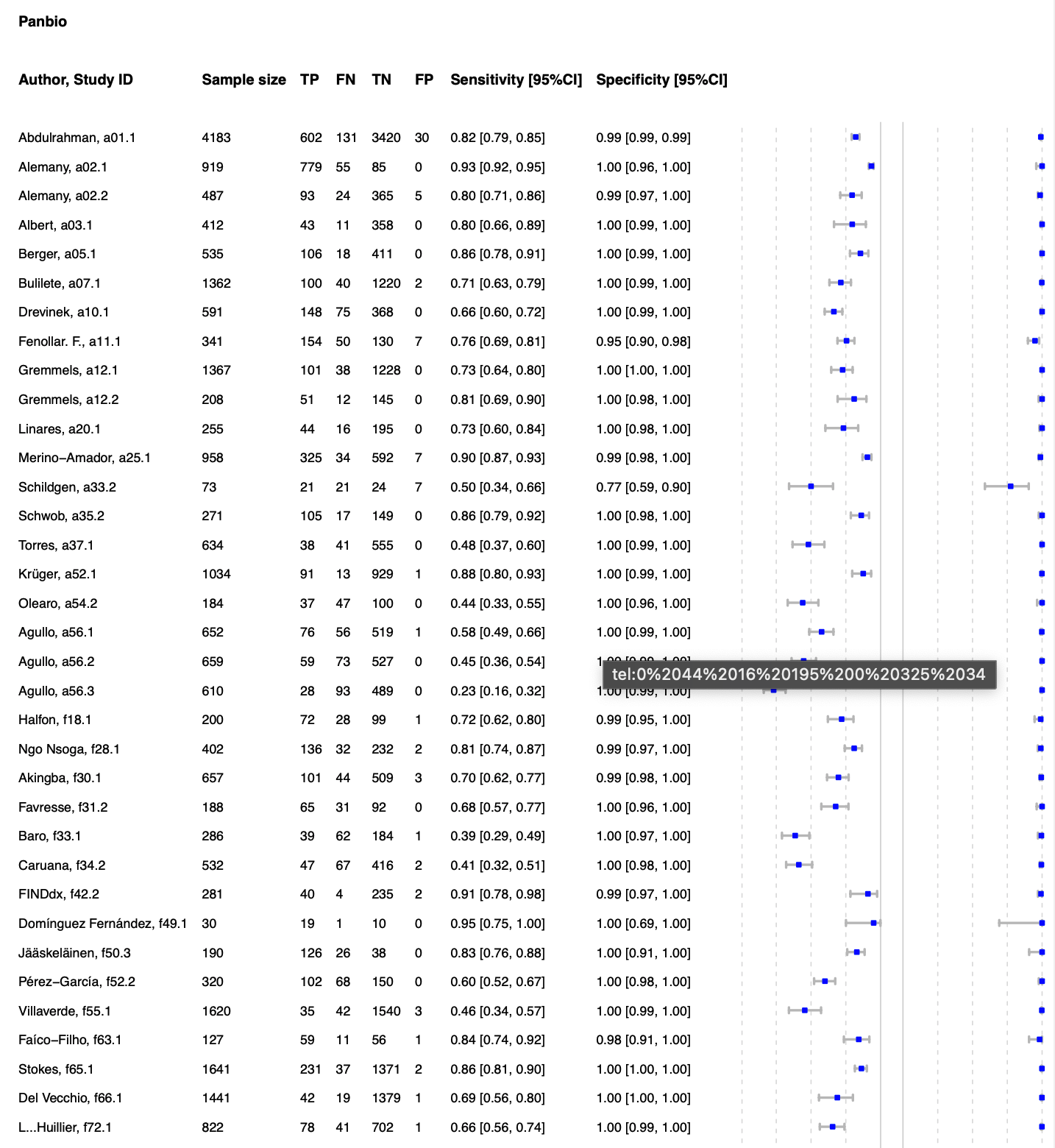

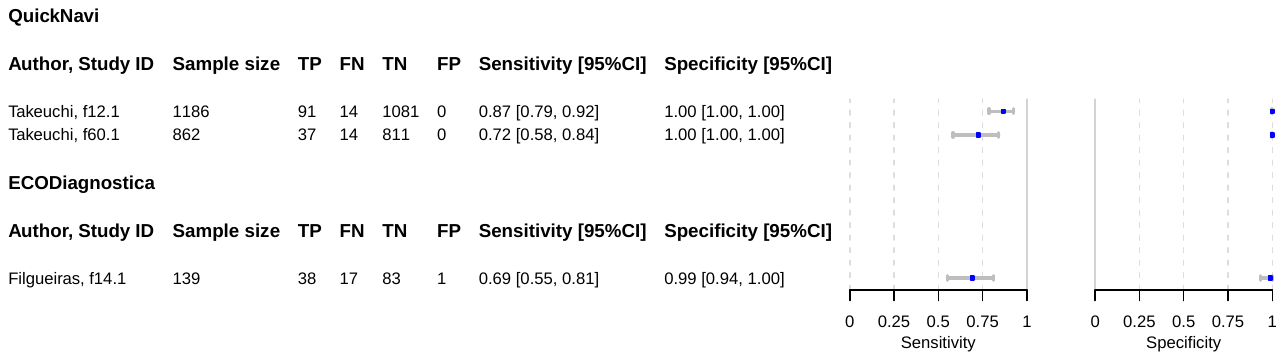

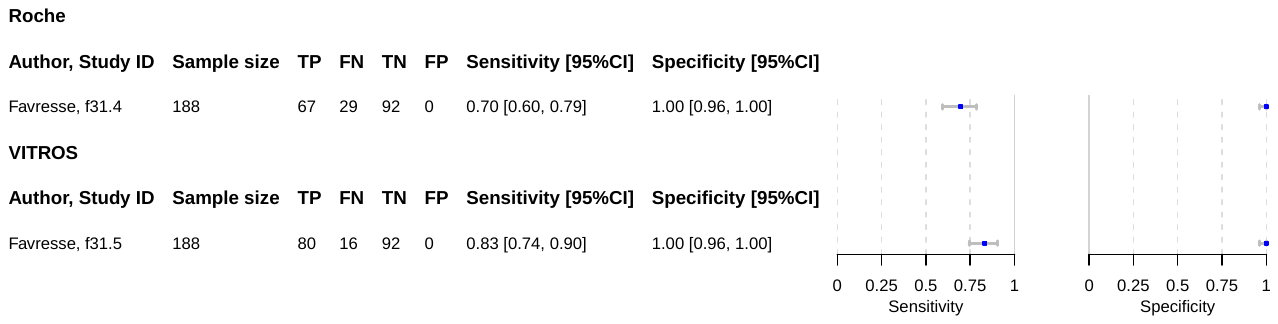

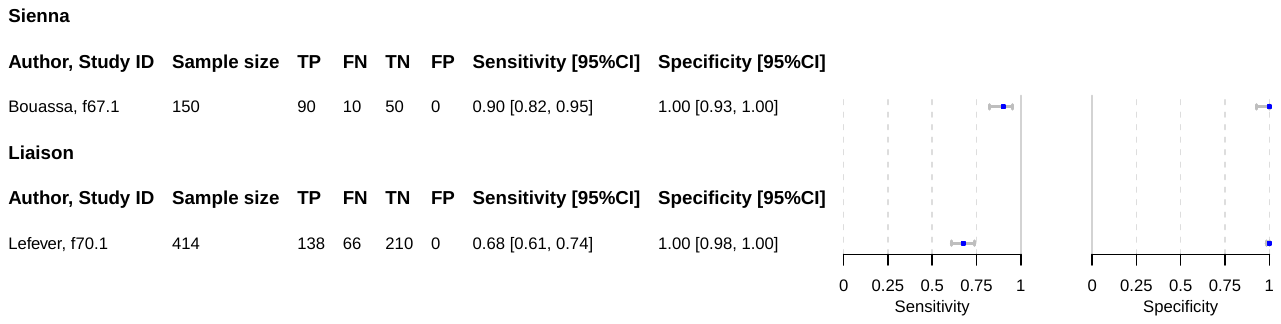

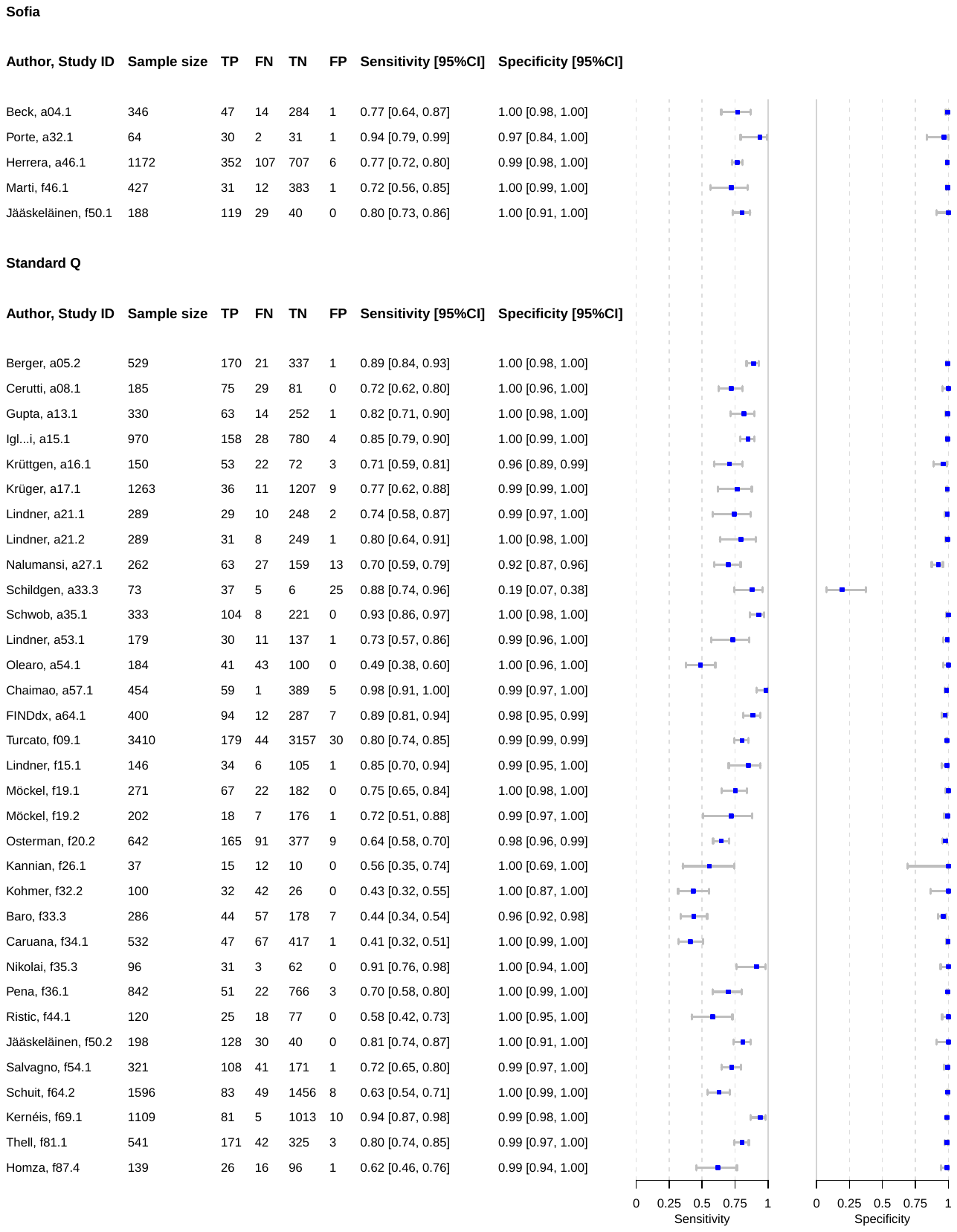

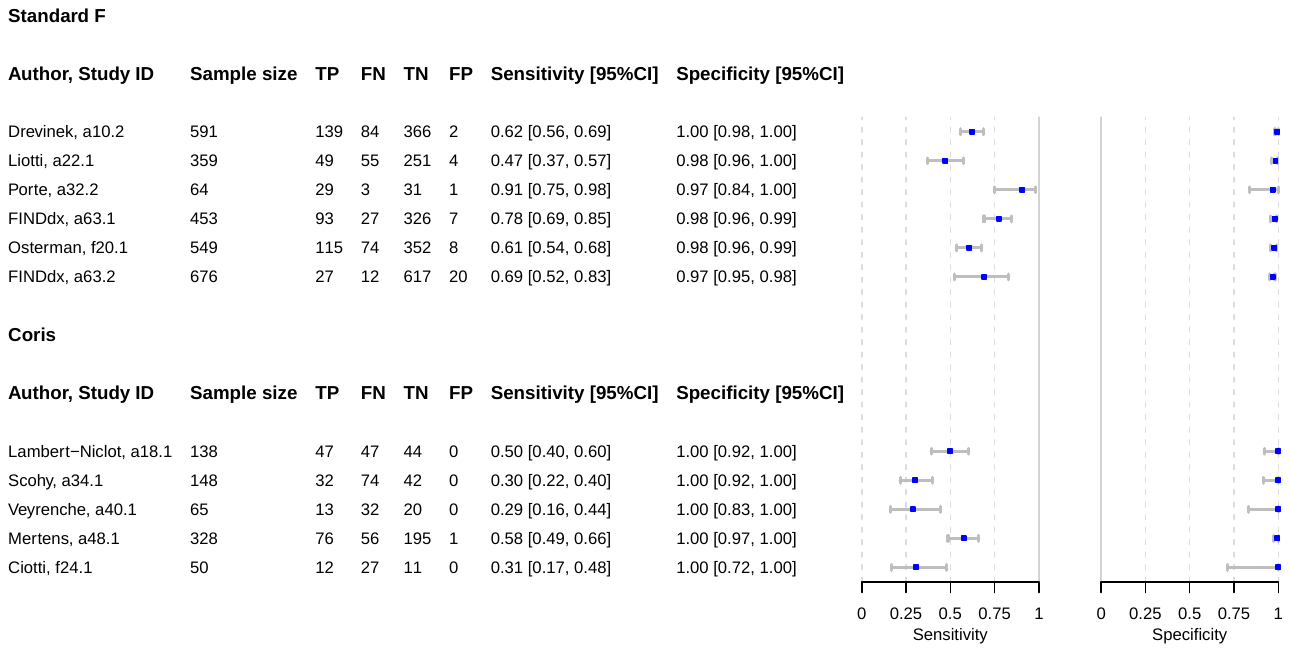

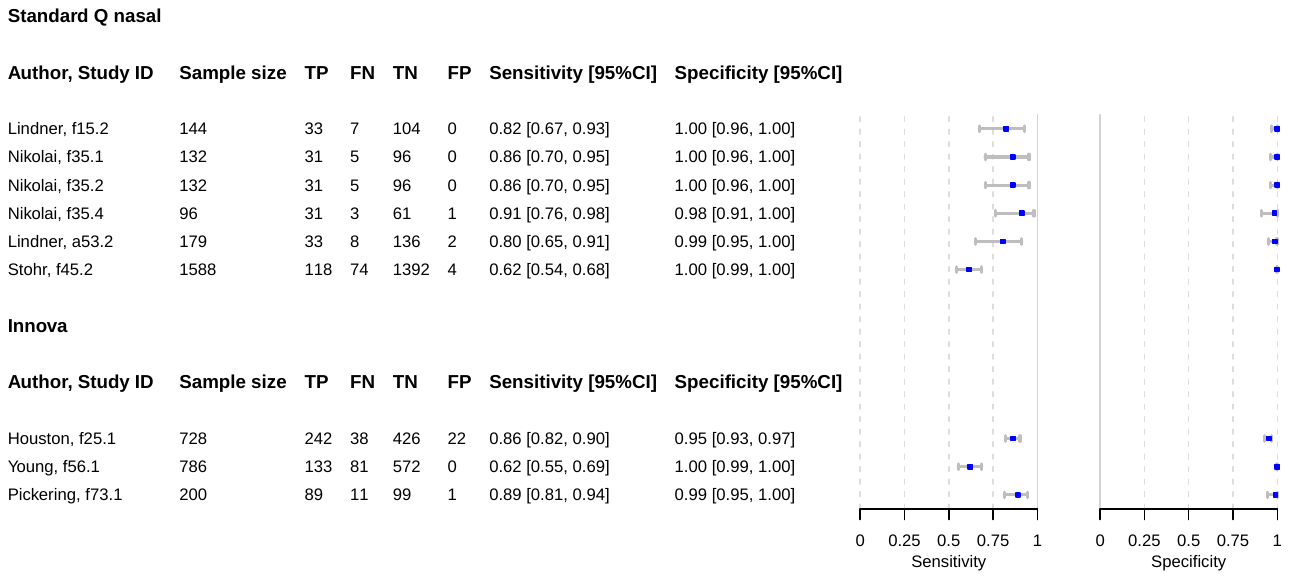

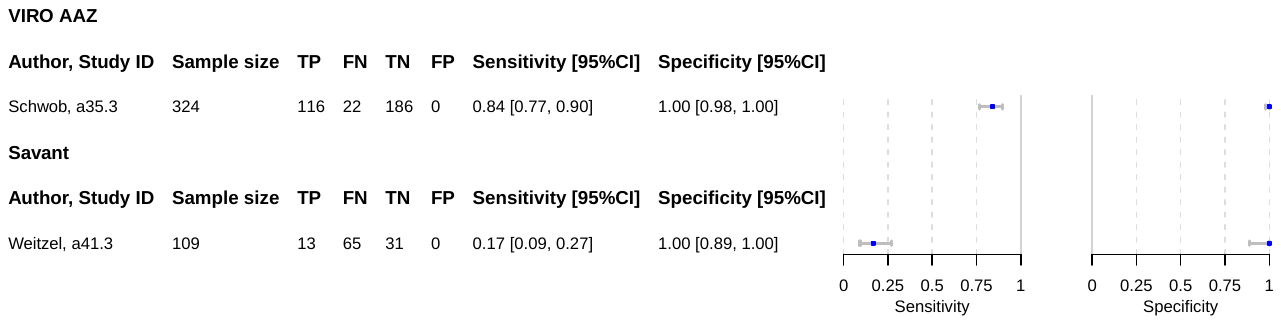

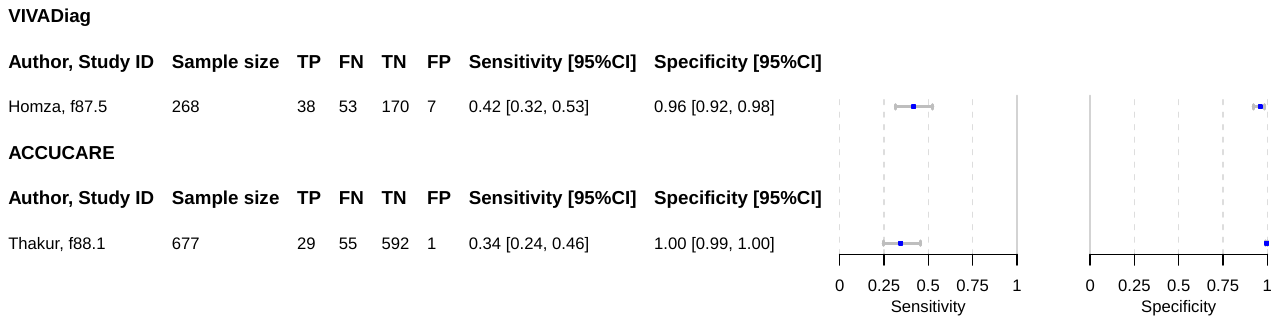

Caption: TP = true positive; FP = false positive; FN = false negative; TN = true negative; CI = confidence interval; NP = nasopharyngeal; OP = oropharyngeal; AN = anterior nasal; MT = mid-turbinate; * = studies with less than 20 PCR positives

*Panbio: Ut=Utrecht site; Ar=Aruba site; Standard Q: ER=Emergency Room; tr=travelers; sc=self-collected nasal swab; pc=professionally collected nasal swab*

**S6 HSROC curves for top-performing Ag-RDTs**

Figure 4 – HSROC curve Standard Q Ag-RDT

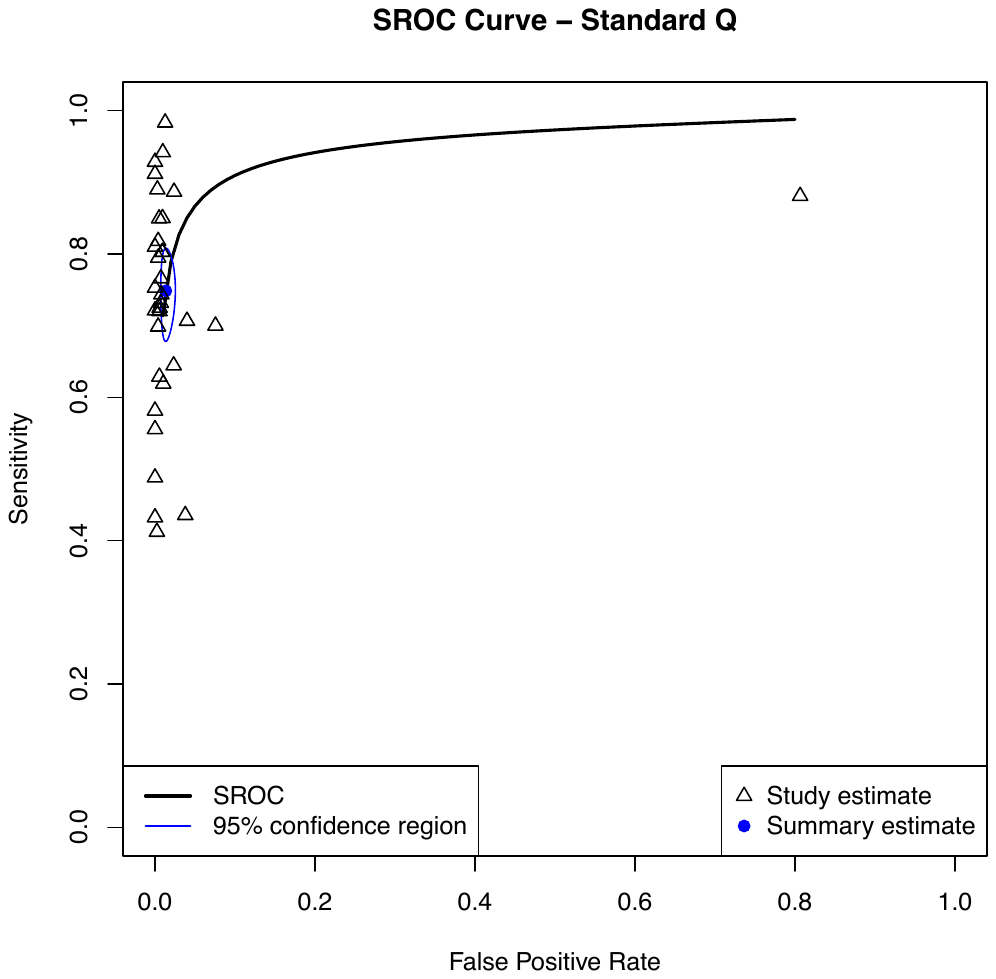

**HSROC = Hierarchical summary receiver-operating characteristic*

Figure 5 – HSROC curve LumiraDx Ag-RDT

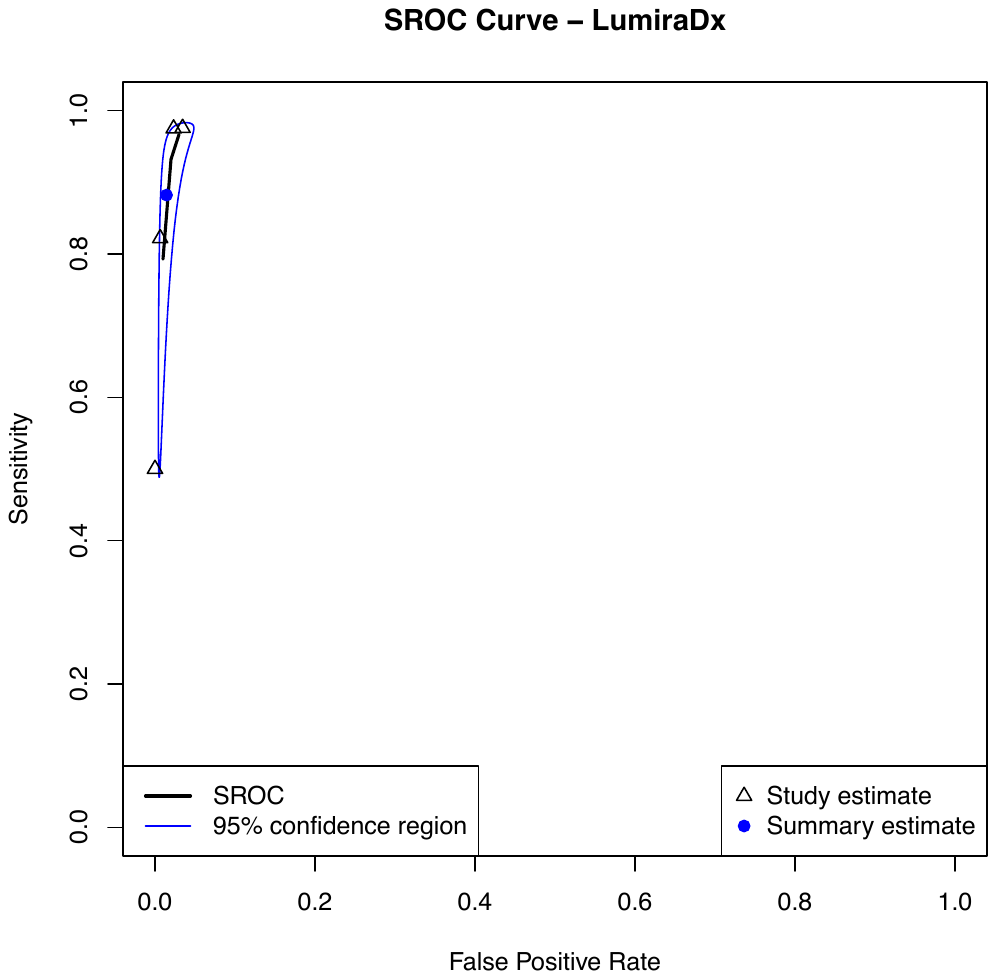

**HSROC = Hierarchical summary receiver-operating characteristic*

#### S7 Forest plots for subgroups analysis by sample type

Figure 6 – Forest plot for anterior nasal and mid-turbinate samples

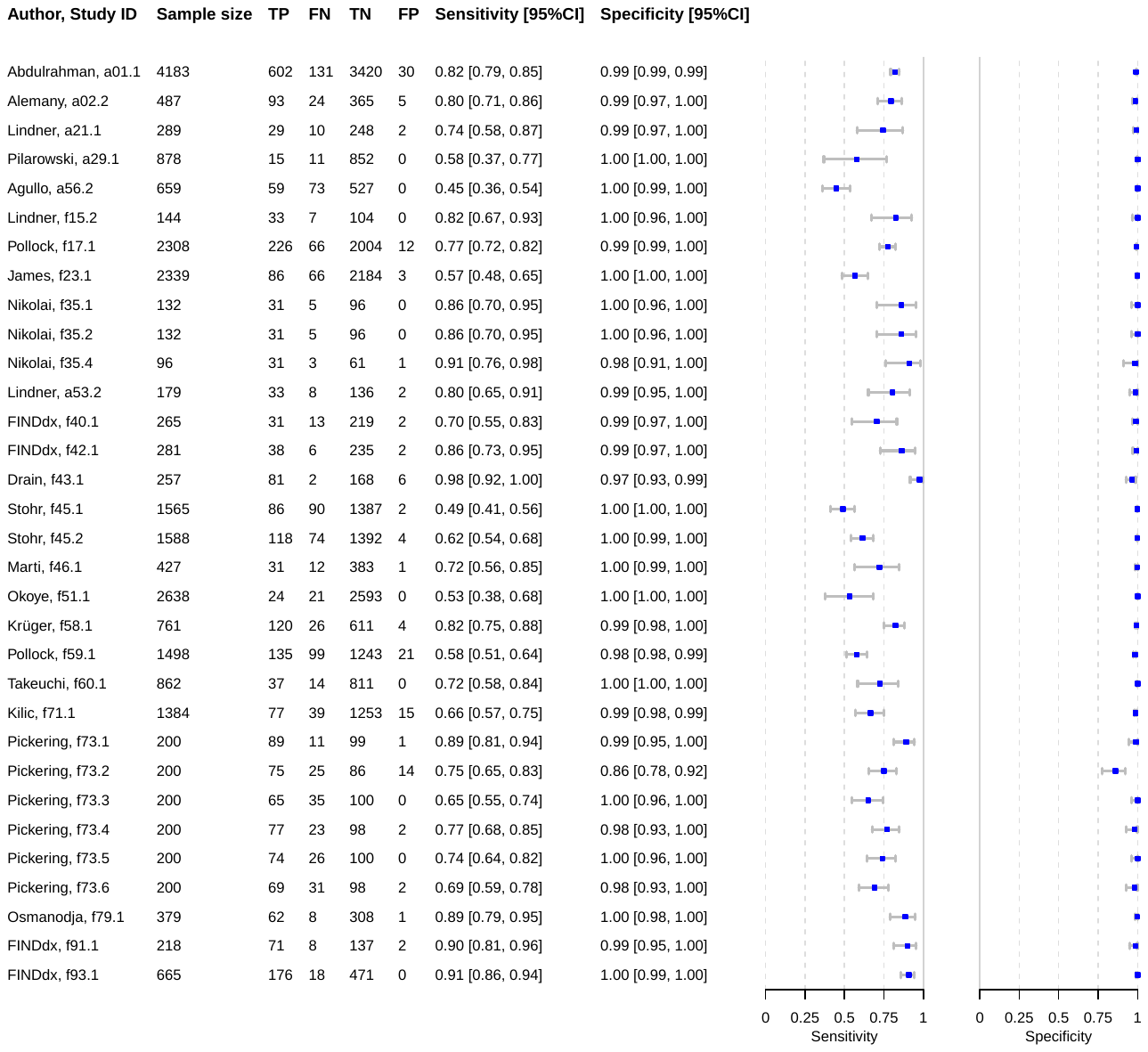

Caption: CI = confidence interval

Figure 7 - Forest plot for nasopharyngeal and nasopharyngeal/oropharyngeal samples

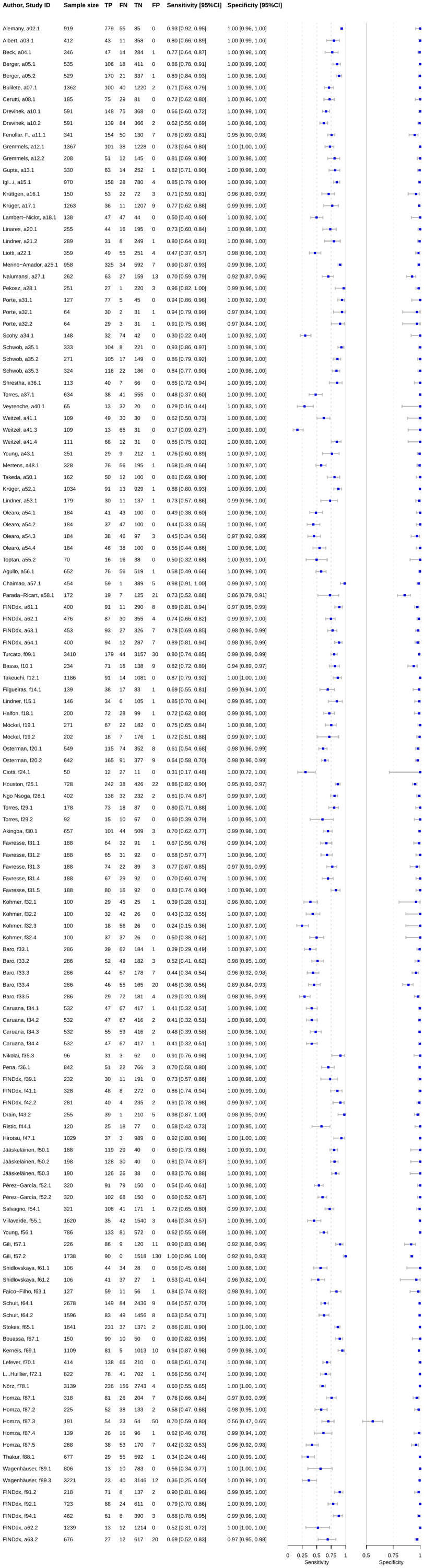

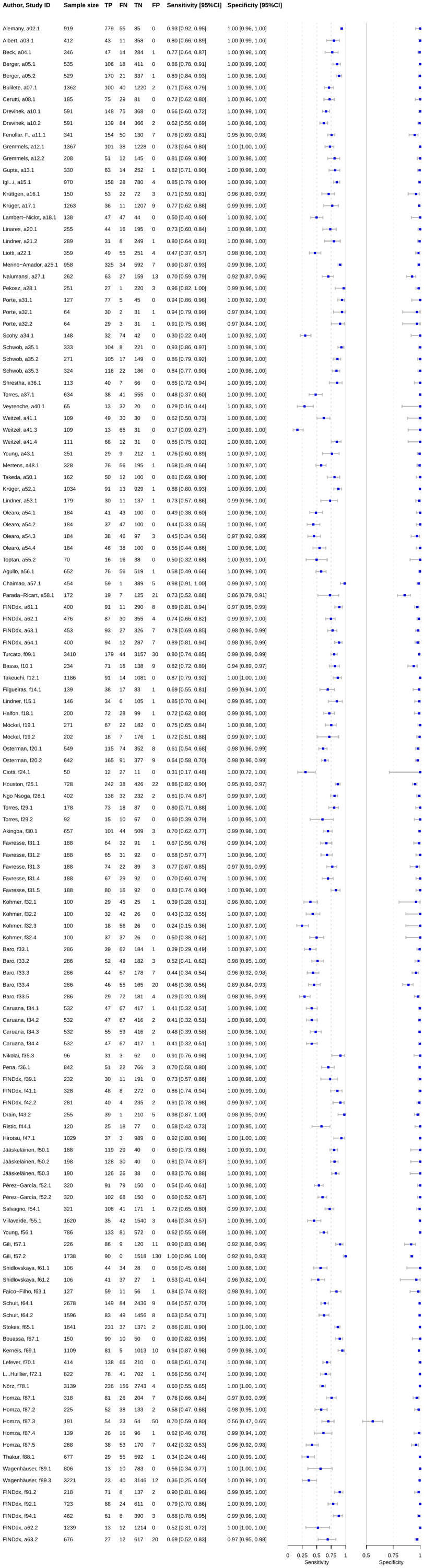

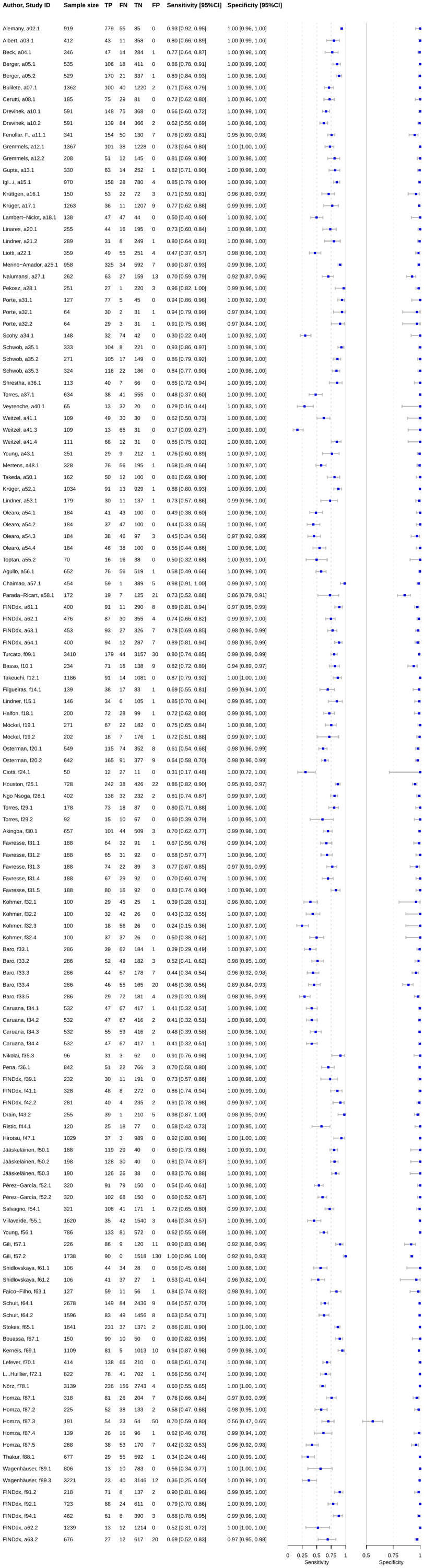

Caption: TP = true positive; FP = false positive; FN = false negative; TN = true negative; CI = confidence interval; NP = nasopharyngeal; OP = oropharyngeal; AN = anterior nasal; MT = mid-turbinate;

Figure 8 - Forest plot for oropharyngeal samples

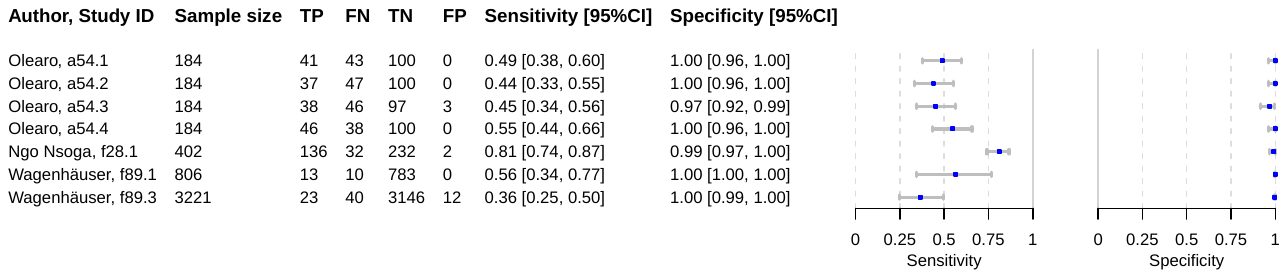

Caption: CI = confidence interval

#### S8 Forest plots for subgroups analysis by symptomatic vs. asymptomatic

Figure 9 - Forest plot of asymptomatic patients

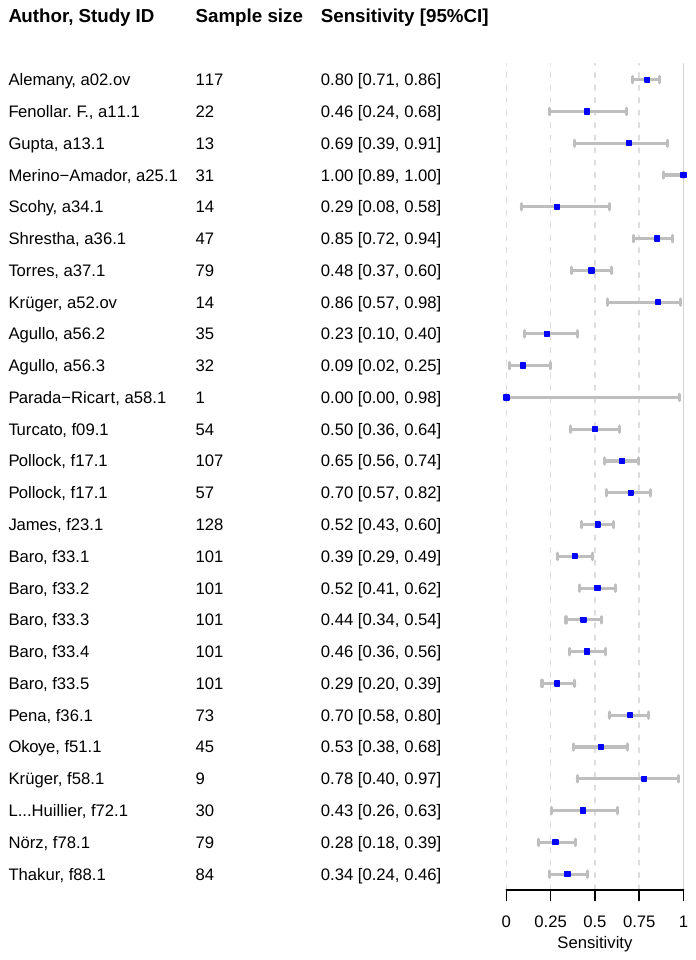

*Caption: CI = confidence interval*

Figure 10 - Forest plot of symptomatic patients

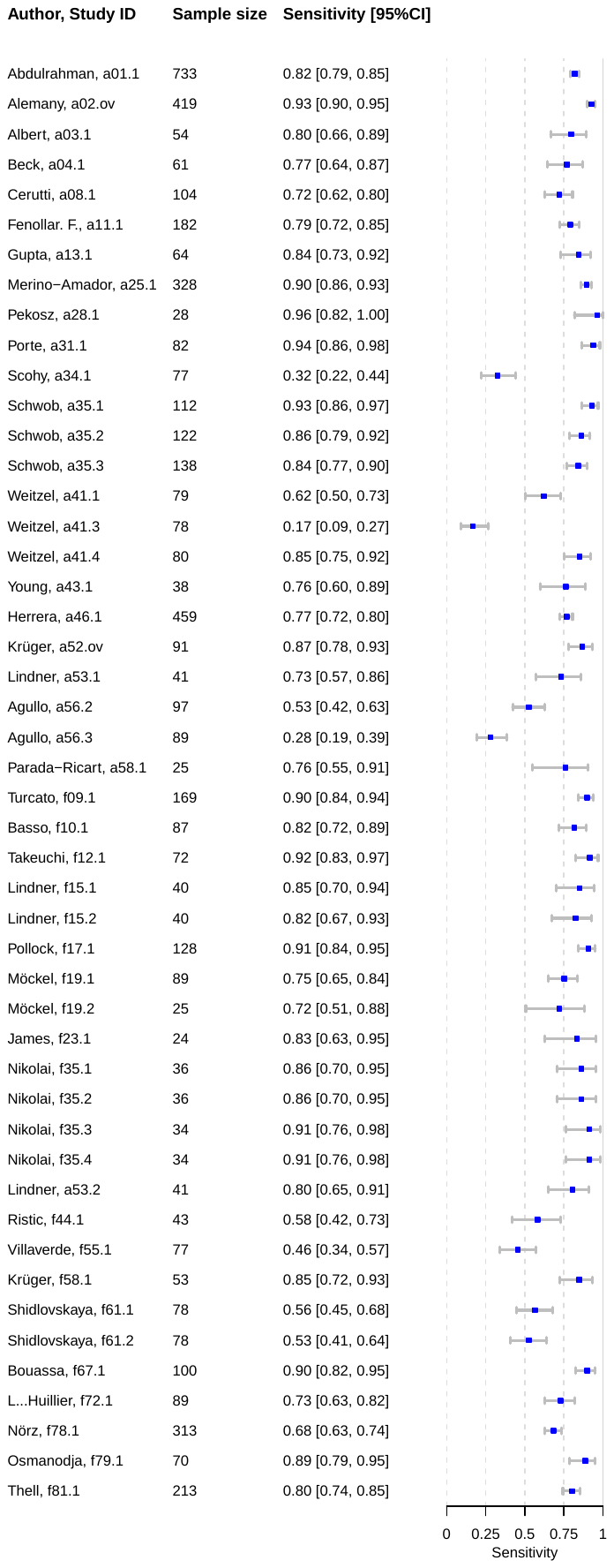

*Caption: CI = confidence interval*

#### S9 Forest plots for subgroups analysis by symptom duration

Figure 11 - Forest plot of patients with symptom onset greater than seven days

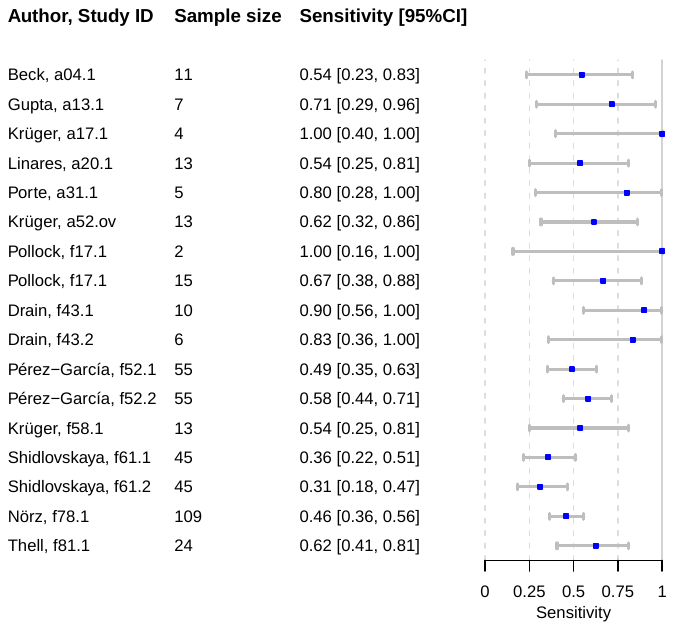

*Caption: CI = confidence interval*

Figure 12 - Forest plot of patients with symptom onset less than seven days

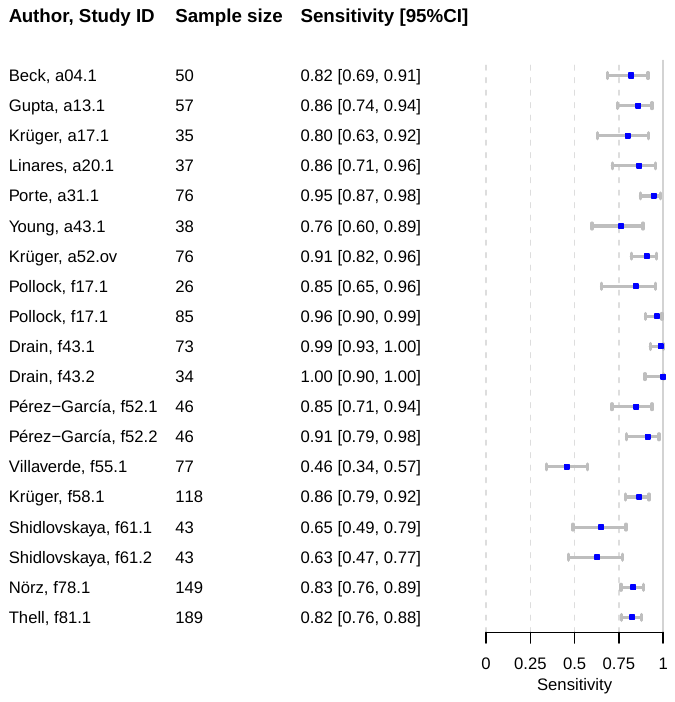

*Caption: CI = confidence interval*

#### S10 Forest plots for subgroups analysis by CT-values

Figure 13 - forest plot for CT values greater 20

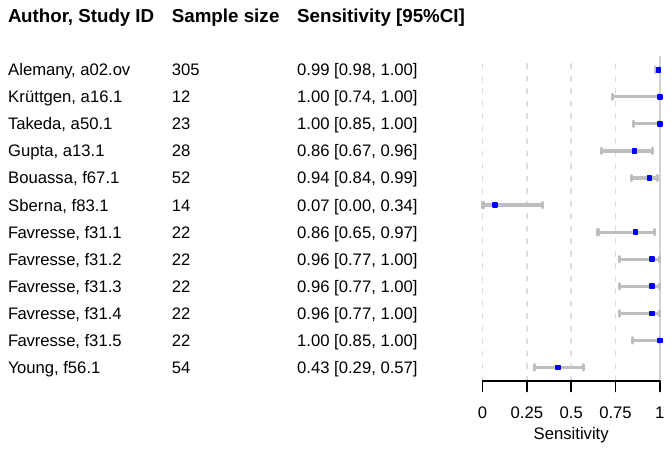

*Caption: CI = confidence interval*

Figure 14 - forest plot for CT values lower 20

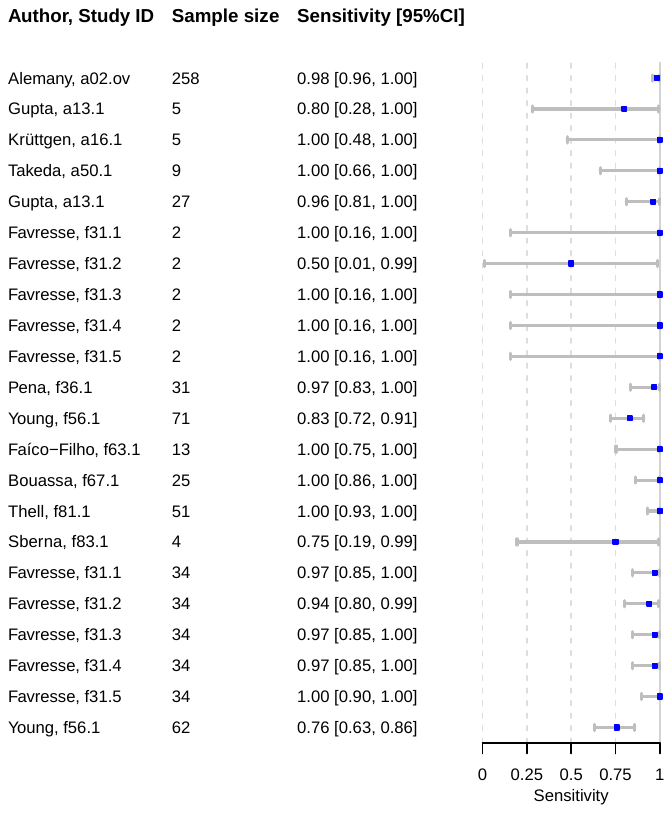

*Caption: CI = confidence interval*

Figure 15 - forest plot for CT values greater 25

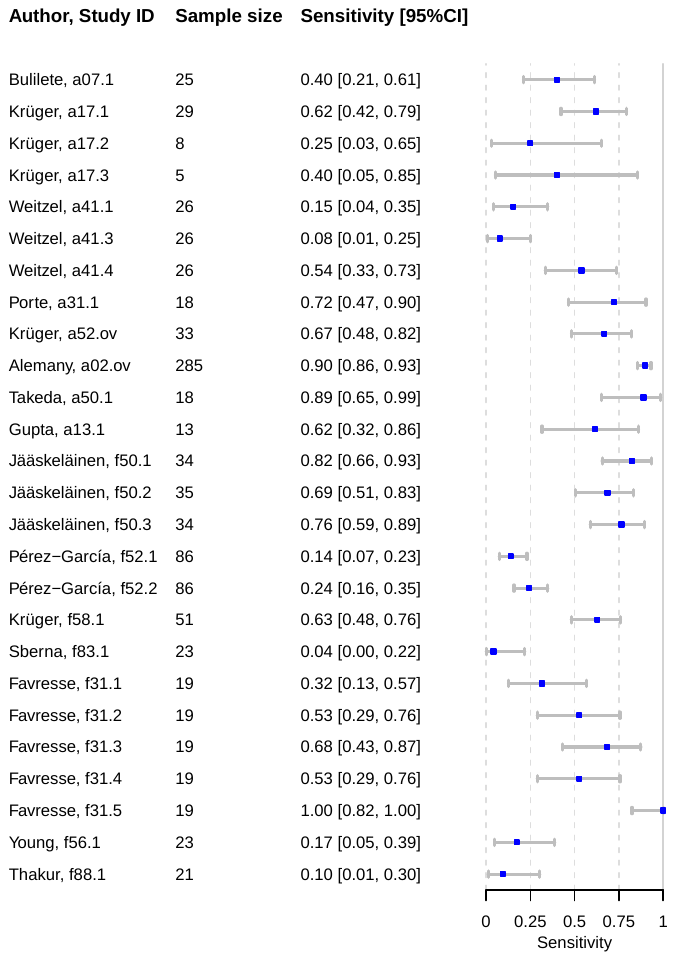

*Caption: CI = confidence interval*

Figure 16 - forest plot for CT values lower 25

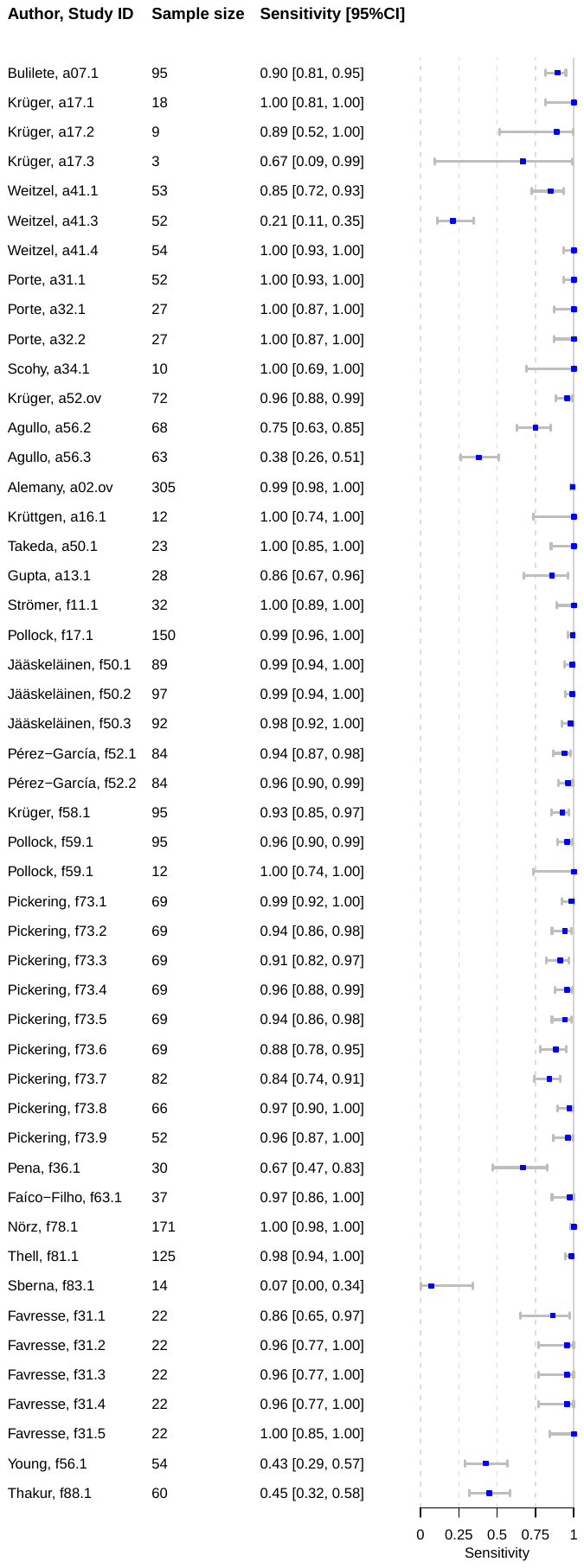

*Caption: CI = confidence interval*

Figure 17 - forest plot for CT values greater 30

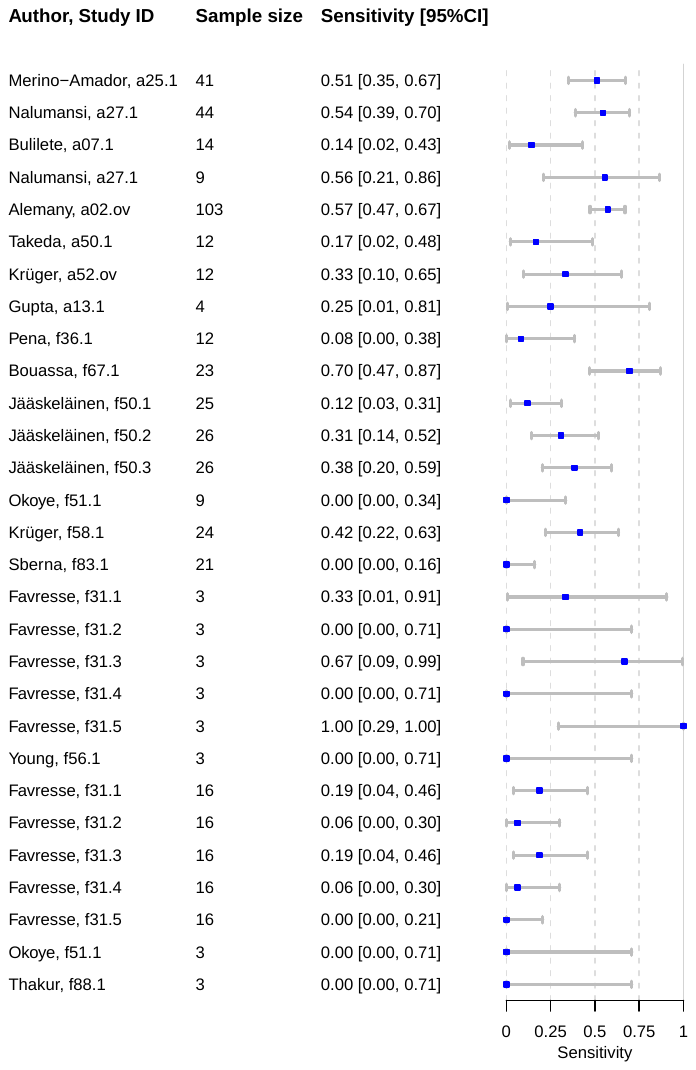

*Caption: CI = confidence interval*

Figure 18 - forest plot for CT value lower 30

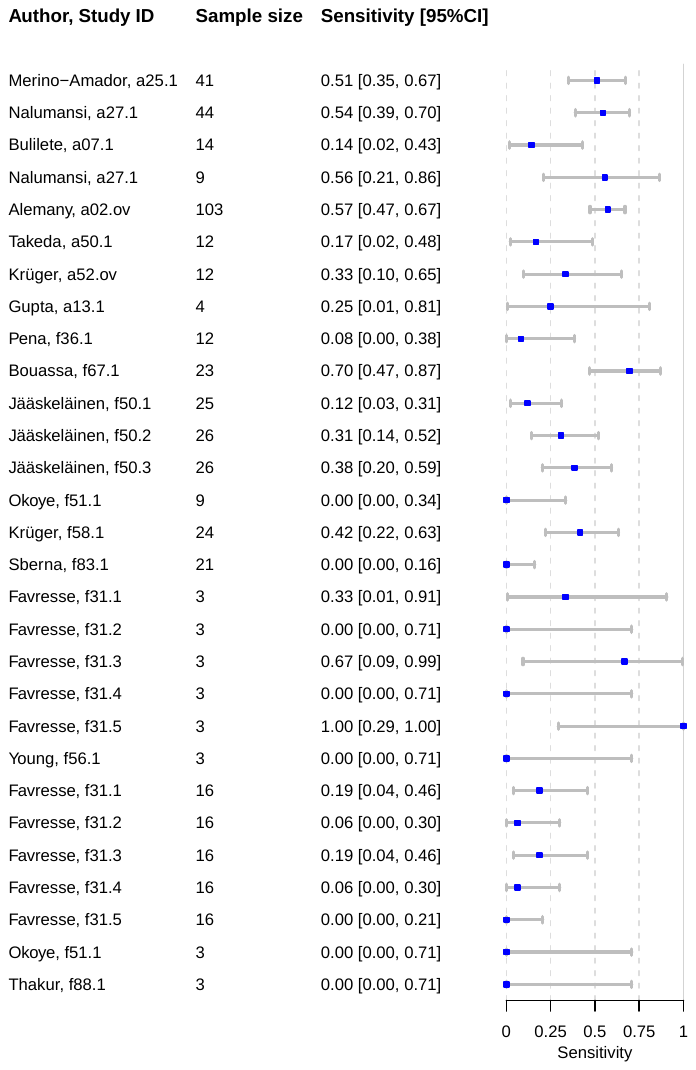

*Caption: CI = confidence interval*

#### S11 Forest plots for subgroups analysis by IFU vs. non-IFU

Figure 19 - Forest Plot for IFU conforming studies

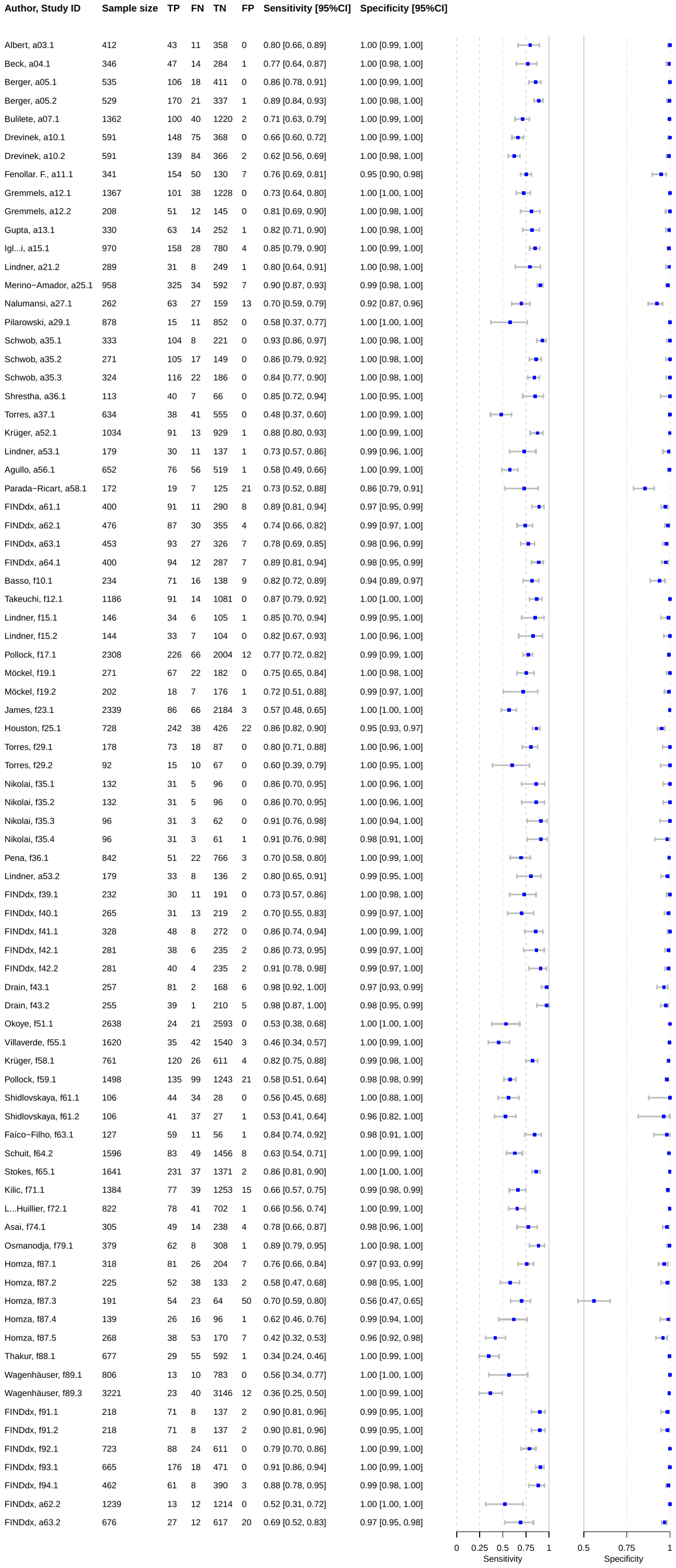

*Caption: TP = true positive; FP = false positive; FN = false negative; TN = true negative; CI = confidence interval*

Figure 20 - Forest plot for non-IFU conforming studies

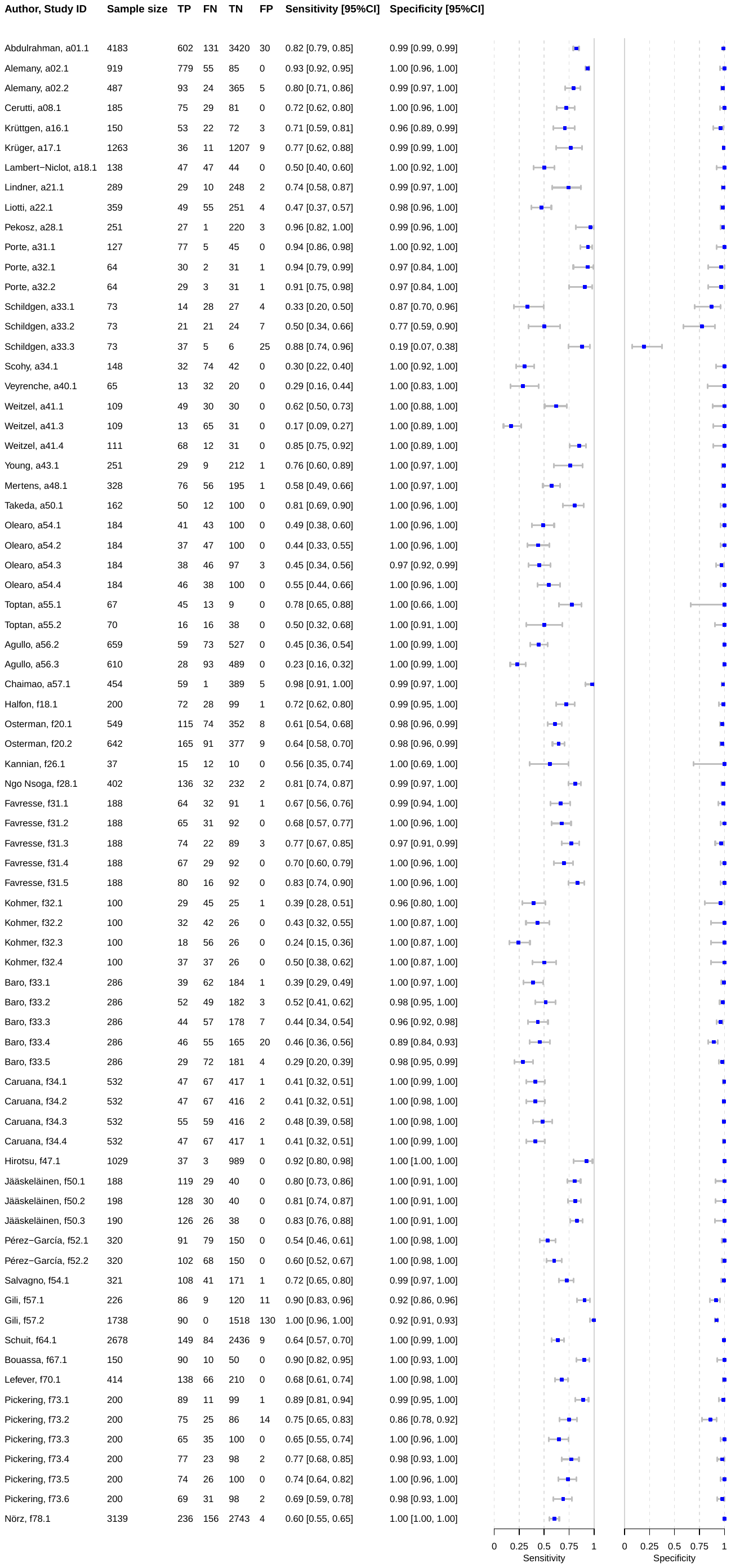

*Caption: TP = true positive; FP = false positive; FN = false negative; TN = true negative; CI = confidence interval*

#### S12 Funnel plot tests

Figure 21 - Funnel plot test for all data sets included in the meta-analysis

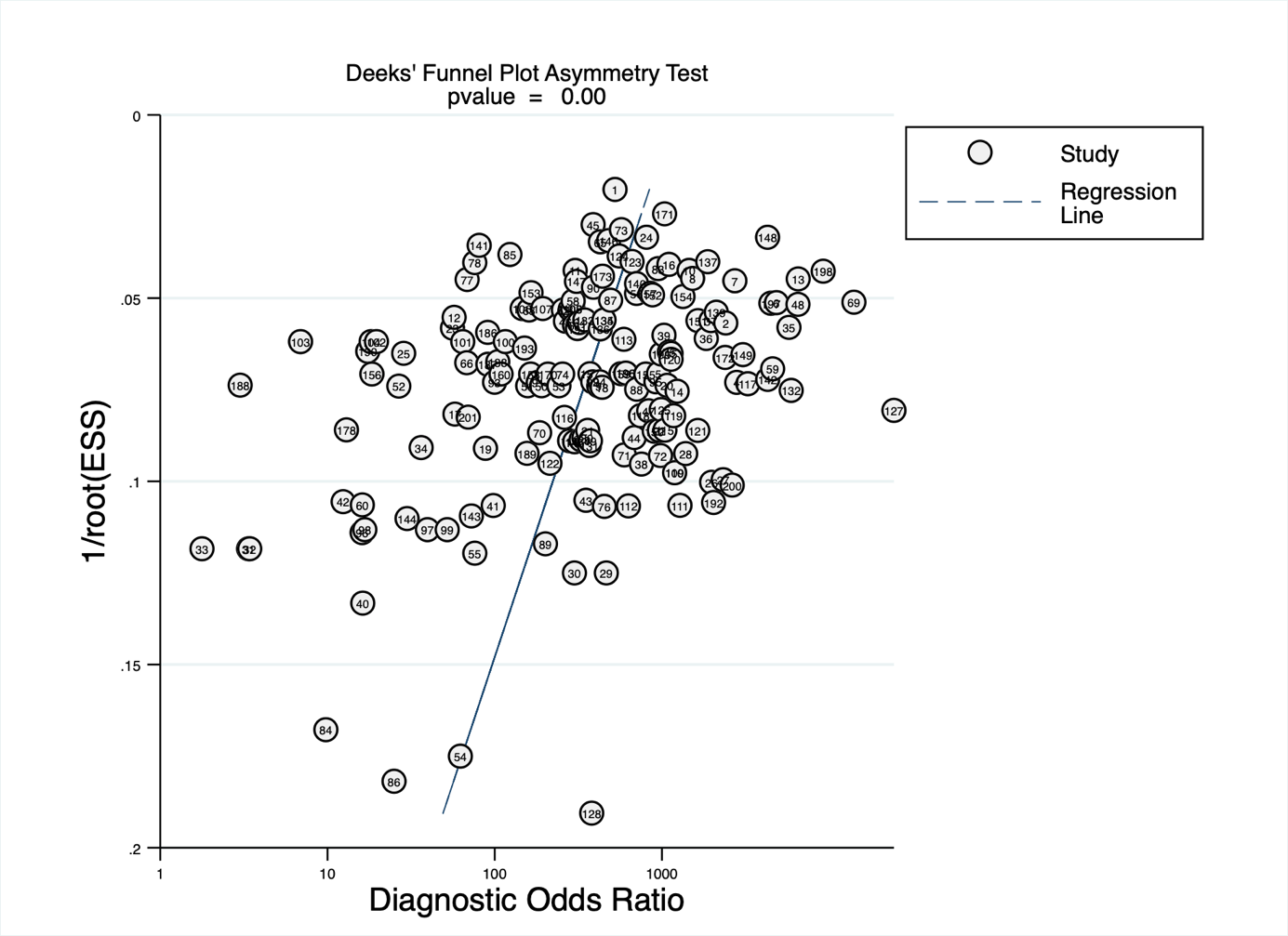

Figure 22 - Funnel plot test for Panbio data sets

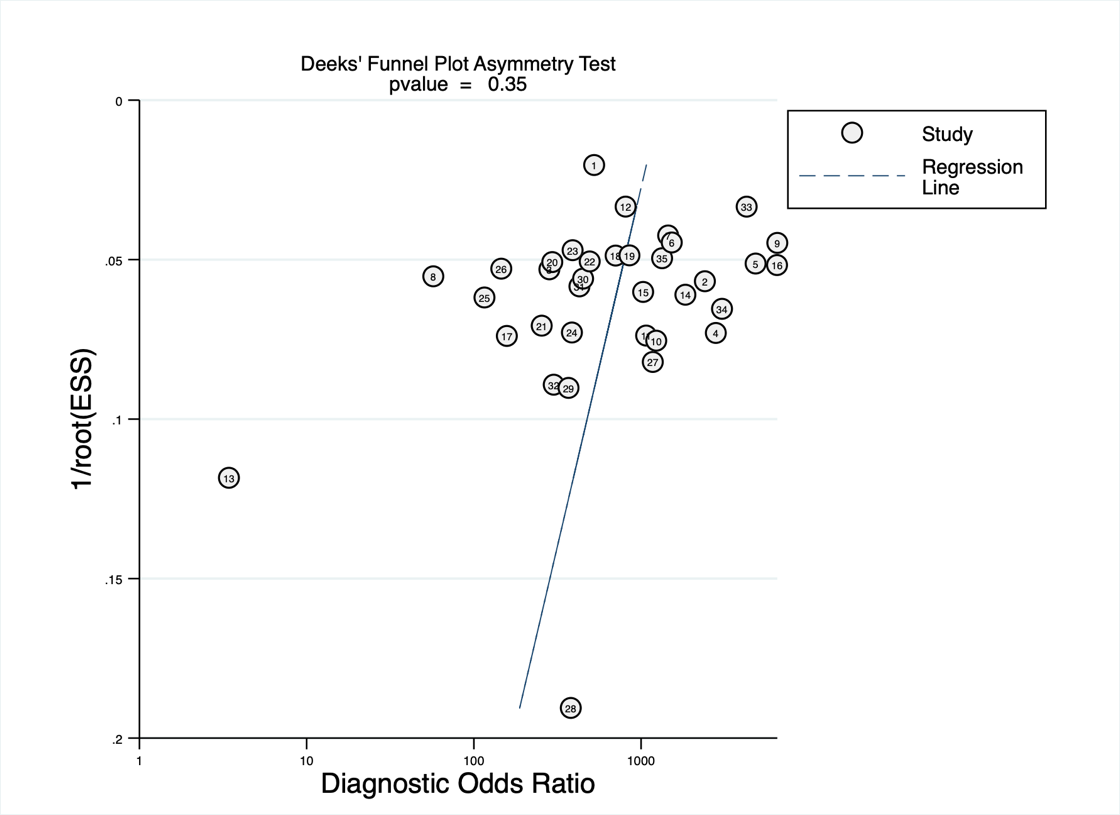

Figure 23 - Funnel plot test for Standard Q data sets

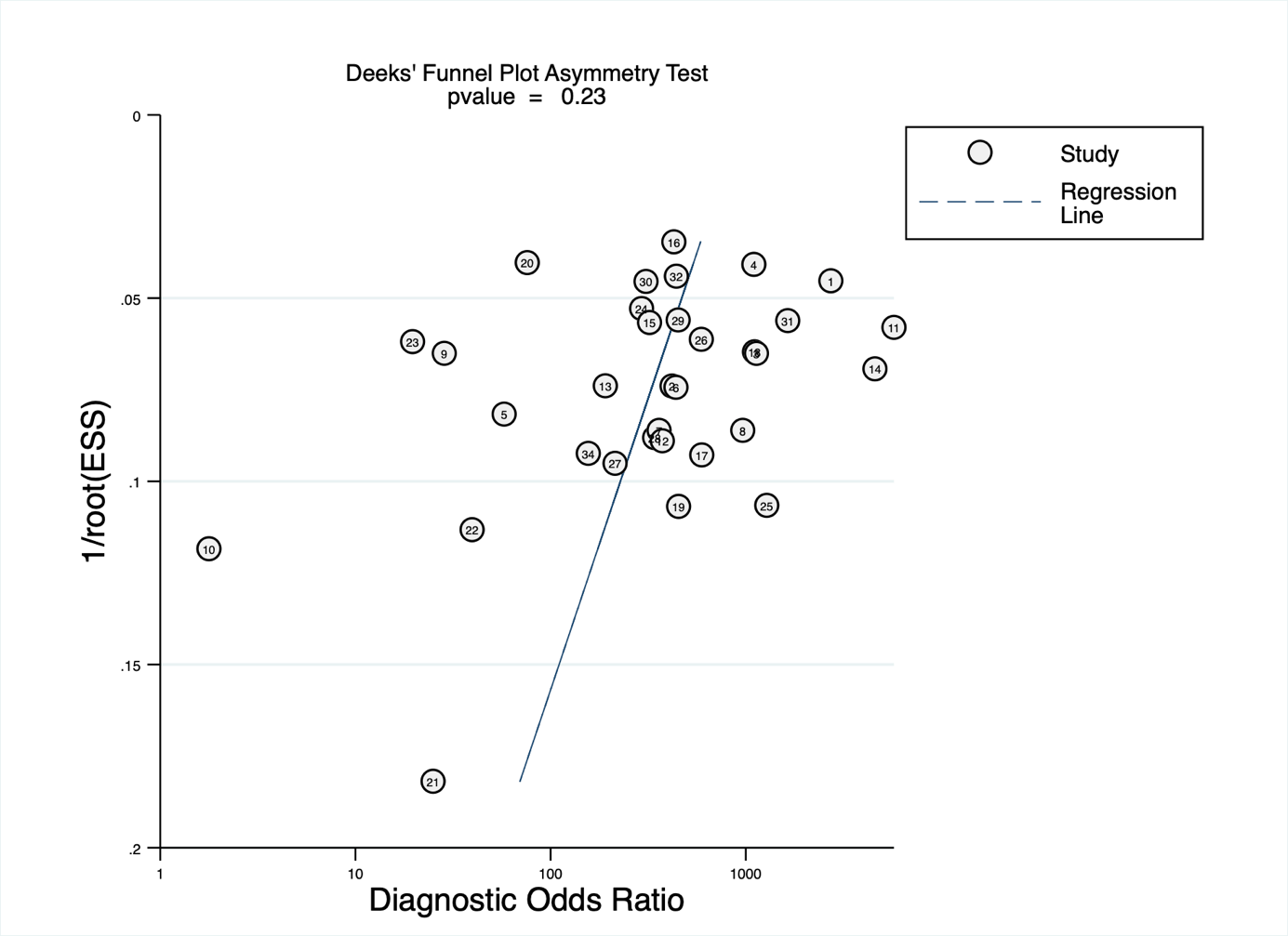

Figure 24 - Funnel plot test for LumiraDx data sets

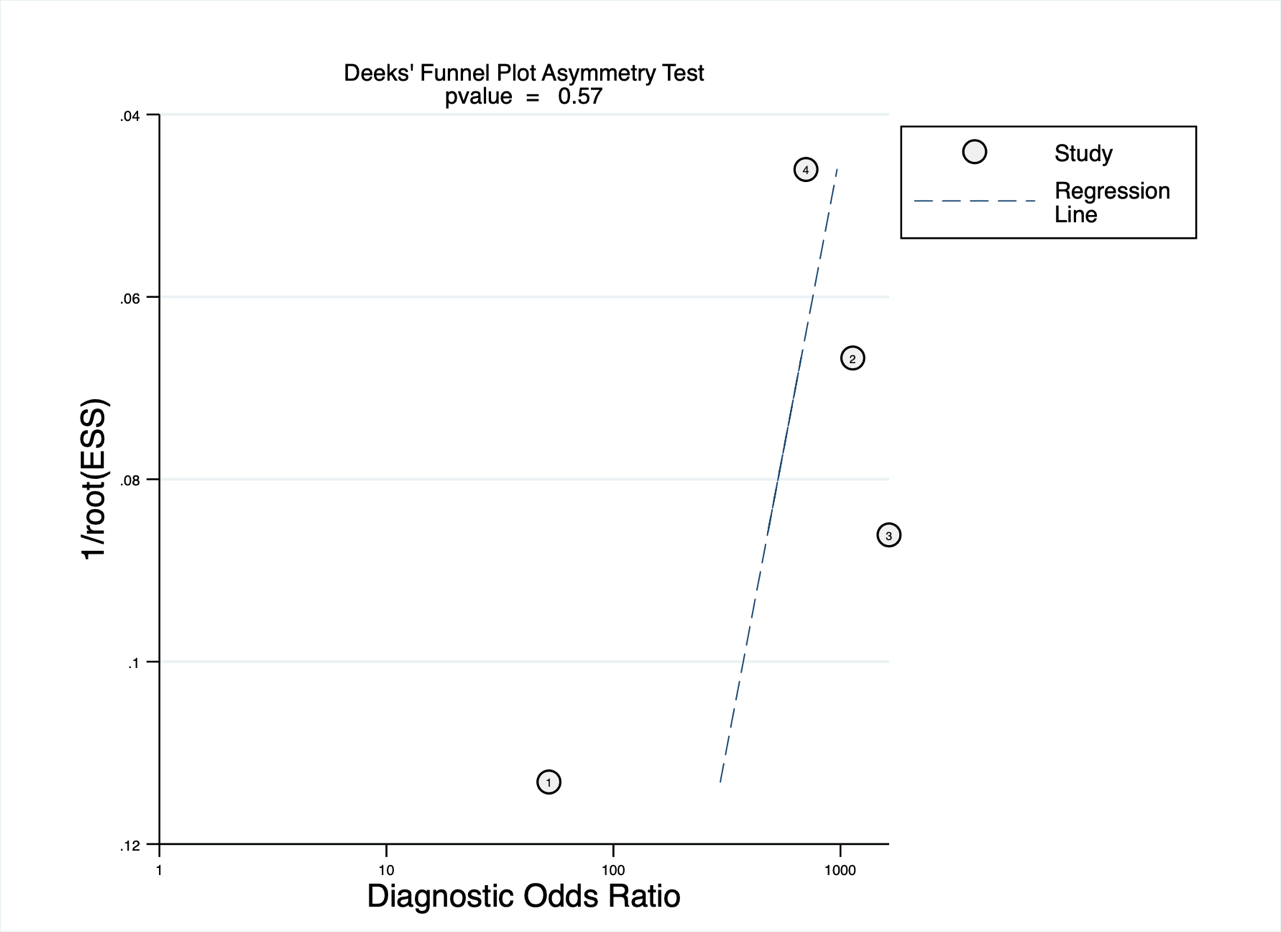

#### S13 Summary of analytical studies

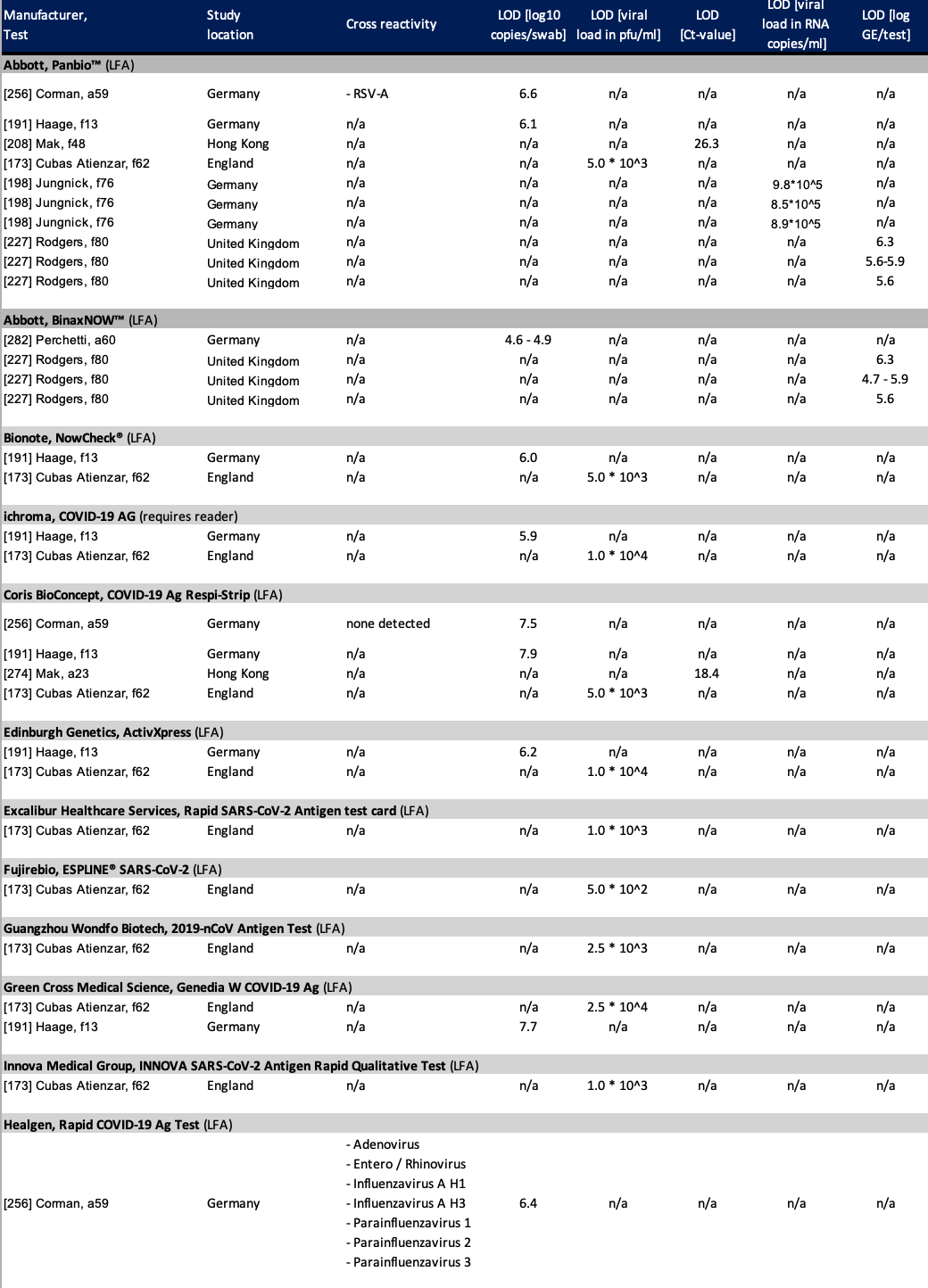

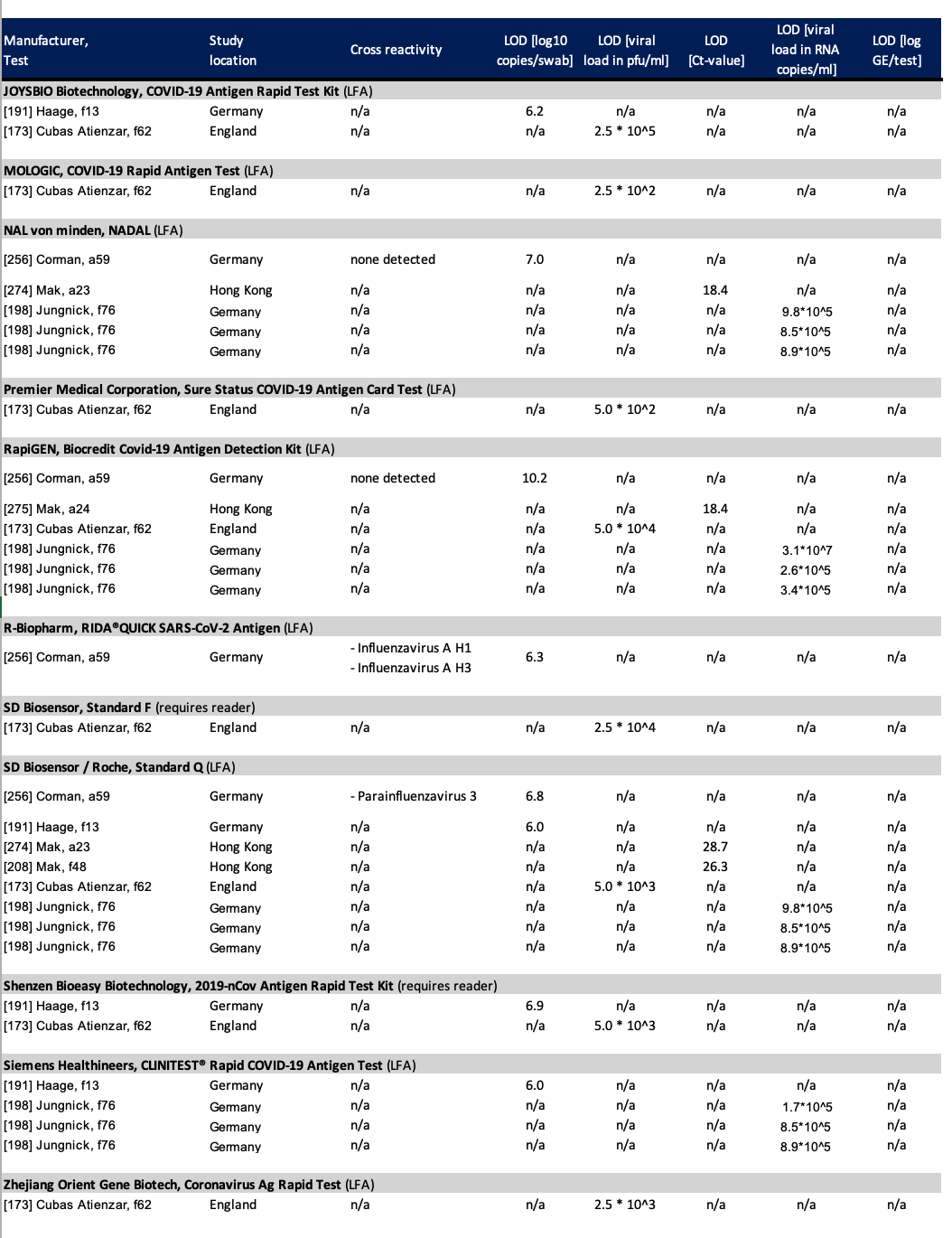
*Caption: LOD = limit of detection; LFA = lateral flow assey; pfu = plaque forming units*

#### S14 List of data items extracted from studies

|  | **Item extracted** | **Data format extracted** |
| --- | --- | --- |
| General | Test assessed | Test Name |
|  | Type of study | Cohort, nested cohort, case-control, cross sectional, randomized, analytic |
|  | Time frame | Retrospective, prospective |
|  | Study location | Country |
|  | Type of location | Clinical, testing site, unclear, other |
|  | Study population | Adult, children |
|  | Screening criteria | Symptomatic, high risk contact, screening independent of symptoms |
|  | Age (median) | Years |
|  | Age (mean) | Years |
|  | Age (lower Range) | Years |
|  | Age (upper Range) | Years |
|  | Age (IQR 1) | Years |
|  | Age (IQR 3) | Years |
|  | Age (STDev) | Years |
|  | Gender | Number of female |
|  | Reference Standard | RT-PCR, viral culture, other |
|  | Type of RT-PCR | Name of RT-PCR |
|  | Type of sample | NP, OP, AN, saliva, BAL/TW, mixed samples |
|  | Type of sample (PCR) | NP, OP, AN, saliva, BAL/TW, mixed samples |
|  | Sampling method | IFU, non-IFU |
|  | Transport medium | Name of medium |
|  | Sample condition | Fresh, cooled, frozen |
| Clinical data | Population size | numeric value |
|  | # of asymptomatic | numeric value |
|  | # of symptomatic | numeric value |
|  | # of hospitalized | numeric value |
|  | # of children | numeric value |
|  | # of adults | numeric value |
|  | CT value (median) | numeric value |
|  | CT value (mean) | numeric value |
|  | CT value (lower range) | numeric value |
|  | CT value (upper range) | numeric value |
|  | CT value (IQR 1) | numeric value |
|  | CT value (IQR 3) | numeric value |
|  | CT Value (STDev) | numeric value |
|  | Viral load (median) | numeric value |
|  | Viral load (mean) | numeric value |
|  | Viral load (Range) | numeric value |
|  | Viral load (IQR 1) | numeric value |
|  | Viral load (IQR 3) | numeric value |
|  | Viral load (STDev) | numeric value |
|  | Duration of symptoms (median) | days |
|  | Duration of symptoms (mean) | days |
|  | Duration of symptoms (lower range) | days |
|  | Duration of symptoms (upper range) | days |
|  | Duration of symptoms (IQR 1) | days |
|  | Duration of symptoms (IQR 3) | days |
|  | Duration of symptoms (STDev) | days |
| Accuracy | True positives | numeric value |
|  | False positives | numeric value |
|  | True negatives | numeric value |
|  | False negatives | numeric value |
|  | Sensitivity | numeric value |
|  | Lower 95 CI sensitivity | numeric value |
|  | Upper 95 CI sensitivity | numeric value |
|  | Specificity | numeric value |
|  | Lower 95 CI specificity | numeric value |
|  | Upper 95 CI specificity | numeric value |
| Analytical | LOD by Ct value | numeric value |
|  | LOD by viral load | numeric value |
|  | viral load UNIT | numeric value |
|  | test-specific values (eg COI for Standard F) | numeric value |

#### S15 Study protocol submitted to PROSPERO

1. **TITLE**

The accuracy of novel antigen-based diagnostics for point of care testing against SARS-CoV-2: a systematic review and meta-analysis.

2. **BRIEF BACKGROUND AND RATIONALE => Introduction ~260 words**

As the COVID-19 pandemic continues around the globe, antigen point of care (POC) diagnostics for SARS-CoV-2 become an important pillar to fight the virus’ spread.^[1]^ An increasing number of such diagnostics appear on the market.^[2]^

For decision-makers to identify well performing diagnostics, independent studies on the tests’ accuracy and ease-of-use are key.^[3]^ While some diagnostics have already been evaluated by multiple of such studies, others still solely rely on the information provided by their manufacturer. This unclear research landscape makes it difficult to judge the to be expected performance of the offered tests.

With this planned systematic review, we will give a structured overview on the currently published accuracy evaluations of SARS-CoV-2 antigen POC diagnostics. We will evaluate the quality of the studies and also discuss any information that is available on the tests’ ease of use.

Where possible, we will perform a meta-analysis, summarizing the sensitivity and specificity of tests that have been evaluated by multiple studies. Furthermore, we will analyze aspects likely to affect the index tests’ accuracy (e.g. duration of symptoms).

3. **REVIEW QUESTION**

To assess the accuracy and ease-of-use of marketable antigen point of care diagnostics for SARS-CoV-2 compared to RT-PCR based on manufacturer independent evaluations.

4. **Criteria for considering studies for the review:**
   a. **Types of studies (designs):**

Clinical and analytical studies evaluating the accuracy of an Ag-RDTs for SARS CoV 2 detection, developed for POC use and comercially available, against a RT-PCR as reference standard. We will consider retrospective or prospective cohort or nested cohort studies, case-control or cross-sectional studies, as well as randomized studies. Publications with less than 10 samples will be excluded. To present the latest available data, we will consider both peer reviewed publications and preprints as eligible.

b. **Types of participants:**

Patients of all age-groups suspected of having or confirmed with SARS-CoV-2. All countries. We will exclude any studies in which patients are being tested for monitoring or end of quarantine. We will exclude an populations of less than 10 participants. Although the size threshold of 10 is arbitrary, such small studies are likely to give unreliable estimates of sensitivity or specificity.

c. **Index test:**

Antigen rapid diagnostic test (ag-RDT) for SARS-CoV-2, developed for POC use and commercially available

d. **Target conditions**

Persons presumed to have COVID-19, independent of symptoms

e. **Reference Standard:**

For the purposes of this review, we considered RT-PCR to be the ‘reference standard’ against which the rapid tests are compared. RT-PCR is the most direct reference standard for an antigen test as RNA to be highly correlated with Antigen quantities.^[4]^ The result of further RT-PCR analysis of discrepant cells (samples with results disagreeing on the rapid test and the RT-PCR) will also be considered in sensitivity analyses. As discrepant analysis involves retesting only a subsample of patients selected according to index and reference standard results, it can introduce bias. ^[5]^Furthermore, we will consider results from retesting with a second test in a sensitivity analysis.

5. **Criteria for considering studies for the METANANLYSIS:**

We will only consider all studies that have been included into the systematic review as eligible for the meta-analysis. We will provide separate meta-analysis for analytical and clinical accuracy. In addition, we will only provide a test specific meta-analysis for index tests that have been evaluated by at least 4 studies.

6. **SEARCH METHODS**

We performed an electronic search in the databases PubMed, Web of Science, medRxiv and bioRxiv using an automated search algorithm. The main search terms were “Severe Acute Respiratory Syndrome Corona-virus 2”, “COVID-19”, “Betacoronavirus”, “Coronavirus” and “Point of Care Testing”. A librarian defined the search. We searched for any literature published between December 01^st^, 2019 and December 11^th^, 2020. No language restrictions were applied.

7. **MAIN OUTCOMES**
   a. **Outcome**

The number of evaluations per index tests, the range of the index tests sensitivity and specificity across the evaluations.

b. **Measures of effect**

A table summarizing the main results from all studies meeting the inclusion criteria will be presented. Results will be presented in four categories: general information, clinical data, accuracy and analytical data. It will also show the assessment of studies, in whether the respective research was independent from the test manufacturers and the quality of its methods (QUADAS, see below).

The general section will include the test assessed, type of study, study location, type of location, study population, screening criteria, age, gender, type of RT-PCR, type of sample, sampling method, transport medium and sample condition. The clinical data will include values such as the population size, number of asymptomatic and symptomatic or the number of hospitalized. The accuracy section will include the number of true positives, false positives, true negatives, false negatives, and sensitivity and specificity with 95% Confidence Intervals. The analytical section will include LOD by Ct value, LOD by viral load, viral load UNIT and any test-specific values.

8. **ADDITIONAL OUTCOMES**
   a. **Outcome**

In addition to the main outcomes, we will also perform investigations of heterogeneity, sensitivity analyses and evaluate the existence of a publication bias.

b. **Measures of effect**

A table summarizing the main results from all studies meeting the inclusion criteria will be presented. Results will be presented in four categories: general information, clinical data, accuracy and analytical data. It will also show the assessment of studies, in whether the respective research was independent from the test manufacturers and the quality of its methods (QUADAS assessment, see Question 27).

The general section will include the test assessed, type of study, study location, type of location, study population, screening criteria, age, gender, type of RT-PCR, type of sample, sampling method, transport medium and sample condition. The clinical data will include values such as the population size, number of asymptomatic and symptomatic or the number of hospitalized. The accuracy section will include the number of true positives, false positives, true negatives, false negatives, and sensitivity and specificity with 95% Confidence Intervals. The analytical section will include LOD by Ct value, LOD by viral load, viral load UNIT and any test-specific values.

9. **REVIEW METHODS**
   a. **Study selection methods:**

Two independent reviewers (LEB and CE or LEB and SS) will review the titles and abstracts of all publications returned by the search algorithm (screen 1). Any publication selected for inclusion by one of the reviewers will be considered as potentially eligible. All potentially eligible publications will be reviewed again in detail by the same reviewers (screen 2). After the detailed review, both authors will independently decide which publications to include into the systematic review. Any disputes will be solved by discussion or by a third reviewer (CMD).

b. **Data extraction methods:**

We will extract only data for Ag-RDT probes that were collected at the same time as the RT-PCR sample. If multiple different RT-PCR tests were performed on one sample within one study, we will utilize the data from the best-performing assay.

If a study contributed data to more than one analysis (e.g., two different alternative sample types in one study used for the Ag-RDT), it will be considered as two or more datasets. In addition to data on test performance, we will extract data for factors likely to affect test performance: (1) asymptomatic versus symptomatic; (2) symptom duration prior to testing (3); Type of RT-PCR (4); populations (e.g. pediatric versus adults); (5) site of sampling (e.g. low prevalence setting vs. diagnostic use case); (6) sampling as per IFU vs. other sampling methods; (7) sample condition (e.g. fresh vs. frozen sample).

Where possible, we will separately extract data according to viral load and Ct-value, and for molecular assays, before and after re-analysis of samples in discrepant cells.

c. **Quality assessment methods [risk of bias in individual studies] and how quality data will be used:**

We will assess the quality of each study by applying the QUADAS-2 tool.^[6]^ The tool consists out of four domains: patient selection, index test, reference standard, and flow and timing. For each domain, the risk of bias is analyzed using different signaling questions. Beyond the risk of bias, the tool also evaluates the applicability of the study design to the research question for every domain. We have adapted the tool for the present review as follows:

**Domain 1 Patient Selection:**

Risk of Bias: Could the selection of patients have introduced bias?

•	Signaling question 1: Was a consecutive or random sample of patients or specimens enrolled?

Score ‘yes’ if the study enrolled a consecutive or random sample of eligible patients; ‘no’ if the study selected patients by convenience, and ‘unclear’ if the study did not report the manner of patient selection or unable to tell.

•	Signaling question 2: Was a case-control design avoided?

We scored ’yes’ to all included studies given that we are excluding case-control study designs.

•	Signaling question 3: Did the study avoid inappropriate exclusions?

We scored ’yes’ to studies which included: all participants a) regardless of symptoms or b) duration of symptoms and c) prior testing (e.g. CXR). We scored ’no’ if studies excluded participants on the basis of symptoms or duration of symptoms or prior testing. We scored ’unclear’ if we could not tell.

- Risk of Bias is scored ‘low concern’ if studies score ‘yes’ on all the question, ‘unclear concern’ if questions are answered with ‘yes’ and ‘unclear’, ‘intermediate concern’ if one question is answered with ‘no’, ‘high concern’ if two or more questions are answered with ‘no’.

Applicability: Are there concerns that the included patients and setting do not match the review question? We were interested in how Ag-RDT performs in patients whose specimens were evaluated as they would be in routine practice. We expected to score most studies as ’low concern’ since we planned to determine test accuracy only for COVID diagnosis. We scored ‘high concern’ if Ag-RDT were evaluated for end of quarantine evaluation or monitoring.

**Domain 2: Index Test**

Risk of Bias: Could the conduct or interpretation of the index test have introduced bias?

•	Signaling question 1: Were the index test results interpreted with knowledge of the results of the reference standard?

We answered ’yes’ if the study interpreted the result of Ag-RDT blinded to the result of the reference standard; we answered ’no’ if the study did not interpret the result of Ag-RDT blinded to the result of the reference standard. We answered ’yes’ for studies in which Ag-RDT was performed on fresh specimens, since reference standard results would be unavailable at the time of test interpretation. We answered ’unclear’ if stored specimens were tested or we could not tell if the index test results were interpreted without knowledge of the reference standard results.

•	Signaling question 2: If a threshold was used, was it prespecified?

We answered ’yes’ if the threshold was prespecified or if the tests was performed by IFU. We scored ’no’ if the threshold was not prespecified, and ’unclear’ if we could not determine if the threshold was prespecified or not.

- Risk of Bias is scored ‘low concern’ if studies score ‘yes’ on all the question, ‘unclear concern’ if questions are answered with ‘yes’ and ‘unclear’, ‘intermediate concern’ if one question is answered with ‘no’, ‘high concern’ if two or more questions are answered with ‘no’.

Applicability: Are there concerns that the index test, its conduct, or its interpretation differ from the review question? If index test methods vary from those specified in the review question, concerns about applicability may exist. We judged ’high concern’ if the test procedure was inconsistent with the manufacturer recommendations, ’low concern’ if the test procedure was consistent with the manufacturer recommendations, and ’unclear concern’ if we could not tell.

**Domain 3: Reference Standard**

Risk of Bias: Could the reference standard, its conduct, or its interpretation have introduced bias?

•	Signaling question 1: Is the reference standard likely to correctly classify the target condition?

NAAT is considered to be the gold standard for COVID. However, the accuracy of this reference standard is not 100%, especially late in the disease and it varies widely across the different non-respiratory samples. However, given that viral loads measured in NAATs correlate well with Antigen, we scored ‘yes’ for all studies included.

•	Signaling question 2: Were the reference standard results interpreted without knowledge of the results of the index test?

We scored ‘yes’ if the test was performed immediately at the POC on fresh samples and ‘no’ if performed in the laboratory on stored samples unless blinding was specifically reported. We scored ‘unclear’ if we could not tell.

- Risk of Bias is scored ‘low concern’ if studies score ‘yes’ on all the question, ‘unclear concern’ if questions are answered with ‘yes’ and ‘unclear’, ‘intermediate concern’ if one question is answered with ‘no’, ‘high concern’ if two or more questions are answered with ‘no’.

Applicability: Are there concerns that the target condition as defined by the reference standard does not match the question? We judged applicability to be of ‘low concern’ for all studies.

**Domain 4: Flow and Timing**

Risk of Bias: Could the patient flow have introduced bias?

•	Signaling question 1: Was there an appropriate interval between the index test and reference standard?

We expected specimens for Ag-RDT and the reference standards to be obtained at the same time and answered ’yes’ for all studies that meet these criteria. We answered ’no’ if specimens were collected for index and reference standard tests greater than 24h apart, and ’unclear’ if we could not tell.

•	Signaling question 2: Did all patients receive the same reference standard?

Answer this question ‘yes’ if all studies used the same reference standard (acceptable reference standard as specified as a criterion for inclusion in the review), answer ‘no’ if different reference standards were used.

•	Signaling question 3: Were all patients included in the analysis?

We determined the answer to this question by comparing the number of participants enrolled in the study with the number of participants included in the two-by-two tables. We answered ’yes’ if all participants enrolled in the study were tested with results presented and accounted for. We answered ’no’ if participants meeting enrolment criteria were not tested or results were not presented, and ’unclear’ if we could not tell.

- Risk of Bias is scored ‘low concern’ if studies score ‘yes’ on all the question, ‘unclear concern’ if questions are answered with ‘yes’ and ‘unclear’, ‘intermediate concern’ if one question is answered with ‘no’, ‘high concern’ if two or more questions are answered with ‘no’.

10. **STRATEGY FOR DATA SYNTHESIS**

We present estimates of sensitivity and specificity for each test brand using paired forest plots, and summarize results using average sensitivity and specificity in tables as appropriate. We estimate summary sensitivities and specificities with 95% confidence intervals (CI) using the bivariate model, via the meqrlogit command of Stata/SE 16.0. Where studies presented only estimates of sensitivity, we will fit univariate random effects logistic regression models.

Where adequate data is available for the subcategories, we will investigate heterogeneity by including indicator variables in the random-effects logistic regression models. Absolute differences between the sensitivity or specificity and the P values are reported from the model

11. **ANALYSIS OF SUBGROUPS OR SUBSETS**

We will analyze the following subgroups to find out whether they have any impact on a test’s sensitivity and specificity:

1) asymptomatic versus symptomatic

2) symptom duration prior to testing

3) Type of RT-PCR

4) populations (e.g. pediatric versus adults)

5) site of sampling (e.g. low prevalence setting vs. diagnostic use case)

6) sampling as per IFU vs. other sampling methods

7) sample condition (e.g. fresh vs. frozen sample)

8) viral load and Ct-value

9) for Ag-RDT with four or more publications: point estimate for sensitivity and specificity

We plan to analyze these by heterogeneity as stated above.

**SOURCES**

Federal Institute for Drugs and Medical Devices (2020). Antigentests zum direkten Erregernachweis des Coronavirus SARS-CoV-2. Online at: https://antigentest.bfarm.de/ords/antigen/r/antigentests-auf-sars-cov-2/liste-der-antigentests?session=13253028659507&tz=1:00.

Hadgu, A. (1999). Discrepant analysis: a biased and an unscientific method for estimating test sensitivity and specificity. *Journal of clinical epidemiology*, *52*(12), 1231-1237.

Pollock, N. R., Savage, T. J., Wardell, H., Lee, R., Mathew, A., Stengelin, M., & Sigal, G. B. (2020). Correlation of SARS-CoV-2 nucleocapsid antigen and RNA concentrations in nasopharyngeal samples from children and adults using an ultrasensitive and quantitative antigen assay. *medRxiv*.

Reitsma, J. B., Glas, A. S., Rutjes, A. W., Scholten, R. J., Bossuyt, P. M., & Zwinderman, A. H. (2005). Bivariate analysis of sensitivity and specificity produces informative summary measures in diagnostic reviews. *Journal of clinical epidemiology*, *58*(10), 982-990.

Whiting, P., Rutjes, A. W., Reitsma, J. B., Bossuyt, P. M., & Kleijnen, J. (2003). The development of QUADAS: a tool for the quality assessment of studies of diagnostic accuracy included in systematic reviews. *BMC medical research methodology*, *3*(1), 25.

World Health Organization. (2020a). *Advice on the use of point-of-care immunodiagnostic tests for COVID-19: scientific brief, 8 April 2020* (No. WHO/2019-nCoV/Sci_Brief/POC_immunodiagnostics/2020.1). World Health Organization.

World Health Organization. (2020b). *Antigen-detection in the diagnosis of SARS-CoV-2 infection using rapid immunoassays: interim guidance, 11 September 2020* (No. WHO/2019-nCoV/Antigen_Detection/2020.1). World Health Organization.

1. World Health Organization 2020b.
2. Federal Institute for Drugs and Medical Devices 2020.
3. World Health Organization 2020a.
4. Pollock 2020.
5. Hadgu 1999.
6. Whiting et al. 2003.
